# A graphSAGE discovers synergistic combinations of Gefitinib, paclitaxel, and Icotinib for Lung adenocarcinoma management by targeting human genes and proteins: the RAIN protocol

**DOI:** 10.1101/2024.04.14.24304384

**Authors:** Sogand Sadeghi, Ali A. Kiaei, Mahnaz Boush, Nader Salari, Masoud Mohammadi, Danial Safaei, Mitra Mahboubi, Arian Tajfam, Sabra Moghadam

## Abstract

**Background:** Adenocarcinoma of the lung is the most common type of lung cancer, and it is characterized by distinct cellular and molecular features. It occurs when abnormal lung cells multiply out of control and form a tumor in the outer region of the lungs. Adenocarcinoma of the lung is a serious and life-threatening condition that requires effective and timely management to improve the survival and quality of life of the patients. One of the challenges in this cancer treatment is finding the optimal combination of drugs that can target the genes or proteins that are involved in the disease process.

**Method:** In this article, we propose a novel method to recommend combinations of trending drugs to target its associated proteins/genes, using a Graph Neural Network (GNN) under the RAIN protocol. The RAIN protocol is a three-step framework that consists of: 1) Applying graph neural networks to recommend drug combinations by passing messages between trending drugs for managing disease and genes that act as potential targets for disease; 2) Retrieving relevant articles with clinical trials that include those proposed drugs in previous step using Natural Language Processing (NLP). The search queries include “Adenocarcinoma of the lung”, “Gefitinib”, “Paclitaxel”, “Icotinib” that searched context based in databases using NLP; 3) Analyzing the network meta-analysis to measure the comparative efficacy of the drug combinations.

**Result:** We applied our method to a dataset of nodes and edges that represent the network, where each node is a drug or a gene, and each edge is a p-value between them. We found that the graph neural network recommends combining Gefitinib, Paclitaxel, and Icotinib as the most effective drug combination to target this cancer associated proteins/genes. We reviewed the clinical trials and expert opinions on these medications and found that they support our claim. The network meta-analysis also confirmed the effectiveness of these drugs on associated genes.

**Conclusion:** Our method is a novel and promising approach to recommend trending drugs combination to target cancer associated proteins/genes, using graph neural networks under the RAIN protocol. It can help clinicians and researchers to find the best treatment options for patients, and also provide insights into the underlying mechanisms of the disease.

**Highlights:**

- Proposing the combination of medicinal compounds together for the treatment of lung adenocarcinoma
- achieved a p-value of 0.002858 between lung adenocarcinoma and targeted proteins/genes
- 3-Leveraging GraphSAGE for Suggesting an Optimal Drug Combinations.

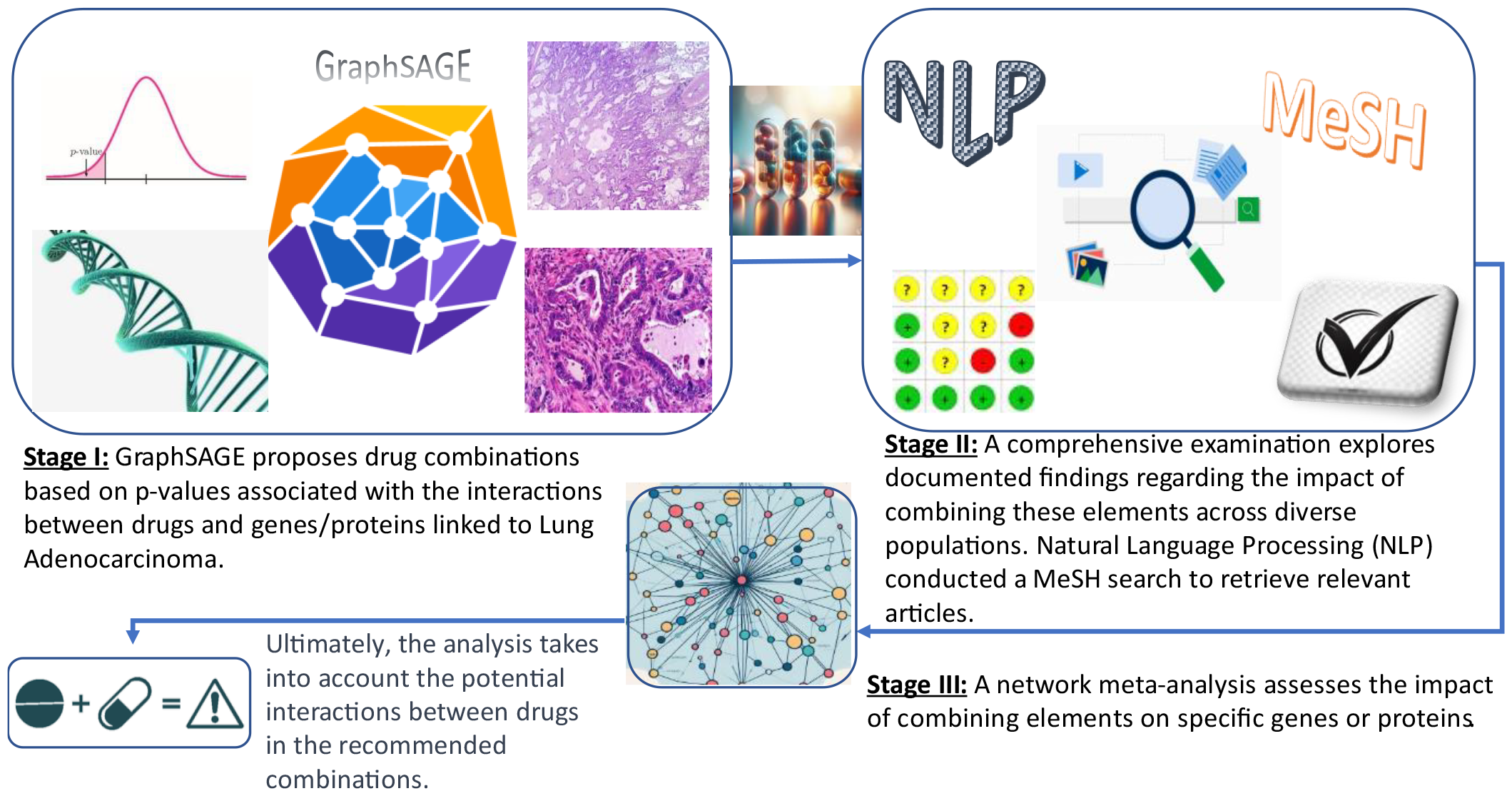

## 1. INTRODUCTION

Adenocarcinoma of the lung is a form of lung cancer that begins in the cells lining the air sacs within the lungs. It represents the most prevalent form of lung cancer, contributing to more than 40% of non-small cell lung cancers. In 2011, the International Association for the Study of Lung Cancer (IASLC), American Thoracic Society (ATS), and European Respiratory Society (ERS) established a novel, global histopathologic classification for lung adenocarcinoma. The new classification includes adenocarcinoma in situ, minimally invasive adenocarcinoma, lepidic predominant adenocarcinoma, and invasive mucinous adenocarcinoma. These classification entities replace the now retired term, bronchoalveolar carcinoma (BAC). The radiographic appearance of these lesions ranges from pure, ground glass nodules to large, solid masses. A thorough understanding of the new classification is essential to radiologists who work with MDT colleagues to provide accurate staging and treatment. ^1^

Lung adenocarcinoma is a serious condition that requires prompt and effective management. The importance of managing lung adenocarcinoma lies in the fact that it is the prevalent form of lung cancer, constituting around 40% of all cases of lung cancer. Timely identification and therapy of lung adenocarcinoma able to significantly enhance the chances of survival and reduce the risk of complications. The World Health Organization (WHO) suggests offering supportive care to individuals with lung cancer for the management of symptoms and the provision of pain relief, and give emotional support. ^2^

According to Yan’s published work in 2023, there are several drug combinations that have shown promise in treating lung adenocarcinoma. Here are some of the benefits of these drug combinations: 1. Improved survival rates: An examination of patients with lung adenocarcinoma in a study group predominantly consisting of never-smoking East Asian women revealed that, in individuals with metastatic non-small cell lung cancer bearing EGFR mutations and without central nervous system metastases, the combination of ramucirumab and erlotinib substantially extended the progression-free survival (PFS) duration. 2. Reduced side effects: A new lung cancer treatment combination approved on NHS in England has shown promising results in reducing the side effects of chemotherapy. The treatment combination includes pembrolizumab, carboplatin, and paclitaxel. 3. Improved quality of existence: Novel medications for lung cancer such as immunotherapy and targeted drugs have extended survival times and enhanced the well-being of individuals with advanced and metastatic non-small cell lung cancer (NSCLC). ^3,4^

### Associated human genes

MALAT1 is a long non-coding RNA (lncRNA) that has been widely studied for its role in various respiratory diseases, including lung adenocarcinoma. According to Wu’s published work in Frontiers in Molecular Biosciences, MALAT1 stimulates the proliferation of lung cancer cells by targeting and controlling the expression of miR-206, thereby activating the PI3K/Akt signaling pathway, and increasing phosphorylation of mammalian rapamycin (mTOR), ribosomal protein S6 kinase 1 (S6K1), and other related proteins. Shi’s published work in BMC Cancer suggests that MALAT1 plays a role in the progression of non-small cell lung cancer by influencing the miR-124/STAT3 axis. MALAT1 has been suggested as a potential indicator for the diagnosis and prognosis of cancer. ^5–10^

The epidermal growth factor receptor (EGFR) is a cellular protein that facilitates cell growth. A mutation in the gene for EGFR can make it grow too much, which can cause cancer. In the United States, approximately 10-15% of lung cancer cases are attributed to EGFR-positive lung cancer and generally appears in adenocarcinoma subtype of non-small cell lung cancer. In relation to how EGFR acts as a potential target for lung adenocarcinoma, Numerous targeted treatments have been created to address lung cancer that is positive for EGFR. These therapies work by blocking the activity of the EGFR protein, which can slow or halt the proliferation of cancerous cells. One such therapy is osimertinib, which is a third-generation EGFR tyrosine kinase inhibitor (TKI) demonstrated effectiveness in addressing EGFR-positive lung cancer. Another therapy is afatinib, a second-generation EGFR tyrosine kinase inhibitor (TKI) that has demonstrated effectiveness in the treatment of EGFR-positive lung cancer. ^11–13^

According to a recent article mutant KRAS activates the expression of CD47, an antiphagocytic signal in cancer cells, leading to decreased phagocytosis of malignant cells by macrophages within adenocarcinoma of the lung patient samples and Kras-driven genetic rodent models of lung cancer. In addition, a review article published on Frontiers in Oncology describes that KRAS mutations can co-exist with other mutations in significant genes in cancer (e. g., STK11 and KEAP1) which induces tumor heterogeneity and promotes different responses to therapies. Another article published on Frontiers in Oncology states that KRAS-activating mutations are the prevailing oncogenic drivers in non-small cell lung cancer (NSCLC); however, multiple efforts to inhibit KRAS proved unsuccessful over the past decade. KRAS mutations are linked to an unfavorable prognosis and limited responsiveness to standard treatment regimens. ^14–17^ the ROS1 gene is altered in about 1-2% of lung cancer patients and generally appears in adenocarcinoma non-small cell lung cancer. ROS1 mutations are predominantly observed in tumors of the adenocarcinoma type that are negative for other driver mutations. Adenocarcinoma tumors frequently exhibit ROS1 mutations that are negative for other driver mutations. Patients who are ROS1-positive tend to be younger than the average lung cancer patient and have little to no smoking history. Zhang’s published work on Nature describes that ROS1 rearrangements present actionable therapeutic targets for non-small cell lung cancer (NSCLC). ^18–22^

Chen’s published work on Oncotarget describes that NKX2-1 may suppress lung adenocarcinoma progression via impeding TGF-β-induced epithelial-to-mesenchymal transition (EMT) due to Elevated expression of E-cadherin and occludin targeted by NKX2-1 impeded migration and induced apoptosis in lung cancer cells. ^23–27^

According to Moeller’s published work in Cancer Control journal, patients with lung adenocarcinoma not expressing TTF1 have a worse prognosis. However, the same study also suggests that the administration of the use of immunotherapy in individuals harboring a KRAS mutation and the change from a regimen containing pemetrexed to a regimen containing no pemetrexed, the corresponding patients no longer seem to have a worse prognosis. ^28–30^

Que’s published work in Frontiers in Oncology journal that describes an individual diagnosed with advanced lung adenocarcinoma exhibiting ALK positivity attained a pathological complete remission (pCR) in the primary lung lesion following various rounds of conversion therapy. utilizing immunotherapy following multiple lines of treatment in advanced lung adenocarcinoma with ALK positivity may provide a survival benefit. Akhoundova’s published work in the same journal that discusses the involvement of ALK in lung neuroendocrine tumors.^31–35^

According to Zhang’s published work in the Journal of Thoracic Disease suggests that NAPSA serves as a valuable indicator for identifying lung adenocarcinoma. Li’s published work in the same journal suggests that the presence of NAPSA expression is linked to a more favorable prognosis in individuals diagnosed with lung adenocarcinoma. ^36,37^

According to Zhou’s published work in Frontiers in Genetics journal that suggests CD274 and PDCD1LG2 are potential targets for immunotherapy in pan-cancer. CD274 and PDCD1LG2 not only impact the prognosis of patients with cancer but are also associated with various immunosuppressive biomarkers, such as CTLA4, TIGIT, and LAG3. Furthermore, CD274 and PDCD1LG2 expression were associated with various immunotherapeutic biomarkers, including mismatch repair (MMR), tumor mutation burden (TMB), microsatellite instability (MSI), and DNA methylation. ^38^

According to Shi’s published work in Frontiers in Oncology, The article reports two cases of early-stage non-small cell lung cancer (NSCLC) displaying full pathological responses to extended neoadjuvant alectinib. The patients had stage IIB (cT3N0M0) EML4-ALK lung adenocarcinoma. The study concludes that administering alectinib as a neoadjuvant treatment for non-small cell lung cancer (NSCLC) is viable; however, comprehensive clinical trials are necessary in the future to establish the treatment protocol and assess the efficacy of neoadjuvant alectinib. Lei’s published work in Spandidos Publications, The article discusses the effectiveness of a novel class of targeted medications in individuals with advanced non-small cell lung cancer (NSCLC) showing ALK rearrangement, who experienced progression after undergoing EML4-ALK targeted therapy. A phase II study on this subject was conducted by the American Society of Clinical Oncology. ^39–42^

According to Mazzaschi’s published work in Springer, the immune properties of STK11 and its role in predicting outcomes versus its role in forecasting outcomes for immunotherapy in non-small cell lung cancer. The conclusion drawn in the article is that STK11 mutations have been universally recognized as indicators of resistance to immunotherapy (IT). Following an extensive review of the literature, it is evident that STK11 mutations influence the prognosis of non-small cell lung cancer (NSCLC) patients undergoing regimens that include immune checkpoint inhibitors (ICIs), signifying notable biological and clinical importance. However, while awaiting conclusive and robust data, there is relatively limited evidence supporting the presumed adverse predictive value of STK11 inactivation concerning immunotherapy. The diverse alterations observed in tumor cells, stroma, and the tumor immune microenvironment (TIME) in STK11 mutant lung cancer, particularly within the molecular subgroup of KRAS co-mutation, can be attributed to the physiological regulation of numerous cellular pathways by STK11. Current information technology (IT) methods in non-small cell lung cancer (NSCLC), predominantly involving anti-PD-1/PD-L1 inhibitors, show limited potential when STK11 is deactivated. Implementing perceptive strategies to manipulate the tumor immune microenvironment (TIME), irrespective of STK11 status or with a specific focus on STK11-mutated cases, is anticipated to offer viable therapeutic choices for clinical application. ^43–47^

FAM83A-AS1 is an antisense transcript of FAM83A gene that has been identified to have a cancer-promoting function in lung adenocarcinoma (LUAD). In G. Wang’s study, it was shown that FAM83A-AS1 was overexpressed in LUAD and involved in the malignant properties of LUAD cells. W. Wang’s study suggested that the progression of lung adenocarcinoma (LUAD) is facilitated by the augmentation of pre-mRNA stability in FAM83A, driven by FAM83A-AS1. However, the exact mechanism of how FAM83A-AS1 acts as a potential target for LUAD is still unclear. ^48–51^

According to Liu’s published work in Frontiers in Oncology, Co-occurring mutations in TP53 within advanced non-small cell lung cancer (NSCLC) characterized by EGFR mutations hold both prognostic and therapeutic importance. Individuals with concurrent mutations in EGFR and TP53 exhibit limited efficacy in responding to EGFR-TKIs, and the prognostic and predictive implications of EGFR/TP53 co-mutation in patients with non-small cell lung cancer (NSCLC) are still a matter of debate. The review further explores the most suitable treatment protocols to effectively extend the survival duration of patients with non-small cell lung cancer (NSCLC) possessing this co-mutation. ^52–55^

According to yan’s published work in Frontiers in Oncology, BRAF mutations commonly occur in non-smokers, women, and aggressive histological types of Non-small cell lung cancer (NSCLC) constitutes 1%-2% of adenocarcinomas. Traditional chemotherapy presents limited efficacy in patients with non-small cell lung cancer (NSCLC) harboring mutations in the BRAF gene. However, the advent of targeted therapy and immune checkpoint inhibitors (ICIs) have greatly altered the treatment pattern of NSCLC. The prevailing and recommended standard treatment for advanced non-small cell lung cancer (NSCLC) featuring BRAF mutations involves the use of targeted therapy specifically designed for BRAF. However, intrinsic or extrinsic resistance mechanisms to tyrosine kinase inhibitors (TKIs) targeting BRAF can emerge in patients. Hence, there are still some problems facing us regarding BRAF-mutated NSCLC. The review further explores the BRAF mutation types, the diagnostic challenges that BRAF mutations present, the strategies to treatment for BRAF-mutated NSCLC, and resistance mechanisms of BRAF-targeted therapy. ^56–58^

Wang’s published work in Frontiers in Oncology, a novel intergenic MIR4299/MIR8070-RET fusion with RET amplification was identified in a lung adenocarcinoma patient Employing next-generation sequencing (NGS). Subsequently, the individual received pralsetinib, and after 3 weeks of therapy, the patient had a partial response. At the time of reporting, the patient was on continuous pralsetinib.^37,59–62^

According to Liu’s published work in the Journal of Oncology, FAM83A has a crucial and fundamental role in the proliferation, advancement, and apoptosis processes in various malignant tumors, including lung adenocarcinoma (LUAD). The objective of the research was to identify the expression pattern of FAM83A in lung adenocarcinoma (LUAD) and examine its association with cancer prognosis and patient survival. Bioinformatics analysis, immunohistochemistry, and Western blotting were employed to investigate and identify the expression of FAM83A in lung adenocarcinoma (LUAD) cells. The investigation focused on understanding the role of FAM83A in proliferation and migration. The association between FAM83A expression and survival rate was evaluated using Kaplan-Meier analysis and Cox regression. The expression of FAM83A was heightened in lung adenocarcinoma (LUAD) tissues and was associated with a shorter overall survival. A notable rise in FAM83A protein levels was evident in the LUAD tissue. In contrast to individuals with early-stage tumors (stage I-II), those with advanced-stage tumors (stage III-IV) exhibited notably elevated levels of FAM83A expression. Reducing the expression of FAM83A resulted in decreased cell proliferation, reduced migration capability, and diminished epithelial-mesenchymal transition (EMT) in the lung cancer cell lines. Elevated expression of FAM83A correlated with early lymph node metastasis and unfavorable overall survival in individuals with lung adenocarcinoma (LUAD). FAM83A could have a crucial role in advancing the progression of lung adenocarcinoma (LUAD) and may therefore function as a new prognostic indicator in LUAD. ^25,63,64^

Liu’s published work in Frontiers in Oncology, RBM10 is among the commonly mutated genes in lung adenocarcinoma (LUAD). Previous studies have confirmed that RBM10 could suppress the disease progression and cell proliferation in LUAD, but its loss-of-function mutations are more frequent in early-stage disease and decrease with the advancement of the clinical stage. However, The absence of RBM10 is linked to heightened immune activity in lung adenocarcinoma (LUAD). tumors with RBM10 mutations displayed higher TMB, and LUADs with RBM10 deficiency also showed higher HLA expression levels, including many HLA class I and II molecules. Additionally, many immune cells, including myeloid dendritic cells, macrophages, neutrophils, and CD8+T cells, showed higher infiltration levels in LUADs with RBM10 deficiency. The findings imply a novel role of RBM10 in the progression of LUAD. ^65–68^

According to the National Comprehensive Cancer Network (NCCN) Clinical Practice Guidelines in Oncology, Mutations in ERBB2 have been documented in 2-3% of individuals diagnosed with advanced lung adenocarcinoma. These mutations are transforming in lung cancer models and result in kinase activation, conferring some in-vitro sensitivity to trastuzumab. Tian’s published work in the Anti-Cancer Drugs journal suggests that ado-trastuzumab emtansine could be effective in treating lung adenocarcinoma with ERBB2 mutations. Mutations in the ERBB2 gene have been observed in 2-3% of individuals diagnosed with advanced lung adenocarcinoma, and these mutations are transforming in lung cancer models and result in kinase activation, conferring some in-vitro sensitivity to trastuzumab. Ettinger’s published work in Frontiers in Oncology reports that ERBB2 ΔEx16 mutations are present in 3% to 4% of individuals with adenocarcinoma NSCLC and 1% to 2% of those with other NSCLC histologies. high-level MET amplification is another potential target for lung adenocarcinoma.^69–72^

Xu’s published work in the Frontiers in Oncology journal, Targeting MET amplification holds promise as a viable approach for addressing lung adenocarcinoma. MET amplification is detected in 3% to 5% of individuals diagnosed with lung adenocarcinoma and is associated with poor prognosis. MET-directed therapies might emerge as a fresh standard of medical practice to tackle MET dysregulation in patients diagnosed with advanced or metastatic non-small cell lung cancer (NSCLC). The research proposes that MET inhibitors, which demonstrated a favorable safety record in the present investigation, could be successful in addressing non-small cell lung cancer (NSCLC) characterized by MET dysregulation. Specifically, tepotinib, savolitinib, and capmatinib emerge as potential effective treatments.^73,74^

Xie’s published work in Frontiers in Endocrinology that discusses the potential role of CDH1 as an oncogene in lung adenocarcinoma (LUAD). Li’s published work in Disease Markers that examines the expression patterns and prognostic significance of members within the CDH family in lung adenocarcinoma (LUAD). The article concludes that CDH1 stands out as one of the genes with the highest frequency of mutations in lung adenocarcinoma (LUAD) and that It might contribute to the growth and advancement of lung adenocarcinoma (LUAD). CDH1 might have a function in the development and advancement of LUAD and that it may be a potential target for LUAD treatment. ^75–77^

PIK3CA gene mutation has been present in a diverse range of human tumors, including lung adenocarcinoma. According to Wang’s systematic review and meta-analysis article, PIK3CA mutation may affect lymph node metastasis and serve as a promising predictive element in patients with non-small-cell lung cancer (NSCLC). However, Additional meticulously planned prospective studies are required to authenticate the abovementioned findings. Regarding the consideration of PIK3CA as a viable target for lung adenocarcinoma, Fusco’s published work in Frontiers in Oncology reported that PIK3CA mutations could be a potential target for hormone receptor-positive breast cancer. However, only pre-clinical studies have so far demonstrated the sensitivity of this mutation to PI3K inhibitors. Zhang’s published work in Dovepress found that mutations in PIK3CA were linked to a reduced duration of progression-free survival in lung adenocarcinoma patients who underwent curative resection. ^78–81^

Moksud’s published work in Journal of Cancer Research and Clinical Oncology that explored the connection between single nucleotide polymorphisms (SNPs) in the PDCD1, CD274, and HAVCR2 genes and the likelihood and consequences of non-small cell lung cancer (NSCLC). subtypes: squamous cell lung cancer (LUSC) and lung adenocarcinoma (LUAD). Frequent inherited variations in the PDCD1, CD274, and HAVCR2 genes distinctly influence the susceptibility and prognosis of adenocarcinoma and squamous cell carcinoma. ^82,83^

Liu’s published work in Frontiers that discusses the role of GNPNAT1 in lung adenocarcinoma (LUAD). GNPNAT1 is a key enzyme in the hexosamine biosynthetic pathway (HBP), which promotes proliferation in some tumors. GNPNAT1 is upregulated in LUAD in contrast to normal tissues, and its overexpression is linked to patients’ clinical phase, size of the tumor, and lymphatic metastasis status. Patients with upregulated GNPNAT1 had a relatively poor prognosis. Furthermore, GNPNAT1 overexpression was correlated with DNA copy amplification, low DNA methylation, and downregulation of hsa-miR-30d-3p. GNPNAT1 expression was linked to B cells, CD4 + T cells, and dendritic cells. Zheng’s article on Europe PMC also mentions GNPNAT1’s role in LUAD. GNPNAT1 is a critical enzyme in the biosynthesis of uridine diphosphate-N-acetylglucosamine. It has many important functions, such as protein binding, monosaccharide binding, and embryonic development and growth. Nevertheless, the function of GNPNAT1 in lung adenocarcinoma (LUAD) is not clearly understood. ^84,85^

Wang’s article on Frontiers that discusses the function of TMPO-AS1 in lung adenocarcinoma (LUAD). TMPO-AS1 is a long non-coding RNA that binds to let-7c-5p and upregulates STRIP2 expression. The article also states that Elevated expression of TMPO-AS1 is linked to an unfavorable prognosis in patients with lung adenocarcinoma (LUAD). ^86–88^

Ren’s article on Cancer Cell International that discusses the function of LINC01116 in lung adenocarcinoma (LUAD). LINC01116, a lengthy non-coding RNA induced by EGR1, promotes the oncogenic characteristics of LUAD by interacting with the miR-744-5p/CDCA4 axis. LINC01116 was expressed at an abnormally high level in LUAD, which was induced by transcription activator EGR1. LINC01116 depletion restrained cell growth, movement and infiltration, yet facilitated apoptosis of Cells of lung adenocarcinoma (LUAD). MiR-744-5p could bind to LINC01116. MiR-744-5p inhibitor reversed the inhibitory effects of silencing LINC01116 on LUAD malignant behaviors. In addition, cell division cycle-associated protein 4 (CDCA4) shared binding sites with miR-744-5p. Silencing LINC01116 elicited decline in CDCA4 mRNA and protein levels. Moreover, CDCA4 up-regulation could counteract the biological effects of LINC01116 knockdown on LUAD cells. ^89–92^

Xu’s published work in Cancer Cell International that explores the biological role and the underlying mechanism of METTL3, the primary catalyst of m6A, in the progression of lung adenocarcinoma (LUAD). METTL3 facilitates the growth of LUAD tumors and hinders ferroptosis by stabilizing m6A modification in SLC7A11. The study also identified Solute carrier 7A11 (SLC7A11), a component of system Xc –, is identified as a direct target of METTL3 through mRNA-seq and MeRIP-seq analyses. The m6A modification facilitated by METTL3 is implicated in the stabilization of SLC7A11 mRNA, enhancing its translation. This process contributes to the promotion of LUAD cell proliferation and the suppression of cell ferroptosis, a recently discovered form of programmed cell death. The conclusion is METTL3 shows potential as an innovative focal point in the diagnosis and treatment of LUAD. ^93–95^

RNA guanine-7 methyltransferase (RNMT) is one of the main regulators of N7-methylguanosine, and the deregulation of RNMT correlated with tumor development and immune metabolism. Huang’s study conducted an integrative pan-cancer analysis of RNMT and found that RNMT represents a possible target for prognosis and therapy in cases of lung adenocarcinoma. the prognosis of patients with lung adenocarcinoma exhibited a significant association with the expression of RNMT. However, the specific function of RNMT in pan-cancer remains unclear. ^96^

Tong’s published work in Frontiers in Genetics that identifies an autophagy-related 12-lncRNA signature and evaluates NFYC-AS1 as a pro-cancer factor in lung adenocarcinoma. The study aims to develop an autophagy-related lncRNA-based risk signature and Nomogram designed to forecast the overall survival (OS) of patients with lung adenocarcinoma (LUAD) and investigate the possible meaning of screened factors. The researchers screened differentially expressed lncRNAs and autophagy genes Comparing samples from the TCGA LUAD dataset, distinctions were observed between normal and lung adenocarcinoma (LUAD) tumor samples. They performed Cox regression analyses, both univariate and multivariate, were conducted to construct the lncRNA-based risk characteristic and nomogram incorporating clinical information. Then, the accuracy and sensitivity were confirmed by the AUC of ROC curves in both training and validation cohorts. qPCR, immunoblot, shRNA, and ectopic expression were used to verify the positive regulation of NFYC-AS1 on BIRC6. CCK-8, immunofluorescence, and Flow cytometry was employed to verify the impact of NFYC-AS1 on cellular proliferation, autophagy, and apoptosis via BIRC6. The conclusion is a 12-lncRNA signature may function as a predictive marker for LUAD patients, and NFYC-AS1 along with BIRC6 may function as carcinogenic factors in a combinatorial manner. ^97,98^

Zavitsanou’s published work in bioRxiv that suggests Mutation in KEAP1 within lung adenocarcinoma encourages evasion of the immune system and resistance to immunotherapy. The study establishes that tumor-intrinsic KEAP1 mutations contribute to immune evasion through suppression of dendritic cell and T cell responses, explaining the observed resistance to immunotherapy of KEAP1 mutant tumors. These results highlight the importance of stratifying patients based on KEAP1 status and open avenues for innovative therapeutic strategies. ^99,100^

LINC00857, identified as a long non-coding RNA (lncRNA), exhibits aberrant expression in lung adenocarcinomas (LUAD) and involved in cell cycle regulation of lung cancer. LINC00857 controls the advancement, apoptosis, and glycolysis of lung adenocarcinoma (LUAD) by interacting with the miR-1179/SPAG5 axis. LINC00857 knockdown led to the repression of cell proliferation and glycolysis, while the apoptosis was elevated in LUAD cell lines. Furthermore, LINC00857 promoted cell growth and glycolysis and repressed apoptosis via sponging miR-1179 and further regulating sperm-associated antigen 5 (SPAG5) expression in LUAD cultured cell models. These discoveries suggest that LINC00857 could be a potential target for clinical treatment of LUAD patients. ^101–103^

KIF5B is a kinesin family member that plays a pivotal role in intracellular transport and mitosis. Zhu’s study has shown that KIF5B plays a role in the progression of lung adenocarcinoma (LUAD). A new chimeric fusion transcript of KIF5B and the oncogenic variant, KIF5B-RET, associated with the RET gene, was detected in 1% to 2% of lung adenocarcinomas (LUADs). The fusion gene involving KIF5B and RET can induce the abnormal growth and maturation of cancer cells through the constitutional overexpression and activation of the RET proto-oncogene, ultimately leading to LADCs. Individuals with non-small cell lung cancer (NSCLC) carrying the KIF5B-RET fusion gene can become sensitive to cytotoxic chemotherapies, including pemetrexed-based regimens, similar to ALK- or ROS1-rearranged lung cancers. Although there are currently no clinically approved RET-selective inhibitors, several multi-targeted drugs with anti-RET activity, including cabozantinib, vandetanib, sunitinib, and Sorafenib have been examined in preclinical models and clinical trials. These findings suggest that targeting KIF5B-RET may be considered as a potential approach for the clinical treatment of patients with lung adenocarcinoma (LUAD). ^104–107^

Li’s article suggests that Biochanin A, a naturally occurring isoflavone, has been demonstrated to exhibit anticancer effects in various tumors, including lung adenocarcinoma. Biochanin A could specifically inhibit the expression of ZEB1, which is associated with both chemoresistance and metastasis. Importantly, Biochanin A chemosensitizes lung adenocarcinoma and inhibits cancer cell metastasis by suppressing ZEB1. At the molecular level, Biochanin A affects the stability of ZEB1 protein through the deubiquitination pathway and thereby influences the progression of lung adenocarcinoma.^108–110^

Li’s article suggests that CCNB2, a protein that belongs to the type B cell cycle family, is upregulated in all subtypes of lung cancer, including lung adenocarcinoma. The study collected 103 high-throughput datasets related to all subtypes of lung cancer and cerebral ischemic stroke with the data of CCNB2 expression. The analysis of standard mean deviation (SMD) and summary receiver operating characteristic (SROC) reflecting expression status demonstrated significant up-regulation of CCNB2 in lung adenocarcinoma (SMD = 1. 40, 95%CI [0. 98–1.83], SROC = 0. 92, 95%CI [0. 89–0. 94]). ^111^

Li’s article suggests that LMO7DN has been suggested as a predictor of lung adenocarcinoma associated with ferroptosis. The research created and confirmed a prognostic model for lung adenocarcinoma using expression profiles of mRNA and lncRNA. The model combined the risk scores of the first two models with clinical variables and included three predictors in total: mRNA risk score, lncRNA risk score, and tumor stage. LMO7DN May serve as a prospective target for the treatment of lung adenocarcinoma. ^112^

According to Liu’s published work in Frontiers in Immunology that investigates the role of ERO1L in shaping the immune-suppressive tumor microenvironment (TIME) in lung adenocarcinoma (LUAD). The study reveals that ERO1L is significantly overexpressed in LUAD in comparison to normal tissue. This overexpression is found to be a result of hypomethylation of the ERO1L promoter. Overexpression of ERO1L results in an immune-suppressive TIME via the recruitment of Cells with immunosuppressive properties, such as regulatory T cells (Tregs), cancer-associated fibroblasts, M2-type macrophages, and Suppressor cells originating from myeloid lineage. Using the Tumor Immune Dysfunction and Exclusion (TIDE) framework, it was identified that patients in the ERO1Lhigh group possessed a significantly lower response rate to immunotherapy in comparison to the ERO1Llow group. Examination of mechanisms indicated that the increased expression of ERO1L was linked to the elevation of JAK-STAT and NF-κB signaling pathways, thus affecting chemokine and cytokine patterns in the TIME. ^113^

Koren’s published work in the Springer journal that investigates the prognostic value of cytokeratin-7 (KRT7) mRNA expression in the complete peripheral blood of individuals with advanced lung adenocarcinoma. KRT7 is strongly expressed in adenocarcinomas specifically, rather than in other forms of non-small-cell lung carcinoma subtypes. The combination of KRT7-positive/KRT20-negative immunostaining has been shown to be useful in discriminating primary from secondary lung adenocarcinomas. ^114^

Cheng’s published work in BMC Cancer that describes TBX5-AS1 serves as an enhancer RNA and represents a potential new prognostic biomarker for lung adenocarcinoma. TBX5-AS1 functions as the primary enhancer RNA (eRNA), with T-box transcription factor 5 (TBX5) identified as its regulatory target. According to KEGG analysis, TBX5-AS1 is likely to play a crucial role through the PI3K/AKT pathway, Ras signaling pathway, etc. Moreover, the findings from quantitative real-time polymerase chain reaction (qRT-PCR) and the Human Protein Atlas (HPA) database revealed a notable downregulation of TBX5-AS1 and TBX5 in tumor samples when compared to corresponding adjacent pairs. The pan-cancer validation results demonstrated that TBX5-AS1 exhibited a survival association in four types of tumors: adrenocortical carcinoma (ACC), lung adenocarcinoma (LUAD), lung squamous cell carcinoma (LUSC), and uterine corpus endometrial carcinoma (UCEC). ^115,116^

Zhang’s published work in the journal Tumor Biology, the long noncoding RNA SFTA1P, which is derived from a pseudogene, experiences downregulation and exerts inhibitory effects on cell migration and invasion in lung adenocarcinoma. Reduced expression of SFTA1P is specifically associated with a shorter survival time in patients with lung adenocarcinoma, with no correlation observed in the survival of patients with lung squamous cell carcinoma. The authors of the study performed gain-of-function studies including growth curves, migration, performing invasion experiments and in vivo investigations to verify the tumor suppressor role of SFTA1P in non–small cell lung cancer. Finally, the potential underlying pathways involved in SFTA1P were investigated by analyzing the SFTA1P-correlated genes Within the lung adenocarcinoma dataset of The Cancer Genome Atlas and normal tissues RNA sequencing data. The conclusion is pseudogene-derived long noncoding RNA SFTA1P exerts the tumor suppressor functions in human lung adenocarcinoma and Could potentially be a novel focus for the diagnosis and treatment of lung adenocarcinoma. ^117,118^

BCL2 is a proto-oncogene responsible for encoding a protein with a molecular weight of 25 kDa, mainly localized to the inner mitochondrial membrane, endoplasmic reticulum, and nuclear envelope. It prevents cells from undergoing apoptosis and overexpression of BCL2 increases the lifespan of B cells, which may maintain memory B cells, plasma cells, and neurons by prolonging lifespan without cell division. BCL2 may participate in ion channel formation and alteration of membrane permeability, necessary for the initiation of apoptosis. Inhibition of BCL2 by Hedyotis diffusa injection (HDI) can induce ferroptosis in lung adenocarcinoma cells via BCL2 inhibition to promote Bax regulation of VDAC2/3The EGFR pathway modulates the Bax/Bcl-2 cascade in non-small cell lung carcinoma (NSCLC). Targets including BCL2 were highly expressed in all tumors, and MCL1 was significantly increased in breast cancer and lung adenocarcinoma^119–121^

Yalimaimaiti’s published work in BMC Bioinformatics, This indicates that LINC00592 plays a role in controlling copper metabolism and cuproptosis, a form of cell demise triggered by the buildup of copper. Zhang’s published work in Frontiers suggests that LINC00592 shows promise as a biomarker for forecasting prognosis and appraising the tumor immune microenvironment in cases of lung adenocarcinoma. The Frontiers article introduces a prognostic signature for lung adenocarcinoma, derived from long non-coding RNAs (lncRNAs) associated with cuproptosis, which includes LINC00592. Thirteen long non-coding RNAs (lncRNAs) associated with cuproptosis could serve as molecular biomarkers with clinical relevance for predicting the prognosis of lung adenocarcinoma (LUAD).^122–125^

Tang’s article discusses a study that aimed to identify effective prognostic predictors for lung adenocarcinoma (LUAD) patients. The study used Long non-coding RNAs associated with cuproptosis (CRlncRNAs) to build a risk model that categorized individuals into groups with elevated and diminished risks. In accordance with the CRlncRNAs in the model, consensus clustering analysis was used to classify LUAD patients into different subtypes. Next, the study explored the differences in overall survival (OS), the tumor immune microenvironment (TIME), and the mutation landscape between various risk categories and molecular subcategories. Finally, the functions of LINC00968 were verified through in vitro experiments. The study found that LINC00968 knockdown significantly reduced LUAD cell proliferation, invasion, and migration ability. ^126–128^

Zhuo’s article discusses a study that aimed to investigate the role of tumor endothelial cell-derived cadherin-2 (CDH2) in lung adenocarcinoma (LUAD). The study used immunohistochemistry to detect CDH2 expression in LUAD tissues and Surrounding healthy tissues. The study discovered that CDH2 the manifestation was significantly higher in LUAD biological tissues than in adjacent normal tissues. The study also found that CDH2 expression was positively correlated with Density of microvessels (MVD) and the expression of vascular endothelial growth factor (VEGF). The conclusion is CDH2 promotes angiogenesis and has prognostic significance for LUAD. ^129^

Morgenstern’s article discusses a study that aimed to understand the role and real impact of LINC00941 in tissue homeostasis and cancer development. LINC00941 was highly upregulated in different forms of cancer, encompassing lung cancer. adenocarcinoma (LUAD), and influenced cell proliferation and metastasis. The study demonstrated multiple potential modes of action of LINC00941 influencing the functionality of various cancer cell types. Correspondingly, LINC00941 was proposed to be involved in regulation of mRNA transcription and modulation of protein stability, respectively. In addition, several experimental approaches suggest a function of LINC00941 as competitive endogenous RNA, thus acting in a post-transcriptional regulatory fashion. ^130,131^

Ruan’s article discusses a study that aimed to construct consensus clusters based on multi-omics data and multiple algorithms to identify specific molecular characteristics and facilitate the use of precision medicine on patients. The study used gene expression, DNA methylation, gene mutations, copy number variation data, and clinical data of LUAD patients for clustering. Consensus clusters were obtained using a consensus ensemble of five multi-omics integrative algorithms. Four molecular subtypes were identified. The CHIAP2 gene was differentially expressed between the four subtypes. The conclusion is CHIAP2 is a potential target for lung adenocarcinoma. ^132^

Lu’s published work in the Journal of Oncology discovered a prognostic biomarker for lung adenocarcinoma, characterized by a ferroptosis-related long non-coding RNA (lncRNA) signature. Shen’s published work in Europe PMC found that WWC2-AS2 is one of the four lncRNAs with reduced expression linked to the prognosis of lung adenocarcinoma in humans. Cui’s published work in Hindawi Journal of Oncology identified WWC2-AS2 as one of the lncRNAs involved in the construction of a Predictive long non-coding RNA (lncRNA) pattern in lung adenocarcinoma.^133–136^

### Lung Adenocarcinoma treatment

#### Targeted Therapy

Gefitinib is a tyrosine kinase inhibitor of small molecular size, specifically designed to target abnormal proteins within cancerous cells It is employed as the initial treatment to address non-small cell lung carcinoma (NSCLC) that meets certain genetic mutation criteria. Gefitinib hinders the internal phosphorylation of various tyrosine kinases linked to transmembrane receptors on the cell surface, including those associated with the epidermal growth factor receptor (EGFR-TK). EGFR is often shown to be overexpressed in certain human carcinoma cells, such as lung and breast cancer cells. Elevated expression results in increased activation of the anti-apoptotic Ras signal transduction pathways, subsequently resulting in increased survival of cancer cells and uncontrolled cell proliferation. Cui’s article published in Investigational New Drugs journal reports that Gefitinib could be a viable treatment choice for individuals with pulmonary enteric adenocarcinoma (PEAC) carrying a combined EGFR L858R and A871G mutation. A postoperative PEAC patient with EGFR L858R+A871G combined mutation underwent treatment with gefitinib and achieved an impressive 5-year progression-free survival (PFS). ^137–144^

Osimertinib, a small molecule tyrosine kinase inhibitor, specifically addresses the aberrant proteins present in cancerous cells. It is employed as the initial treatment in the management of non-small cell lung carcinoma (NSCLC) that meets certain genetic mutation criteria. Osimertinib obstructs the internal phosphorylation of various tyrosine kinases linked to transmembrane receptors on the cell surface, including those associated with the epidermal growth factor receptor (EGFR-TK). Gen’s published work in BMC Cancer journal reports that Osimertinib has been extensively employed as the primary therapeutic approach for individuals with metastatic non-small cell lung cancer (NSCLC) harboring EGFR mutations. While Osimertinib exhibited efficacy in managing central nervous system metastases, its effectiveness against distant organs such as bone and liver remains uncertain. Osimertinib yielded superior clinical advantages compared to first- and second-generation EGFR-TKIs in individuals with EGFR-mutant non-small cell lung cancer (NSCLC), especially those with brain or bone metastases and exon 19 deletion. Nevertheless, its effectiveness against liver metastasis was significantly reduced. There is a requirement for new therapeutic advancements for patients with EGFR-mutant NSCLC and liver metastases. ^145–152^

Crizotinib, a small molecule tyrosine kinase inhibitor, specifically addresses aberrant proteins in cancerous cells. Employed as an initial treatment, it is utilized in the management of non-small cell lung carcinoma (NSCLC) that meets certain genetic mutation criteria. Crizotinib hinders the internal phosphorylation of various tyrosine kinases linked to transmembrane receptors on the cell surface, including those related to anaplastic lymphoma kinase (ALK) and ROS1. Wu’s published work in Anti-Cancer Drugs journal reports that In the management of stage IV lung adenocarcinoma, initial crizotinib therapy proves effective for a newly identified SEC31A-ALK fusion in a patient. A computed tomography (CT) scan showed numerous scattered areas of increased density in both lungs. The individual received crizotinib and experienced a partial positive outcome. ^153–158^

Afatinib is a small molecule drug that is used as a targeted therapy for Lung adenocarcinoma. According to Chen’s article, Afatinib has demonstrated positive response rates and extended progression-free survival in individuals with lung cancer possessing EGFR mutations when compared to the conventional platinum-based chemotherapy. Commencing Afatinib at a reduced initial dosage (30mg) can be used to reduce the incidence of moderate and severe adverse drug reactions (ADRs). Nie’s article reports an instance of late-stage lung adenocarcinoma carrying a new NPTN-NRG1 fusion, which achieved durable response to Afatinib. The report supports that Afatinib can provide potential benefit for NRG1 fusion patients, and RNA-based next-generation sequencing (NGS) is A precise and economical option strategy for fusion detection and isoform identification. Afatinib works by irreversibly binding to The tyrosine kinase region within the epidermal growth factor receptor (EGFR). This binding inhibits the Signaling pathways further downstream that facilitate cellular growth, angiogenesis, and metastasis. ^159–167^

Erlotinib is a small molecule drug that is used as a targeted therapy for Lung adenocarcinoma. According to Majeed’s article, Erlotinib has demonstrated positive response rates and extended progression-free survival in lung cancer patients possessing EGFR mutations when compared to the conventional platinum-based chemotherapy. Erlotinib may be employed as the initial treatment for individuals with EGFR mutations who have advanced non-small cell lung cancer (NSCLC). Araghi’s article reports that Erlotinib can be used as a maintenance therapy for Individuals with advanced non-small cell lung cancer (NSCLC) who show no signs of progression following four rounds of platinum-based chemotherapy. Erlotinib works by reversibly attaching to the tyrosine kinase region of the epidermal growth factor receptor (EGFR). This binding inhibits the Signaling pathways further downstream that stimulate the growth of cells, angiogenesis, and metastasis.^168–170^

Icotinib is a targeted therapy employed as a small molecule drug for the treatment of lung adenocarcinoma. According to Tan’s article, Icotinib has shown promising results as targeted therapy for non-small cell lung cancer (NSCLC) individuals possessing EGFR mutations. The study reports that Icotinib exhibits a favorable rate of response and a more extended period of progression-free survival when contrasted with the usual platinum-based chemotherapy. Jin’s article reports that For individuals with lung adenocarcinoma carrying the combined EGFR L858R/A871G mutation, Icotinib may serve as a viable and effective treatment choice. Icotinib functions by specifically attaching to the tyrosine kinase domain of the epidermal growth factor receptor (EGFR) at its ATP-binding site. This binding inhibits Signaling pathways further downstream that encourage the proliferation of cells, angiogenesis, and metastasis. ^168–172^

Alectinib is a kinase inhibitor that selectively hinders the function of the anaplastic lymphoma kinase (ALK) tyrosine kinase, specifically employed for addressing non-small cell lung cancer (NSCLC) characterized by the presence of the ALK-EML4 (echinoderm microtubule-associated protein-like 4) fusion protein, which triggers the growth of NSCLC cells. Shi’s article is a systematic review of case reports, it suggests that neoadjuvant therapy with alectinib resulted in complete pathological response in a pair of instances early-stage NSCLC with EML4-ALK rearrangement. Alectinib is an orally administered small molecule medication and is classified as a targeted therapy. Alectinib is a small molecule medication that specifically hinders the function of ALK tyrosine kinase. It is used as a targeted therapy for Non-small cell lung cancer (NSCLC) that exhibits the ALK-EML4 fusion protein. Neoadjuvant therapy with alectinib could be effective in achieving complete pathological response in early-stage NSCLC with EML4-ALK rearrangement. ^173–177^

Ceritinib is a kinase inhibitor in the form of a small molecule, employed for the treatment of metastatic non-small cell lung cancer (NSCLC) with positive anaplastic lymphoma kinase (ALK) in individuals with inadequate clinical response or intolerance to crizotinib. It is a type of ALK inhibitor belonging to the second generation that was developed to overcome crizotinib resistance. Ceritinib achieves its therapeutic impact by preventing the autophosphorylation of ALK, inhibiting ALK-induced phosphorylation of the downstream signaling protein STAT3, and suppressing the proliferation of cancer cells dependent on ALK. Ceritinib is employed to treat metastatic non-small cell lung cancer (NSCLC) in adults with anaplastic lymphoma kinase (ALK) positivity, specifically after the ineffectiveness (due to resistance or intolerance) of previous crizotinib therapy. The mechanism of action of Ceritinib is that It suppresses Anaplastic Lymphoma Kinase (ALK), also recognized as the ALK tyrosine kinase receptor or CD246 (cluster of differentiation 246), an enzyme encoded by the ALK gene in humans. ^178,179^

Anlotinib serves as a targeted therapy medication for non-small cell lung cancer (NSCLC), functioning as a small molecule tyrosine kinase inhibitor. Anlotinib works by inhibiting the functioning of multiple receptor tyrosine kinases, such as the vascular endothelial growth factor receptor (VEGFR), fibroblast growth factor receptor (FGFR), and platelet-derived growth factor receptor (PDGFR), and c-Kit. Xiong’s published work in Frontiers in Oncology reported that Anlotinib demonstrates favorable tolerance and effectiveness in individuals with advanced non-small cell lung cancer (NSCLC). Anlotinib was administered to 80 patients with non-small cell lung cancer (NSCLC) in stage IIIB-IV diagnosed between November 2018 and February 2020. The median duration of progression-free survival (PFS) was 4.3 months, and The prevalent adverse events included fatigue, reduced hemoglobin levels, high blood pressure, hand-foot syndrome, oral mucositis, and hoarseness. The median progression-free survival (PFS) for individuals receiving a combination of anlotinib and immunotherapy was marginally extended compared to those receiving anlotinib alone (4.2 vs. 3.1 months). Nevertheless, this disparity did not reach statistical significance. Xu’s published work in World Journal of Surgical Oncology presented a case of lung adenocarcinoma with HER2 p. Asp769Tyr mutation that was successfully treated with a combination of afatinib and anlotinib. ^163,180–182^ Lorlatinib is a targeted therapy drug employed for the treatment of non-small cell lung cancer (NSCLC), acting as a small molecule tyrosine kinase inhibitor. Lorlatinib functions by suppressing the activity of anaplastic lymphoma kinase (ALK), c-ros oncogene 1 (ROS1), and proto-oncogene tyrosine-protein kinase (RET). Lee’s published work in Anti-Cancer Drugs reported that Lorlatinib demonstrates good tolerability and effectiveness in the treatment of patients with advanced ALK-positive non-small cell lung cancer (NSCLC). lorlatinib was administered to 22 ALK-positive individuals who had undergone at least one second-generation Inhibitor of ALK. The rate of positive responses was 35.7%, and the overall disease control rate reached 64.3%. The median duration of progression-free survival (PFS) was 6.2 months. Patients receiving only second-generation ALK inhibitors showed Progression-free survival surpasses that of individuals treated with both crizotinib and second-generation ALK inhibitors. ^183–186^

Pralsetinib is a kinase inhibitor that primarily targets wild-type RET (IC50s between 0. 14 and 0. 43 nmol/L) as well as kinase-activating RET mutations (V804L, V804M, V804E, M918T, C634W) and fusions (CCDC6-RET and KIF5B-RET) in in vitro and/or in vivo studies. On September 4, 2020, the FDA provided expedited approval for pralsetinib in the treatment of metastatic non-small cell lung cancer with positive RET fusion. Pralsetinib is a once-daily, oral precision therapy, which selectively targets cancer-causing RET alterations. Pralsetinib is a small-molecule medication that hampers the function of the RET protein, playing a role in cell growth and viability. Pralsetinib is used as a targeted therapy for Individuals diagnosed with non-small cell lung cancer (NSCLC) characterized by the presence of metastatic RET fusion. ^187^

Brigatinib is a small molecule drug that hampers the function of the RET protein, which is participating in the expansion and cell survival. Brigatinib is used as a targeted therapy for Individuals diagnosed with non-small cell lung cancer (NSCLC) characterized by the presence of metastatic RET fusion. Dagogo-jack’s article reports that Brigatinib serves as a reversible dual inhibitor targeting both anaplastic lymphoma kinase (ALK) and epidermal growth factor receptor (EGFR). Notably, it demonstrates specificity for mutant EGFR variants over the wild-type. Additionally, the compound displays selectivity against nine distinct crizotinib-resistant mutants associated with the EML4-ALK fusion gene, a crucial factor in the transformation of vulnerable lung parenchyma. On September 4, 2020, the FDA provided expedited approval for brigatinib in the treatment of metastatic non-small cell lung cancer with positive RET fusion. Brigatinib is a once-daily, oral precision therapy, which selectively targets cancer-causing RET alterations. ^188,189^

Ensartinib is a small molecule drug that is used as a targeted therapy for Individuals diagnosed with non-small cell lung cancer (NSCLC) characterized by rearrangement in the anaplastic lymphoma kinase (ALK) gene. It is a strong suppressor of ALK and most of its alternations (fusions, mutants). Ensartinib has demonstrated excellent effectiveness in individuals with ALK-positive non-small cell lung cancer (NSCLC) who experienced advancement despite prior treatment with crizotinib in the Phase 1/2 trial. Ensartinib has also been studied as neoadjuvant therapy designed to treat individuals diagnosed with ALK-positive conditions, resectable for stage II to IIIB(N2) NSCLC. Ensartinib acts by inhibiting of ALK whose aberration causes cancers. ^190–194^

Aumolertinib is a small molecule drug that is used as a targeted therapy for non-small cell lung cancer (NSCLC) patients with Increasing sensitivity epidermal growth factor receptor (EGFR) mutations. This EGFR tyrosine kinase inhibitor (EGFR-TKI) belongs to the third generation and demonstrates selectivity for mutant EGFR compared to wild-type EGFR. Aumolertinib has been developed to address advanced non-small cell lung cancer (NSCLC) with positive EGFR mutations. In the phase 3 AENEAS trial conducted among Chinese patients, aumolertinib demonstrated a significant extension of progression-free survival (PFS) and duration of response (DoR) as a first-line treatment compared to gefitinib in individuals with advanced EGFR mutation-positive NSCLC. In the phase 1/2 APOLLO trial, aumolertinib exhibited favorable clinical activity (as indicated by objective response rate, PFS, DoR, and overall survival) in Chinese patients with locally advanced or metastatic NSCLC carrying the EGFR T790M mutation, who had experienced progression following prior EGFR-TKI therapy. Aumolertinib operates by inhibiting the activity of EGFR whose aberration causes cancers.^195–201^

Savolitinib, designed as a small molecule inhibitor targeting the tyrosine kinase of the mesenchymal epithelial transition factor (MET), is mainly intended for treating non-small cell lung cancer (NSCLC) associated with MET mutations. Functioning as a MET tyrosine kinase inhibitor with high specificity, it exhibits promise in treating various cancers, such as NSCLC, breast, head and neck, colorectal, gastric, pancreatic, and other gastrointestinal cancers. Lee’s published work in Cancers, both preclinical and clinical studies have shown effectiveness in lung, kidney, and stomach cancers. Savolitinib, an oral anti-cancer medication, is given at a daily dosage of 600 mg. It serves as a monotherapy for individuals with non-small cell lung cancer (NSCLC) harboring MET mutations and can be combined with epidermal growth factor receptor (EGFR) inhibitors for those who have become resistant. Additionally, positive outcomes have been observed for gastric cancer therapy, especially when administered alongside docetaxel.^202,203^

Tepotinib is a small molecule drug utilized for the treatment of individuals with non-small cell lung cancer (NSCLC) who have MET exon 14 skipping alterations. Tepotinib is a kinase inhibitor specifically targeting MET, including METex14-skipping alterations. Given once daily, tepotinib hinders MET phosphorylation and the ensuing downstream signaling pathways, effectively restraining tumor growth, anchorage-independent growth, and the migration of MET-dependent tumor cells. Albers’s research indicates that tepotinib displays significant, dose-dependent anti-tumor efficacy in models of MET-dependent tumors across different cancer types. Makimoto’s study reported that tepotinib shows an antitumor effect for individuals with non-small cell lung cancer (NSCLC) possessing MET exon 14 skipping mutations. In fact, Tepotinib is presently authorized for managing adult individuals with advanced or metastatic non-small cell lung cancer containing mutations involving exon 14 skipping in the MET gene.^204–207^

Capmatinib is a small-molecule medication employed for the therapy of non-small cell lung cancer (NSCLC) patients exhibiting mutations involving exon 14 skipping in the MET gene. It functions as a kinase inhibitor targeting MET, encompassing METex14-skipping alterations. Given once a day, capmatinib suppresses the heightened activity of c-Met, a receptor tyrosine kinase encoded by the MET proto-oncogene, known to be implicated in the proliferation of various cancers, including non-small cell lung cancer (NSCLC). Capmatinib inhibited the growth of cancer cells resulting from exon 14 skipping at concentrations feasible for clinical use. It also demonstrated anti-tumor effectiveness in murine tumor xenograft models representing human lung tumors with either a mutation leading to METex14 skipping or MET amplification. Capmatinib is presently authorized for treating adults diagnosed with metastatic non-small cell lung cancer that exhibits MET exon 14 skipping alterations.^208–212^

Dacomitinib is a small molecule that inhibits the tyrosine kinases of the epidermal growth factor receptor (EGFR) family, primarily Created for addressing non-small cell lung cancer (NSCLC) characterized by EGFR exon 19 deletion or exon 21 L858R substitution mutations. It is an oral medication taken once daily. dacomitinib has been shown to be effective in treating NSCLC characterized by either EGFR exon 19 deletion or exon 21 L858R substitution mutations. Dacomitinib increased the length of time without disease progression in individuals diagnosed in advanced stages NSCLC.^213,214^

Trametinib is a small molecule drug that is used as a targeted therapy for Lung adenocarcinoma. BRAF and KRAS represent pivotal oncogenes within the RAS/RAF/MEK/MAPK signaling pathway. Concurrent mutations involving both KRAS and BRAF genes have been detected in non-small cell lung cancer (NSCLC), influencing the growth, maturation, and programmed cell death of cancerous cells through activation of the RAS/RAF/MEK/ERK signaling pathway. Currently, therapeutic agents targeting the RAS/RAF/MEK/ERK signaling pathway have been explored in NSCLC patients with BRAF mutations. Approval has been granted for BRAF and MEK inhibitors in treating NSCLC patients. Combining MEK inhibitors with chemotherapy, immune checkpoint inhibitors, epidermal growth factor receptor-tyrosine kinase inhibitors, or BRAF inhibitors holds significant promise for enhancing effectiveness in clinical settings and delaying the onset of resistance to medication. Trametinib, a MEK1/MEK2 inhibitor, was effective in treating KRAS-mutant Late-stage non-small-cell lung cancer (NSCLC). Another study reported a rapid response to Trametinib monotherapy in an individual diagnosed with advanced adenocarcinoma of the lungs, presenting with de novo SND1-BRAF fusion. Trametinib works by inhibiting the activity of MEK1 and MEK2 proteins, which are Participating within the signaling cascade of the RAS/RAF/MEK/ERK pathway. By blocking this pathway, Trametinib can slow down or halt the proliferation of cancerous cells.^194,215–222^

#### Chemotherapy

Cisplatin is a first-line chemotherapy medication employed in the therapy of lung cancer. It is a small molecule drug that is commonly used in ovarian, lung, and breast cancer treatment as a first-line therapeutic and also in combination chemotherapy. The most challenging problems in treating lung cancer with this drug include the development of drug resistance by the cells, low water solubility, and adverse effects on the normal cells. Pavan’s published work in the Journal of Materials Science, nanotechnology-based drug delivery approach has demonstrated encouraging outcomes in increasing the cellular absorption of drugs by the cancer cells with minimal adverse effects. Cisplatin formulations including polymeric nanoparticles, micelles, dendrimers, and liposomes have a greater chance of delivering persistent cisplatin to the growth in reaction to alterations in the tumor’s microenvironment. ^223–227^

Pemetrexed is a small molecule drug that is employed in the therapy of non-squamous non-small cell lung cancer (NSCLC). It is a chemotherapeutic substance that works by inhibiting the enzymes participating in the creation of nucleotides, which are essential for DNA replication and cell division. This results in the demise of cancerous cells. According to Gyawali’s published work in JAMA Oncology, pemetrexed has been shown to provide durable responses among individuals with progressed non-squamous NSCLC. Patil’s published work in Frontiers in Oncology reported that pemetrexed is an established treatment for individuals in an advanced stage non-squamous NSCLC. Targeted therapy constitutes a form of cancer treatment that uses drugs or other substances to identify and attack specific cancer cells without harming normal cells. Pemetrexed is not a targeted therapy drug. Immunotherapy is another type of cancer treatment that helps the immune system fight cancer. Pemetrexed is not an immunotherapy drug either.^167,228–232^

Carboplatin is a chemotherapeutic medication comprising the metallic element platinum. It stops or slows the growth of cancer cells and other rapidly growing cells by damaging their DNA. Carboplatin is authorized for solo use or alongside other medications as part of the therapy for ovarian cancer that is advanced. It is utilized in conjunction with other chemotherapy as first-line treatment. It is used alone as palliative treatment for disease that has recurred (come back) after earlier chemotherapy. Takashima’s published work in 2020 investigated the effectiveness and acceptability of CBDCA (area under the curve 5) in addition gemcitabine (GEM, 1000 mg/m 2) as adjuvant chemotherapy for non-small cell lung cancer (NSCLC) patients who had undergone total removal. The study concluded that CBDCA + GEM was efficient and easily endured as adjuvant chemotherapy, with a manageable toxicity profile. Carboplatin predominantly acts by attaching alkyl groups to the nucleotides, leading to the formation of monoadducts, and DNA fragmenting when repair enzymes attempt to correct the error. 2% of carboplatin’s activity comes from DNA cross-linking from a base on one strand to a base on another, preventing DNA strands from separating. ^233–235^

Platinum doublet chemotherapy is a combination of two chemotherapy drugs, one of which is a platinum-based drug such as carboplatin or cisplatin. Bodor’s published work in 2018 states that Conventional therapy for non-small-cell lung cancer (NSCLC) has typically relied on chemotherapy featuring platinum-based agents. although In the past few years, immunotherapy has become a prominent therapeutic approach option and can result in robust and durable treatment responses in a portion of individuals. Vellanki’s published work in 2021 also mentions that platinum doublet chemotherapy has historically been the typical initial therapeutic approach therapy for individuals who are diagnosed accompanied by metastatic lung adenocarcinoma devoid of a targetable mutation. Platinum-based antineoplastic agents cause crosslinking of DNA as monoadduct, interstrand crosslinks, intrastrand crosslinks or DNA protein crosslinks. ^236–239^

Paclitaxel is a type of chemotherapy drug employed in the management of diverse forms of cancer, including lung adenocarcinoma. It’s a small-molecule medication that disrupts the usual process of microtubule growth.Paclitaxel is one of several cytoskeletal drugs that target tubulin. Paclitaxel is used in conjunction with other drugs as an initial therapy for progressed non-small cell lung cancer (NSCLC). Additionally used as a second-line therapy for NSCLC when other treatments have failed. Paclitaxel has been shown to have the capacity to upregulate articulation of PD-L1 on cancer cells and to promote antitumor immunogenicity, via activation of cytotoxic T lymphocytes, maturation of antigen-presenting cells, depletion of immunosuppressive T cells with regulatory functions and/or expansion of cells with origins in the myeloid lineage that exert suppressive functions. ^240–246^

Docetaxel is a small molecule chemotherapy drug utilized in the treatment of different kinds of cancer, including lung adenocarcinoma. It works by interfering with the normal function of microtubule growth, which is essential for cell division. Docetaxel is a taxane and is administered intravenously. docetaxel has been shown to demonstrate efficacy in addressing lung adenocarcinoma. Docetaxel, when used in combination with other chemotherapy drugs, improved survival rate rates among individuals diagnosed with advanced adenocarcinoma of the lungs. ^247^

#### Immunotherapy

Pembrolizumab is a drug that has shown promise in treating lung adenocarcinoma. according to Song’s article, pembrolizumab has been found to be effective in treating large-cell carcinoma (LCC) and large-cell neuroendocrine carcinoma (LCNEC) of the lung. Pembrolizumab, when combined with chemotherapy, improved the overall rate of response (ORR) and rate of disease control (DCR) of LCC and LCNEC patients. The median duration of progression-free survival (mPFS) was 7. 0 months for LCC and 5. 5 months for LCNEC. Pembrolizumab is a type of immunotherapy drug that works by hindering lymphocytes’ PD-1 receptors, blocking the ligands that would deactivate it and prevent an immune response. It is a monoclonal antibody that attaches to the protein PD-1 present on the exterior of immune cells called T cells. This allows the body’s defense mechanism to target and eliminate cancerous cells, but also blocks a key mechanism preventing the immune system from attacking the body itself. ^248–252^

Nivolumab is a monoclonal antibody that is used as an immunotherapy drug as a treatment-related approach for non-small cell lung cancer (NSCLC). It is a biologic drug that is administered intravenously. Nivolumab works by blocking the T cells’ programmed death-1 (PD-1) receptor, which helps enable the immune system to identify and attack cells affected by cancer. Rosner’s published work in Clinical Cancer Research reported that neoadjuvant nivolumab is secure and viable for resectable NSCLC. the investigation found that two doses of nivolumab (3 mg/kg) were administered for 4 weeks before surgery to 21 individuals diagnosed with non-small cell lung cancer (NSCLC) in Stage I-IIIA. The 5-year recurrence-free survival (RFS) and overall survival (OS) rates were 60% and 80%, respectively. The presence of major pathological responses (MPR) and pre-treatment tumor PD-L1 positivity each trended toward favorable RFS. Dong’s published work in Frontiers in Immunology presented a case of PD-L1-overexpressing lung adenocarcinoma that harbors both EML4-ALK gene reorganization and BRAF mutation. The study found that the individual exhibited a comprehensive response to nivolumab. Johan’s published work in BMC Pulmonary Medicine highlighted the safe and effective use of nivolumab for managing metastatic lung adenocarcinoma that occurred in a patient with sporadic LAM. ^253–255^

Atezolizumab is a type of immunotherapy drug that helps the immune system of the body to track down and fight cancer. It is a monoclonal antibody that works by binding to the protein PD-L1 present on the exterior of some cancer cells, which keeps cancer cells from suppressing the immune system. This allows the immune system to attack and kill the cancer cells. Atezolizumab has been studied As a therapeutic approach for non-small cell lung cancer (NSCLC). Atezolizumab is used alone as a therapy for NSCLC To lower the risk of lung cancer recurrence following the surgical removal of your tumor(s) and you have received platinum-based chemotherapy, and you have stage 2 to stage 3A NSCLC (talk to your healthcare provider about what these stages mean), and your cancer tests positive for PD-L1.

Atezolizumab is a type of targeted therapy drug called an immune checkpoint inhibitor. It is a monoclonal antibody that works by binding to the protein PD-L1 Present on the exterior of certain cancerous cells, which keeps cancer cells from suppressing the immune system. ^256–260^

#### Radiotherapy

FDG is a radiopharmaceutical, specifically a radiotracer, used in the medical imaging modality positron emission tomography (PET). FDG is a glucose analog that concentrates in cells that rely upon glucose as an energy source, or in cells whose dependence on glucose increases under pathophysiological conditions. Li’s published work in 2021 aimed to establish a predictive model based on 18 F-FDG PET/CT for diagnosing the nature of pleural effusion (PE) Among individuals diagnosed with lung adenocarcinoma. Maiga’s published work in 2017 investigated The function of FDG PET/CT in diagnosing and determining the stage of lung cancer. FDG is not a chemotherapy drug and does not directly treat cancer. Instead, it is used to help diagnose cancer, heart disease, and epilepsy by detecting areas of increased metabolic activity in the body. ^261,262^

### Objective

There are some new papers propose RAIN protocol to treating diseases by combining Substances that induce the p-value associated with the relationship between the disease and target proteins/genes close to 1. The RAIN protocol acts such as systematic review and meta-analysis approach that employs the STROBE method to address a single medical question ^280,281^.

There are different newly published fundamental/ applicable artificial intelligence algorithms such as Graph Neural Network or Utilizing reinforcement learning for the purpose of medical tasks. ^263–272^

Such as various newly published medical AI papers, on the other hand, the RAIN protocol is unique in that it employs AI to address a specific medical question; suggesting combinations of drugs. Typically, They conduct a network meta-analysis to evaluate the efficacy in relation to effectiveness. ^280–289^

## 2. METHOD

The RAIN protocol involves three clear phases. Firstly, artificial intelligence is utilized to suggest the best medication compound tailored for treating and handling a particular illness. Next, a thorough examination is carried out using language comprehension and analysis, methodically scrutinizing recent literature and clinical experiments to evaluate how effective different variations of the recommended blend are. Figure 2 illustrates the article allocation among each section of the STROBE list of items to be checked. Lastly, during the third phase, the efficacy of medications along with their linked human proteins/genes is assessed through Network meta-analysis.

**Figure 1:**
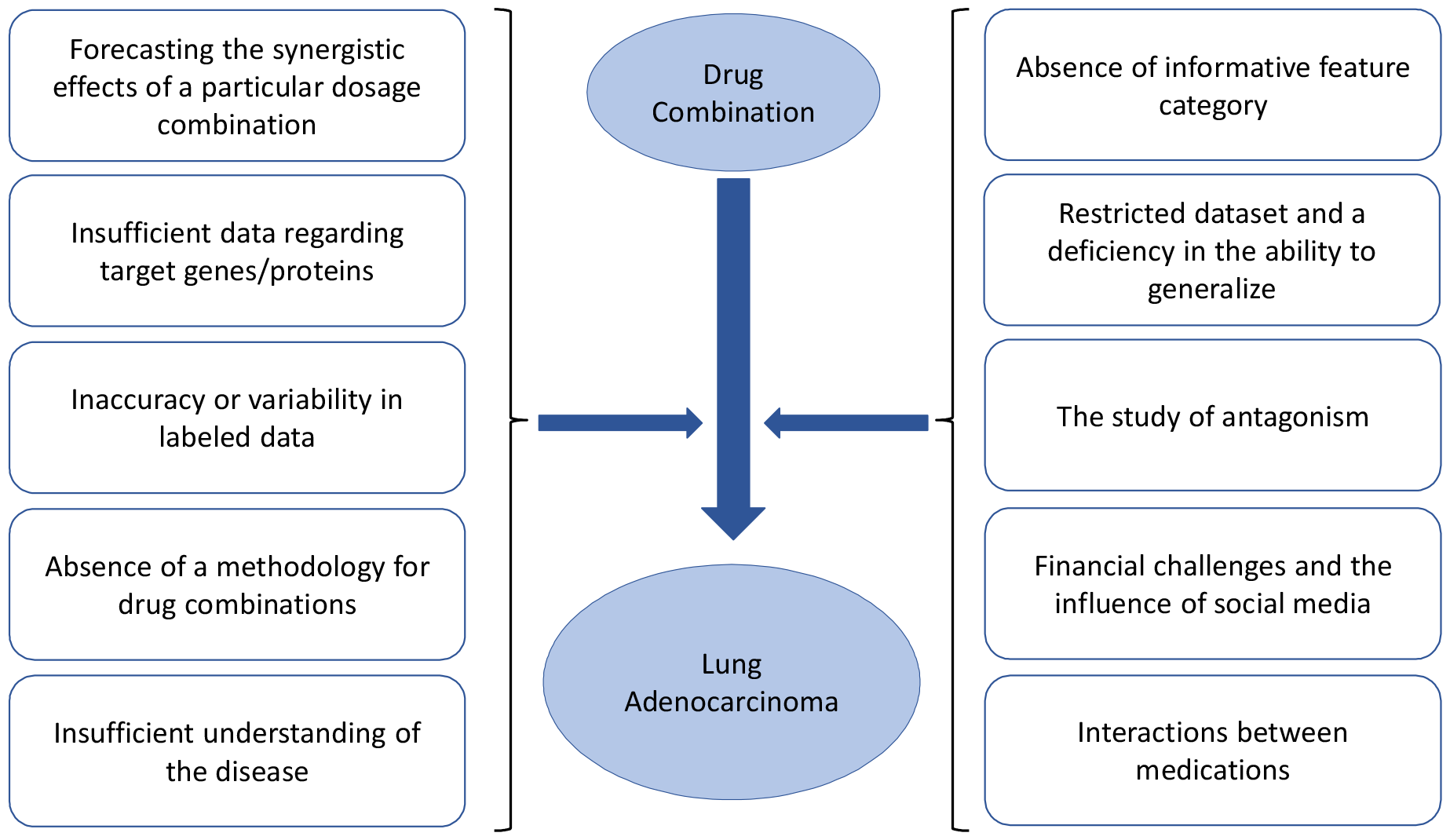
The impact of suggested drug combinations on the management of Lung adenocarcinoma cases

**Figure 2:**
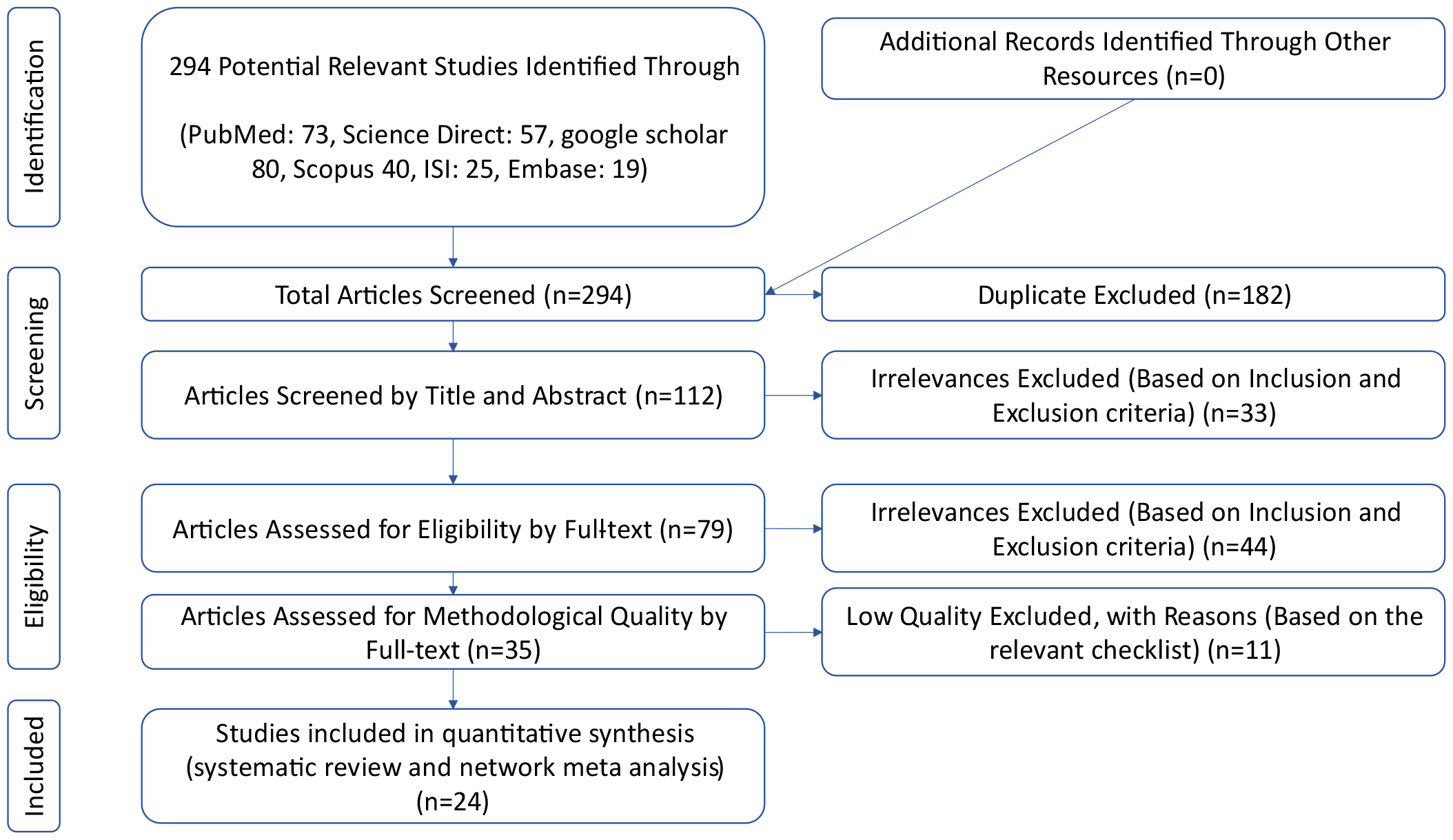
PRISMA (2020) flow chart illustrating the stages of article screening in the RAIN protocol

### Stage I: Recommending Drug Combinations

GraphSAGE is a framework for inductive representation learning on large-scale graphs, enabling the generation of node embeddings for unseen nodes during training. By sampling and aggregating information from the local neighborhood of each node, GraphSAGE can produce embeddings that encode both local and global structural information of the graph. With the ability to learn from nodes at different distances in the graph through multiple layers, GraphSAGE becomes a powerful tool for analyzing and understanding complex relational data.

The RAIN protocol integrates the GNN (Graph Neural Network) methodology to encourage cooperation between the connections aimed at achieving a particular objective. This tactic employs a network meta-analysis to pinpoint connections to established p-values, which are then converted into collaborations of restricted importance. The suggested model comprises two primary phases: the stages of the forward and reverse phases processing. Throughout the forward phase, the GNN algorithm computes aggregated p-values for the relationships between associations and the target variable, in the end choosing the connection with the lowest aggregated p-value. During the following reverse phase, p-values concerning characteristics of the interface and its intended purpose are modified according to the selected connections. The importance of these collaborative connections is assessed by Repeating their aggregated p-values. This repeating procedure persists up to the point of the importance diminishes to a pre-established limit. A graphic depiction of this procedure is depicted in Figure 3.

**Figure 3:**
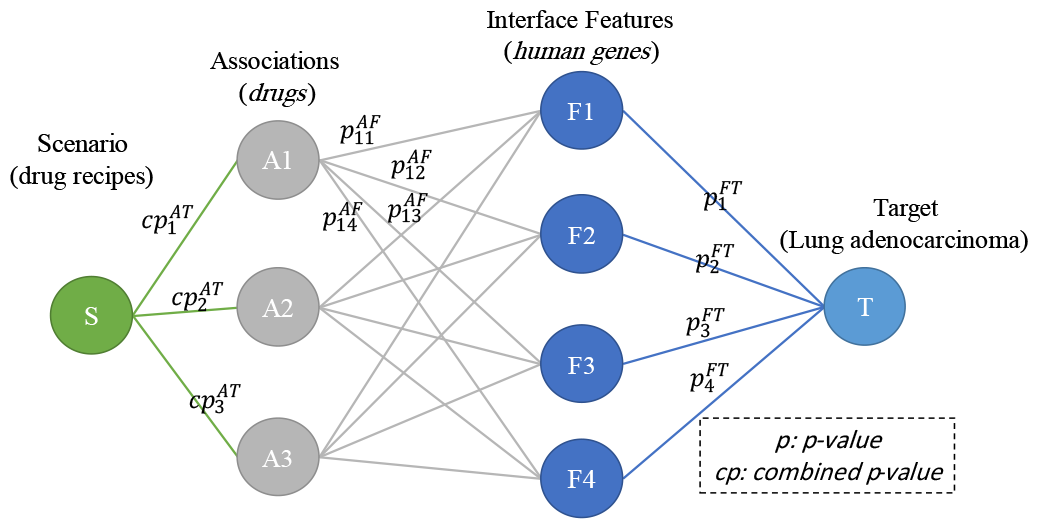
The overall framework of the GNN model for proposing efficient drug combinations in disease management, leveraging human proteins/genes as interface characteristics.

### Stage II: A comprehensive Systematic Review

In this systematic review and meta-analysis, the search queries include “Adenocarcinoma of the lung”, “Gefitinib”, “Paclitaxel” and “Icotinib”.

We use Natural Language Processing (NLP) in our search queries, since NLP is based on context not just keywords. For example, the keyword “Adenocarcinoma of the lung” in our NLP search is equal to the search query something like:

“Adenocarcinoma of the lung” OR “Lung Adenocarcinoma” OR “Lung Adenocarcinomas”.

PubMed search queries:

((((Adenocarcinoma of the lung[Title/Abstract]) OR (Lung Adenocarcinoma[Title/Abstract]) OR (Lung Adenocarcinomas[Title/Abstract])) AND (Gefitinib[Title/Abstract])) OR (((Adenocarcinoma of the lung[Title/Abstract]) OR (Lung Adenocarcinoma[Title/Abstract]) OR (Lung Adenocarcinomas[Title/Abstract])) AND (Paclitaxel[Title/Abstract])) OR (((Adenocarcinoma of the lung[Title/Abstract]) OR (Lung Adenocarcinoma[Title/Abstract]) OR (Lung Adenocarcinomas[Title/Abstract])) AND (Icotinib[Title/Abstract])))

Next, scholars conducted an extensive search across various databases including PubMed, WoS, Scopus, ScienceDirect, and Google Scholar (supplementary) without restricting the time frame. They utilized keywords such as GraphSAGE, Lung adenocarcinoma, human Genes, RAIN protocol, and machine learning. The gathered data were organized using information management software (Endnote), and additional efforts were made to manually review the sources cited in the identified articles, aiming to encompass as many relevant studies as possible.

#### Inclusion and exclusion criteria

Inclusion criteria for the study comprised: 1) Research encompassing the term lung adenocarcinoma along with at least one of the suggested medications for this condition. 2) Cross-sectional studies. 3) Availability of full-text studies. 4) Studies offering adequate data, including sample size and prevalence. Exclusion criteria included: 1) Studies presented at conferences. 2) Case report studies. 3) Case series studies. 4) Duplicate studies.

#### Selection of studies

Researchers utilized EndNote software to manage the studies. Initially, duplicates across various databases were eliminated. During the primary assessment, titles and abstracts were meticulously reviewed, excluding articles irrelevant to the research aim. Subsequently, in the secondary assessment, full-text articles were scrutinized, and those meeting inclusion criteria were retained. Thirty-five articles proceeded to the qualitative evaluation phase. To mitigate bias and errors, all stages including article search, study selection, quality assessment, and data extraction were independently conducted by two researchers. Disagreements regarding study inclusion were resolved through discussion involving a third researcher to ensure impartiality. A final consensus was achieved through deliberation.

#### Evaluation of the quality of studies

To assess and ensure the quality of articles, a checklist designed for observational studies was employed. This checklist, known as the Strengthening the Reporting of Observational Studies in Epidemiology (STROBE), comprises six scales: title, abstract, introduction, methods, results, and discussion, encompassing a total of 32 items. These items cover various methodological aspects of the study, including the title’s clarity, statement of the problem, study objectives, study type, statistical population, sampling method, determination of sample size, definition of variables and procedures, data collection tools, statistical analysis methods, and findings interpretation. Articles scoring 16 or higher on this checklist were deemed to possess good or moderate methodological quality and were included in the study, while those scoring below 16 were considered to have poor methodological quality and were therefore excluded.

#### Data extraction

Two researchers conducted data extraction using a pre-established checklist. This checklist encompassed the author’s name, publication year, research location, sample size, gender and age distribution of participants, data collection tool, and prevalence percentage.^290^

### Stage III: Network meta-analysis

In the third stage, a network meta-analysis is employed to explore the influence of possible combinations of artificial medications affecting human proteins and genes. This methodology facilitates the assessment of numerous medications in a unified research, amalgamating data from both direct and indirect sources originating from randomized trials linking conditions treated with medications via proteins and genes. Through the utilization of proteins and genes as pivotal elements in a network, this examination aids in discerning the comparative efficacy of frequently prescribed medications in practical clinical scenarios. The assessment of individual drug’s effectiveness relies on biological information provided.

## 3. RESULTS

### Stage I: Graph Neural network

The medication blend recommended by the GNN comprises Gefitinib, paclitaxel, and Icotinib, with Table 1 illustrating the associated p-values for this combination’s significance. For example, examining the p-value associated with lung adenocarcinoma and Gefitinib (Scenario 1), the value is 0.01531. This value significantly reduces to 0.005595 upon the addition of Paclitaxel (Scenario 2), and further drops to 0.002858 in the third scenario, indicating a positive effect of the proposed drug combination on disease control.

**Table 1:** p-value associated with scenarios and Lung adenocarcinoma.

| SCENARIO | DRUG COMBINATION | P-VALUE |
| --- | --- | --- |
| **S1** | Gefitinib | 0.01531 |
| **S2** | S1 + paclitaxel | 0.005595 |
| **S3** | S2 + Icotinib | 0.002858 |

Table 2 presents an analysis of how p-values between lung adenocarcinoma and human proteins/genes fluctuate across various situations. The column labeled ‘S0’ displays the p-values indicating the associations between lung adenocarcinoma and the corresponding human proteins/genes that are affected. Upon introducing Gefitinib (S1 column), the aggregated p-values are illustrated. Notably, in the ‘S3’ column, the p-values between Lung adenocarcinoma and human proteins/genes reach 1, suggesting a reduction in the importance of the target proteins/genes.

**Table 2:** p-values indicating the relationship between Lung adenocarcinoma and human proteins/genes following the implementation of various scenarios.

| Association Name | p-value | S1 | S2 | S3 | Association Name | p-value | S1 | S2 | S3 |
| --- | --- | --- | --- | --- | --- | --- | --- | --- | --- |
| **MALAT1** | 3.04E-06 | 0.961228 | 0.994803 | 0.994803 | **GNPNAT1** | 0.00018 | 0.00018 | 0.945391 | 0.945391 |
| **EGFR** | 3.71E-06 | 0.999999 | 1 | 1 | **TMPO-AS1** | 0.000209 | 0.000209 | 0.000209 | 0.000209 |
| **KRAS** | 8.34E-06 | 0.999865 | 1 | 1 | **LINC01116** | 0.000211 | 0.995882 | 0.999957 | 0.999957 |
| **ROS1** | 9.4E-06 | 0.999521 | 0.999995 | 1 | **METTL3** | 0.000213 | 0.988153 | 0.988153 | 0.988153 |
| **NKX2-1** | 1.41E-05 | 0.995558 | 0.9997 | 0.999998 | **RNMT** | 0.000234 | 0.999868 | 0.999981 | 0.999981 |
| **TTF1** | 1.59E-05 | 0.99602 | 0.999871 | 0.999999 | **NFYC-AS1** | 0.000284 | 0.000284 | 0.000284 | 0.000284 |
| **ALK** | 2.01E-05 | 0.99796 | 0.999733 | 0.999733 | **KEAP1** | 0.000287 | 0.908733 | 0.994849 | 0.999773 |
| **NAPSA** | 2.37E-05 | 0.850868 | 0.983292 | 0.99991 | **LINC00857** | 0.000289 | 0.000289 | 0.000289 | 0.000289 |
| **CD274** | 3.08E-05 | 0.998557 | 1 | 1 | **KIF5B** | 0.000319 | 0.000319 | 0.965614 | 0.965614 |
| **TXK** | 3.77E-05 | 0.999997 | 1 | 1 | **ZEB1** | 0.000329 | 0.996797 | 0.999983 | 0.999983 |
| **EML4** | 4.37E-05 | 0.999439 | 0.999988 | 1 | **CCNB2** | 0.000331 | 0.000331 | 0.557733 | 0.557733 |
| **STK11** | 4.79E-05 | 0.994139 | 0.999517 | 0.999982 | **LMO7DN** | 0.000336 | 0.000336 | 0.000336 | 0.000336 |
| **FAM83A-AS1** | 4.98E-05 | 4.98E-05 | 4.98E-05 | 4.98E-05 | **ERO1L** | 0.000338 | 0.000338 | 0.000338 | 0.000338 |
| **TP53** | 5.84E-05 | 0.996982 | 0.999999 | 1 | **KRT7** | 0.000342 | 0.709673 | 0.992221 | 0.992221 |
| **BRAF** | 6.07E-05 | 0.999371 | 0.999993 | 1 | **TBX5-AS1** | 0.000364 | 0.000364 | 0.98475 | 0.98475 |
| **RET** | 6.98E-05 | 0.999995 | 1 | 1 | **SFTA1P** | 0.000368 | 0.000368 | 0.000368 | 0.999676 |
| **FAM83A** | 7.5E-05 | 0.978833 | 0.978833 | 0.978833 | **BCL2** | 0.000373 | 0.997887 | 1 | 1 |
| **RBM10** | 0.000104 | 0.950423 | 0.950423 | 0.999974 | **LINC00592** | 0.000379 | 0.000379 | 0.989935 | 0.989935 |
| **ERBB2** | 0.000138 | 0.999955 | 1 | 1 | **LINC00968** | 0.000385 | 0.000385 | 0.000385 | 0.000385 |
| **MET** | 0.000148 | 0.999943 | 0.999999 | 1 | **CDH2** | 0.000388 | 0.99727 | 0.999976 | 0.999997 |
| **SYNPR-AS1** | 0.000151 | 0.000151 | 0.000151 | 0.000151 | **LINC00941** | 0.000404 | 0.000404 | 0.000404 | 0.000404 |
| **CDH1** | 0.000152 | 0.999189 | 0.999998 | 1 | **CHIAP2** | 0.000421 | 0.000421 | 0.000421 | 0.000421 |
| **PIK3CA** | 0.000158 | 0.999322 | 0.999999 | 1 | **WWC2-AS2** | 0.000421 | 0.000421 | 0.000421 | 0.000421 |
| **PDCD1** | 0.000168 | 0.992931 | 0.999993 | 0.999993 |  |  |  |  |  |

### Stage II: A comprehensive Systematic Review

The examination of the impact of the specified medications on the treatment of lung adenocarcinoma reached an advanced stage. A meticulous selection procedure, adhering to the PRISMA guidelines and the RAIN framework, was employed to compile pertinent articles up to the point when this article was written. In total, 294 articles deemed potentially relevant were identified and incorporated into the EndNote reference management system. Following the elimination of duplicates (182 articles), the remaining 112 papers underwent scrutiny through title and abstract review, utilizing predetermined criteria for including and excluding. As a result, 33 studies were eliminated during the screening process. Subsequently, during the suitability assessment, 79 studies met the criteria, and their complete texts were comprehensively examined. After applying the criteria for inclusion and exclusion, 44 researches were subsequently excluded. Throughout the stage of assessing quality, the remaining 35 research papers underwent assessment utilizing the STROBE checklist ratings and methodological rigor, leading to the removal of 11 researches considered to be of low quality. Eventually, the final analysis incorporated 24 cross-sectional studies. A thorough examination of the complete texts of these articles took place, and every individual article received a score based on the STROBE checklist (refer to Figure 3). Figure 4 illustrates the chemical compositions of the drugs utilized in the research, and Table 3 presents a summary of the properties associated with these drugs. Additional details and characteristics can be found in Table 4.^291–314^

**Table 3:** Characteristics of potential medications for effective management of lung adenocarcinoma.

| Drug name | Accession Number | Type |  | Formula | Mechanism of Action |
| --- | --- | --- | --- | --- | --- |
|  |  | Small | Biotech |  |  |
| Gefitinib | DB00317 | * |  | C22H24ClFN4O3 | Gefitinib functions as an inhibitor of the tyrosine kinase linked to the epidermal growth factor receptor (EGFR), targeting the adenosine triphosphate (ATP)-binding region of the enzyme. Elevated expression of EGFR is frequently observed in specific human carcinoma cells, such as those found in lung and breast cancers. This increased expression leads to heightened activation of anti-apoptotic Ras signal transduction pathways, ultimately promoting enhanced survival and uncontrolled proliferation of cancer cells. Gefitinib, recognized as the initial a specific inhibitor of EGFR tyrosine kinase, additionally known as Her1 or ErbB-1, hinders the downstream signaling cascades. Consequently, malignant cell proliferation is restrained through the inhibition of EGFR tyrosine kinase. |
| Paclitaxel | DB01229 | * | | C47H51NO14 | Paclitaxel disrupts the normal growth function of microtubules. In contrast to substances like colchicine that induce microtubule depolymerization in vivo, paclitaxel halts their function by hyper-stabilizing their structure. This disrupts the cell's ability to utilize its cytoskeleton with flexibility. Specifically, paclitaxel associates with the $\beta$ subunit of tubulin, the fundamental component of microtubules, immobilizing these building blocks. The resulting paclitaxel-bound microtubule complex lacks the capability to disassemble. This interference negatively impacts cell function as the dynamic instability, involving microtubule shortening and lengthening, crucial for their role as a cellular transportation network, is compromised. Processes like mitosis, where chromosomes depend on this microtubule property, are affected. Additionally, recent investigations suggest that paclitaxel triggers programmed cell death (apoptosis) in cancer cells by binding to Bcl-2 (B-cell leukemia 2), a protein that inhibits apoptosis, thereby inhibiting its function. |
| Icotinib | DB11737 | * |  | C22H21N3O4 | Icotinib is a remarkably specific first-generation a substance that blocks the activity of the tyrosine kinase associated with the epidermal growth factor receptor (EGFR-TKI). It attaches reversibly to the ATP binding site of the EGFR protein, impeding the progression of the pathway of cellular signaling . EGFR functions as an oncogenic receptor, and individuals with activating somatic genetic variations, for example an exon 19 deletion or exon 21 L858R genetic variations, in the tyrosine kinase domain, experience uncontrolled cell proliferation. |

**Table 4:** Several significant research studies have been conducted concerning potential medications for managing lung adenocarcinoma.

| study name | Gefitinib | Paclitaxel | Icotinib |
| --- | --- | --- | --- |
| Chen H et al., 2023 | * | * |  |
| Hu L et al., 2023 | * | * |  |
| Shen L et al., 2023 | * |  | * |
| Wang X et al., 2023 | * | * |  |
| Bai X et al., 2022 | * |  | * |
| Chen H et al., 2022 | * | * |  |
| Hsieh PC et al., 2022 | * |  | * |
| Yan Q et al., 2022 | * | * |  |
| Zhang H et al., 2022 |  | * | * |
| Zhang L et al., 2022 | * |  | * |
| Zhang Q et al., 2022 | * |  | * |
| Zhao R et al., 2022 | * | * |  |
| Bie Y et al., 2021 | * |  | * |
| Cai J et al., 2021 | * |  | * |
| Chen H et al., 2021 | * | * |  |
| Du X et al., 2021 | * |  | * |
| Jiang S et al., 2021 | * |  | * |
| Liu B et al., 2021 | * | * |  |
| Xiang C et al., 2021 | * |  | * |
| Qian J et al., 2020 | * |  | * |
| Ruan X et al., 2020 | * |  | * |
| Shang Y et al., 2020 | * |  | * |
| Šťastná N et al., 2020 | * | * |  |
| Wang Y et al., 2020 | * |  | * |

**Figure 4:**
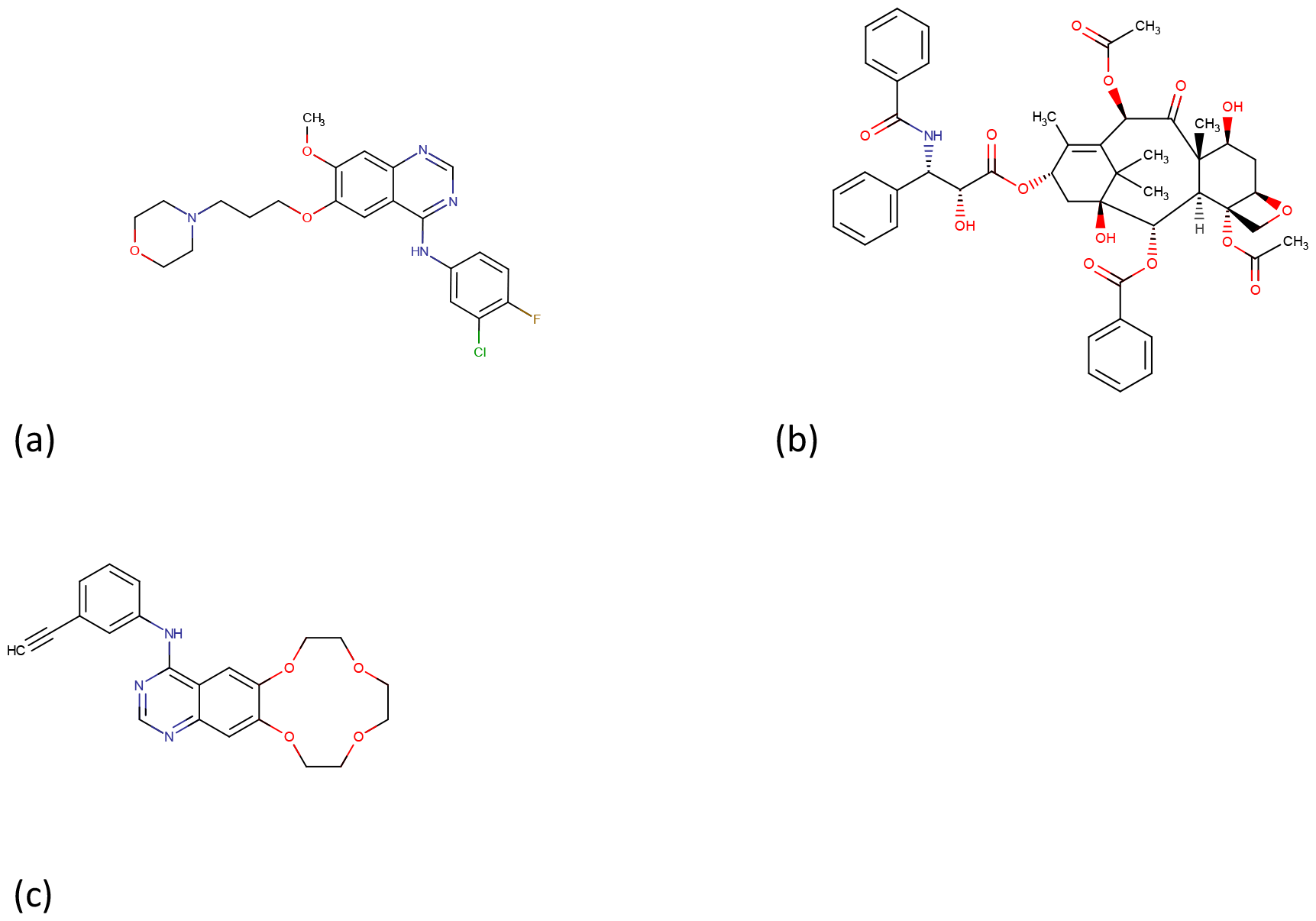
Drug structure for (a) Gefitinib, (b) Paclitaxel, (c) Icotinib from https://www.drugbank.com/

### Stage III: Network Meta-Analysis

Figure 5 displays the p-values associated with human proteins/genes impacted by Lung adenocarcinoma, while Figure 6 showcases the p-values resulting from the third scenario’s execution. The effectiveness of drugs selected via the drug selection algorithm is illustrated in Figure 7 using a radar chart, showcasing the statistical significance (p-values) of the relationships among lung adenocarcinoma and human proteins/genes following the usage of the chosen medications; the significance between each gene and the drugs are shown in Supplemental Table 1. Figure 8 depicts the p-values associated with relationships and targets utilizing alternative interface characteristics. P-values below 0.01 are depicted in green, while those between 0.01 and 0.05 are shown in blue. Each colored line on the radar chart represents the efficacy of the corresponding drug in the specified scenario.

**Figure 5:**
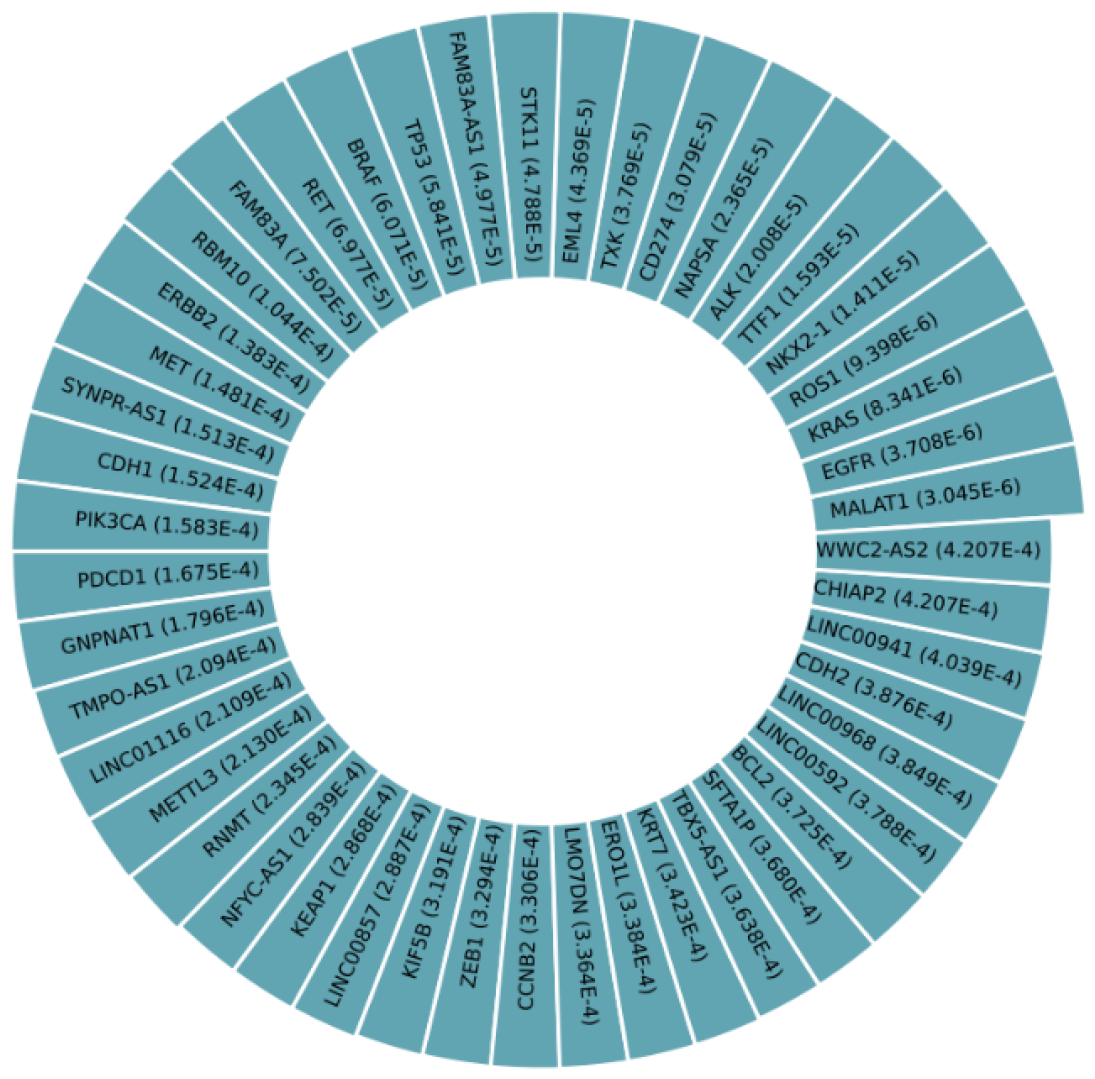
The p-values indicating the relationship between impacted human proteins/genes and Lung adenocarcinoma.

**Figure 6:**
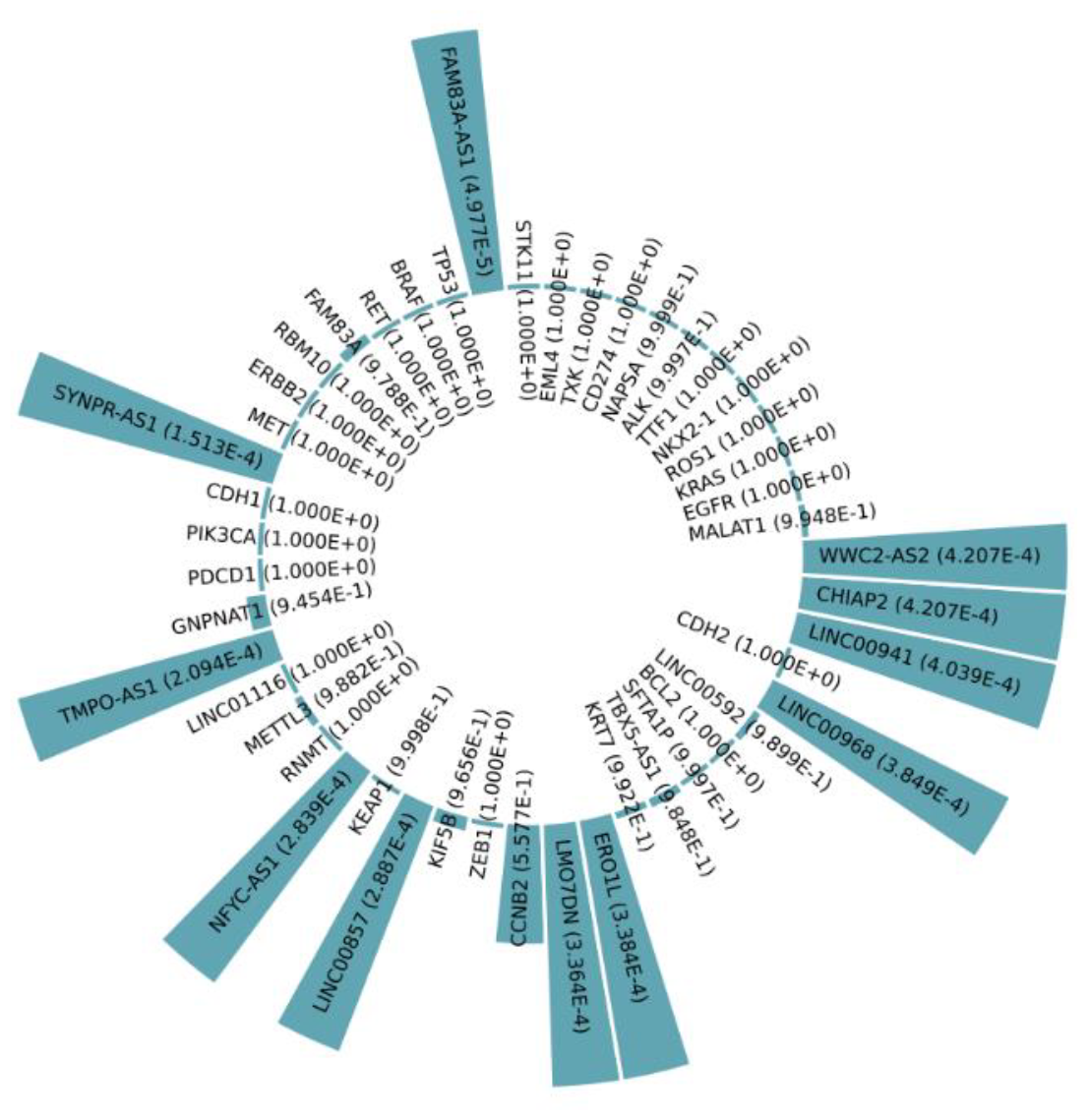
After applying Scenario 4, the p-values associated with human proteins/genes affected by Lung adenocarcinoma were computed.

**Figure 7:**
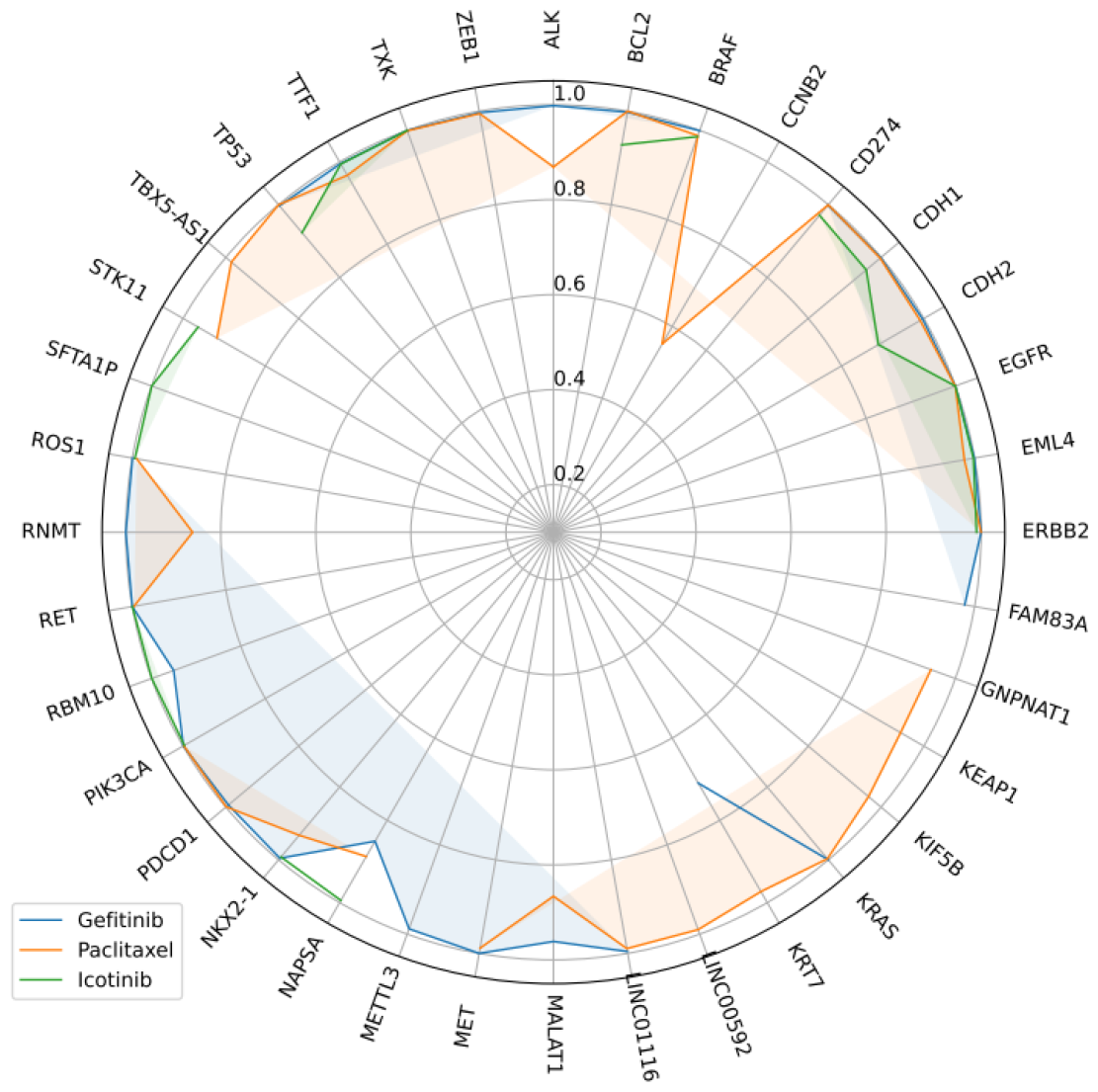
Radar chart depicting p-values indicating associations between lung adenocarcinoma and affected proteins/genes following the administration of each drug.

**Figure 8:**
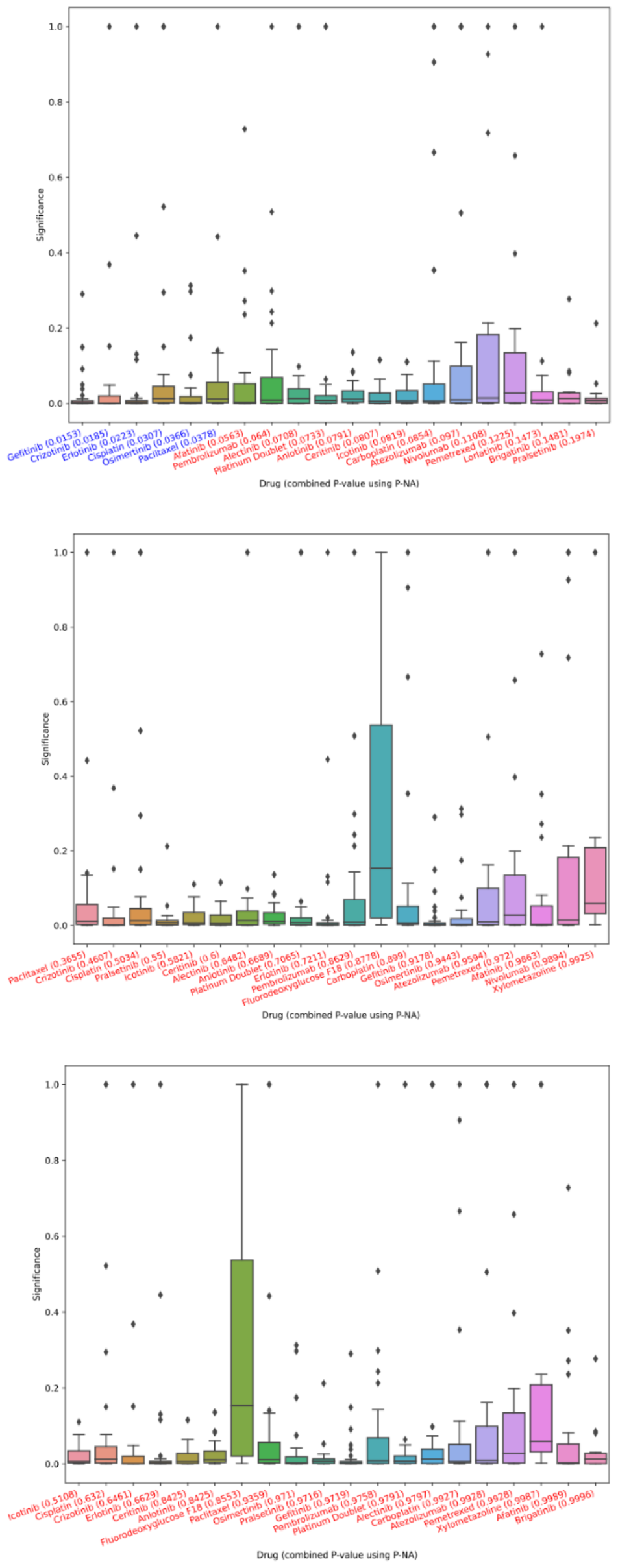
P-values indicating associations between specific interface features and the target variable across different stages: (a) Overall, (b) after the first drug from (a) is administered, (c) after the first drugs from (a) and (b) are administered.

## 4. DISCUSSION

We utilized our approach on a dataset comprising nodes and edges symbolizing the network, with each node denoting either a drug or a gene, and every edge indicating a corresponding p-value. Our findings revealed that the graph neural network suggests the amalgamation of Gefitinib, Paclitaxel, and Icotinib as the optimal drug combination for targeting proteins/genes linked to this specific cancer. An examination of clinical trials and expert viewpoints regarding these medications substantiated our assertion. Additionally, the network meta-analysis provided further validation of the effectiveness of these medications on the related genes.

Information about prescribed medications plays a critical role in exploring potential interactions, adverse effects, and the associated risks related to drug usage. To identify these details, diverse reliable online platforms such as Medscape, WebMD, Drugs, and Drugbank were referenced to enable a thorough analyzing different medications for comparison. Utilizing these dependable databases, an in-depth evaluation of combinations of medications was carried out, Eventually uncovering specific interactions between pairs of medications. It is crucial to highlight that we observed certain interactions between medications.

The specified medication (Gefitinib) is processed by the CYP3A5 enzyme, and the influenced drug (Paclitaxel) is also a CYP3A5 substrate with a limited therapeutic range. There is a potential for metabolic competition since both drugs are metabolized by the same enzyme. Medications with a restricted therapeutic range need to be carefully regulated within a specific concentration range to ensure safety and effectiveness. Elevated concentrations resulting from metabolic competition in drugs with a narrow therapeutic range could result in severe adverse effects and toxicity.^315–318^

The metabolism of Gefitinib may be enhanced when co-administered with Icotinib. The prescribed amount of gefitinib for managing metastatic non-small cell lung cancer (NSCLC) in individuals with tumors featuring epidermal growth factor receptor (EGFR) exon 19 deletions or exon 21 (L858R) substitution mutations, as confirmed by an FDA-approved test, is 250mg administered orally on a daily basis.^319^

The suggested dosage of icotinib for treating advanced lung adenocarcinoma in Individuals with the EGFR exon 21 L858R mutation is 250mg three times a day (tid). The effectiveness and tolerability of a high dosage of icotinib (250mg tid) For patients diagnosed with advanced lung adenocarcinoma, carrying the EGFR exon 21 L858R mutation were validated in a phase II trial. However, in cases of adverse events, the icotinib dose may be adjusted, with a reduction of no more than two dose levels. This adjustment includes a change to 250mg twice daily or 125mg three times daily in the high-dose group, or 125mg twice daily in the routine-dose group.^320–322^

The metabolic rate of Paclitaxel may decrease when administered in combination with Gefitinib. When combined with Icotinib, the metabolism of Paclitaxel may be accelerated.

The amount of paclitaxel needed to treat lung adenocarcinoma varies depending on factors like the patient’s overall health, the stage of cancer, and whether it’s given alone or with other medications. In cases where patients haven’t received treatment before, paclitaxel is usually given through an IV at a dose of 175 mg/m^2^ over a 3-hour period every 3 weeks. In the treatment of advanced or metastatic lung adenocarcinoma, paclitaxel is frequently paired with cisplatin. This combined therapy typically consists of administering paclitaxel intravenously at a dose of 135 mg/m^2^ over 24 hours, followed by cisplatin. Dosages can be adjusted according to factors such as renal function, neuropathy, and other relevant considerations. It’s crucial to monitor blood counts, liver function, and kidney function regularly throughout the treatment process. Additionally, all patients should receive premedication to prevent severe hypersensitivity reactions. Refer to the manufacturer’s guidelines for precise recommendations regarding premedication.^323–325^

## 5. CONCLUSION

In conclusion, our study presents a pioneering approach to recommending combinations of trending drugs aimed at targeting cancer-associated proteins/genes, leveraging graphSAGE within the RAIN protocol. Through a three-step framework, we effectively identified Gefitinib, Paclitaxel, and Icotinib as one highly effective drug combination for addressing adenocarcinoma of the lung. This recommendation then, was substantiated by clinical trial data; affirming its potential clinical utility. Moreover, our network meta-analysis further validated the efficacy of these drugs in modulating associated genes. With a p-value of 0.002858, we reject the null hypothesis, supporting the conclusion that Gefitinib, paclitaxel, and icotinib constitute one of the most potent combinations with a significant therapeutic impact on lung adenocarcinoma cancer.

By providing a novel tool for drug combination recommendation, our method offers valuable insights for clinicians and researchers striving to optimize treatment strategies and deepen their understanding of disease mechanisms.

## Data Availability

Datasets are available through the corresponding author upon reasonable request.

## Abbreviations

STROBE: Strengthening the Reporting of Observational studies in Epidemiology
PRISMA: Preferred Reporting Items for Systematic Reviews and Meta-Analysis
RAIN: Systematic Review and Artificial Intelligence Network Meta-Analysis

## Authors’ contributions

**Sogand Sadeghi:** Data Gathering, Writing original draft. **Ali A. Kiaei:** Conceptualization, Methodology, Review & Editing, AI model implementation, Formal analysis, Supervision. **Mahnaz Boush:** Conceptualization, Methodology, AI model implementation. **Nader Salari:** Conceptualization. **Masoud Mohammadi:** Review & Editing, Validation. **Danial Safaei:** Review & Editing. **Mitra Mahboubi:** Investigation,. **Arian Tajfam:** Investigation, Review & Editing. **Sabra Moghadam:** Investigation

## Availability of data and materials

Datasets are available through the corresponding author upon reasonable request.

## Ethics approval and consent to participate

Not applicable.

## Consent for publication

Not applicable.

