## Supplemental Table 1 for "A graphSAGE discovers synergistic combinations of Gefitinib, paclitaxel, and Icotinib for Lung adenocarcinoma management by targeting human genes and proteins: the RAIN protocol"

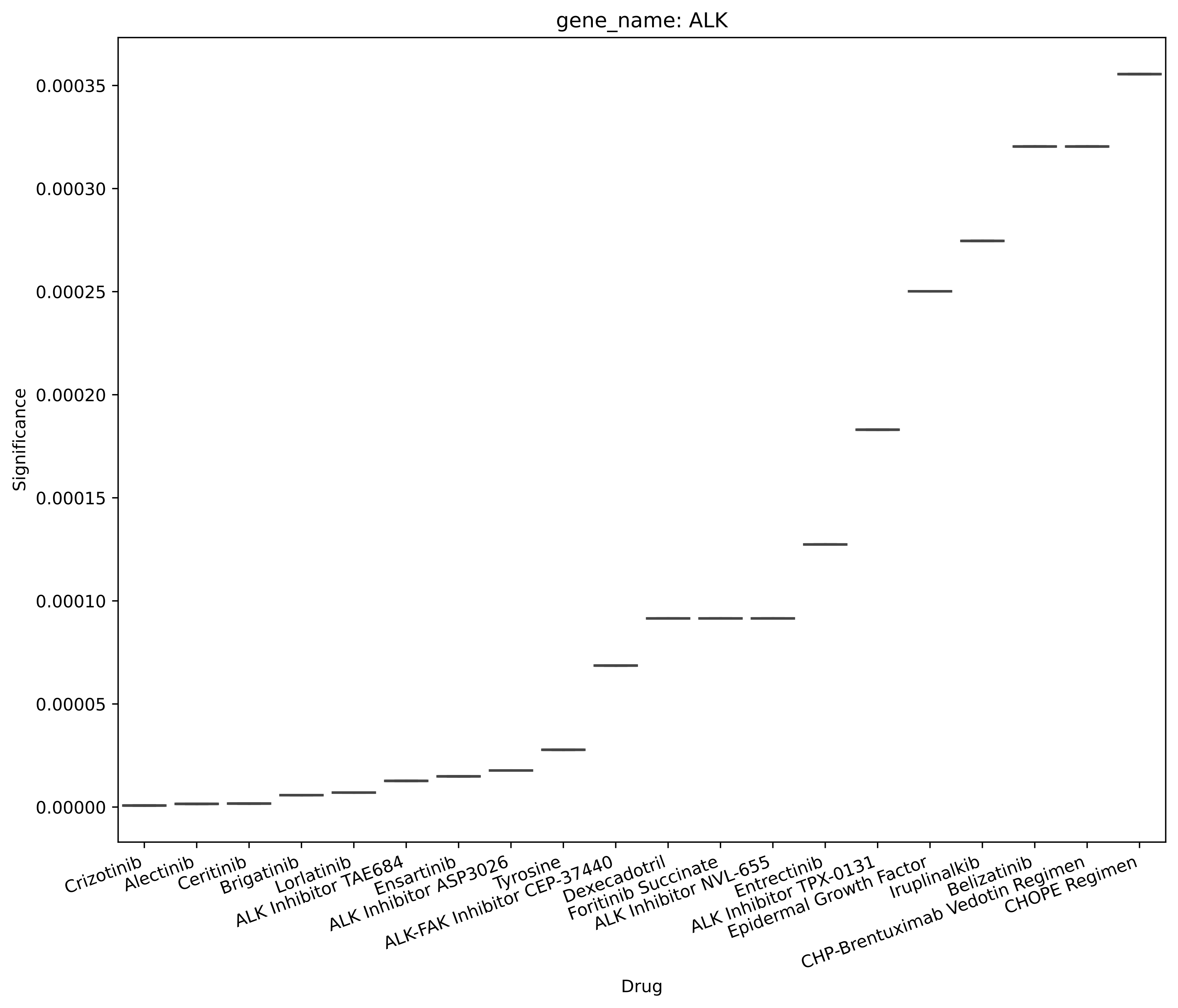
gene_name: ALK
Significance
0.00035
0.00030
0.00025
0.00020
0.00015
0.00010
0.00005
0.00000
Crizotinib
Alectinib
Ceritinib
Brigatinib
Lorlatinib
ALK Inhibitor TAE684
Ensartinib
ALK Inhibitor ASP3026
Tyrosine
ALK-FAK Inhibitor CEP-37440
Dexecadotril
Foritinib Succinate
ALK Inhibitor NVL-655
Entrectinib
ALK Inhibitor TPX-0131
Epidermal Growth Factor
Iruplinalkib
Belizatinib
CHP-Brentuximab Vedotin Regimen
CHOPE Regimen
Drug

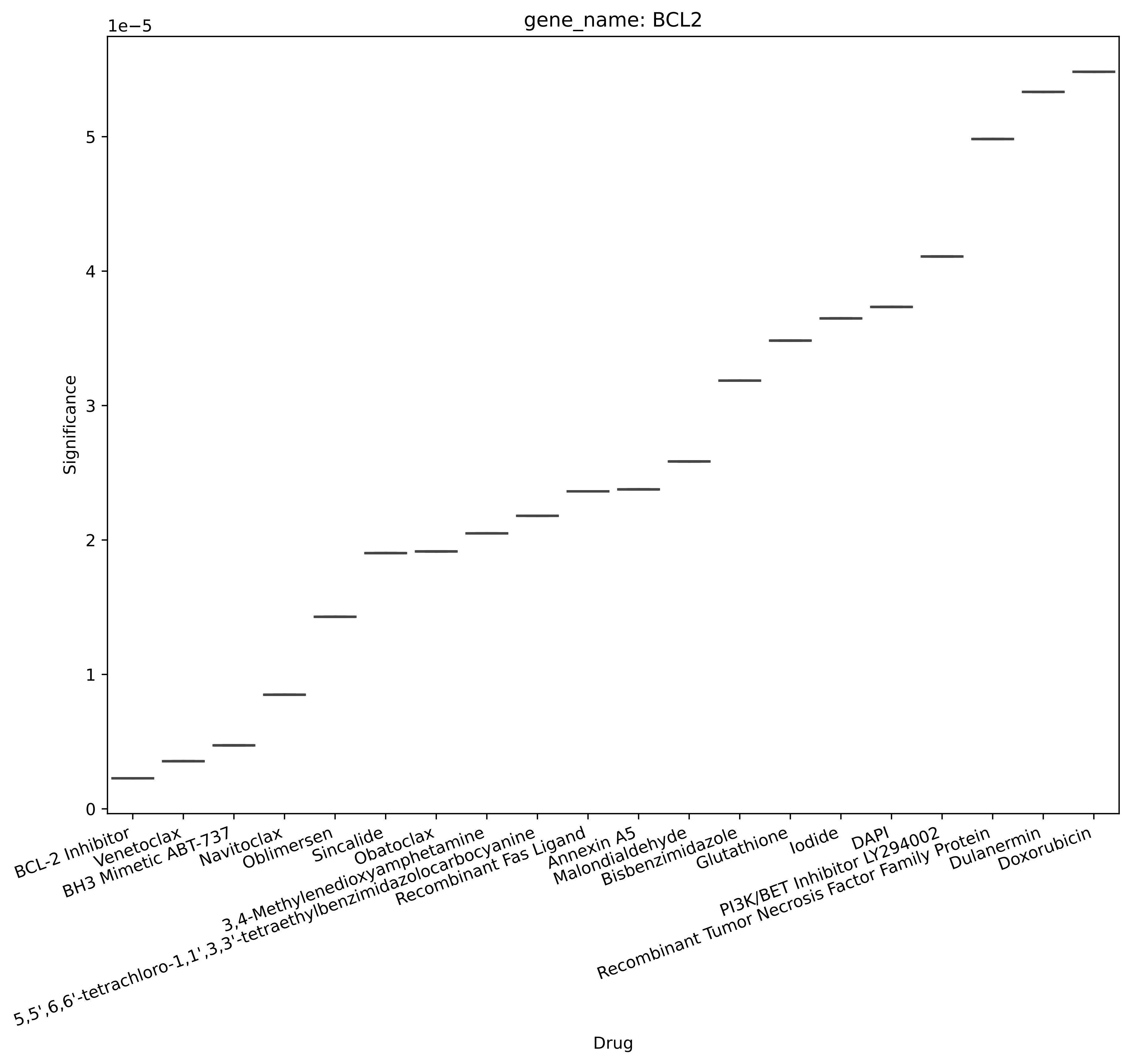
gene_name: BCL2
1e−5
Significance
0
1
2
3
4
5
BCL-2 Inhibitor
Venetoclax
BH3 Mimetic ABT-737
Navitoclax
Oblimersen
Sincalide
Obatoclax
3,4-Methylenedioxyamphetamine
5,5',6,6'-tetrachloro-1,1',3,3'-tetraethylbenzimidazolocarbocyanine
Recombinant Fas Ligand
Annexin A5
Malondialdehyde
Bisbenzimidazole
Glutathione
Iodide
DAPI
PI3K/BET Inhibitor LY294002
Recombinant Tumor Necrosis Factor Family Protein
Dulanermin
Doxorubicin
Drug

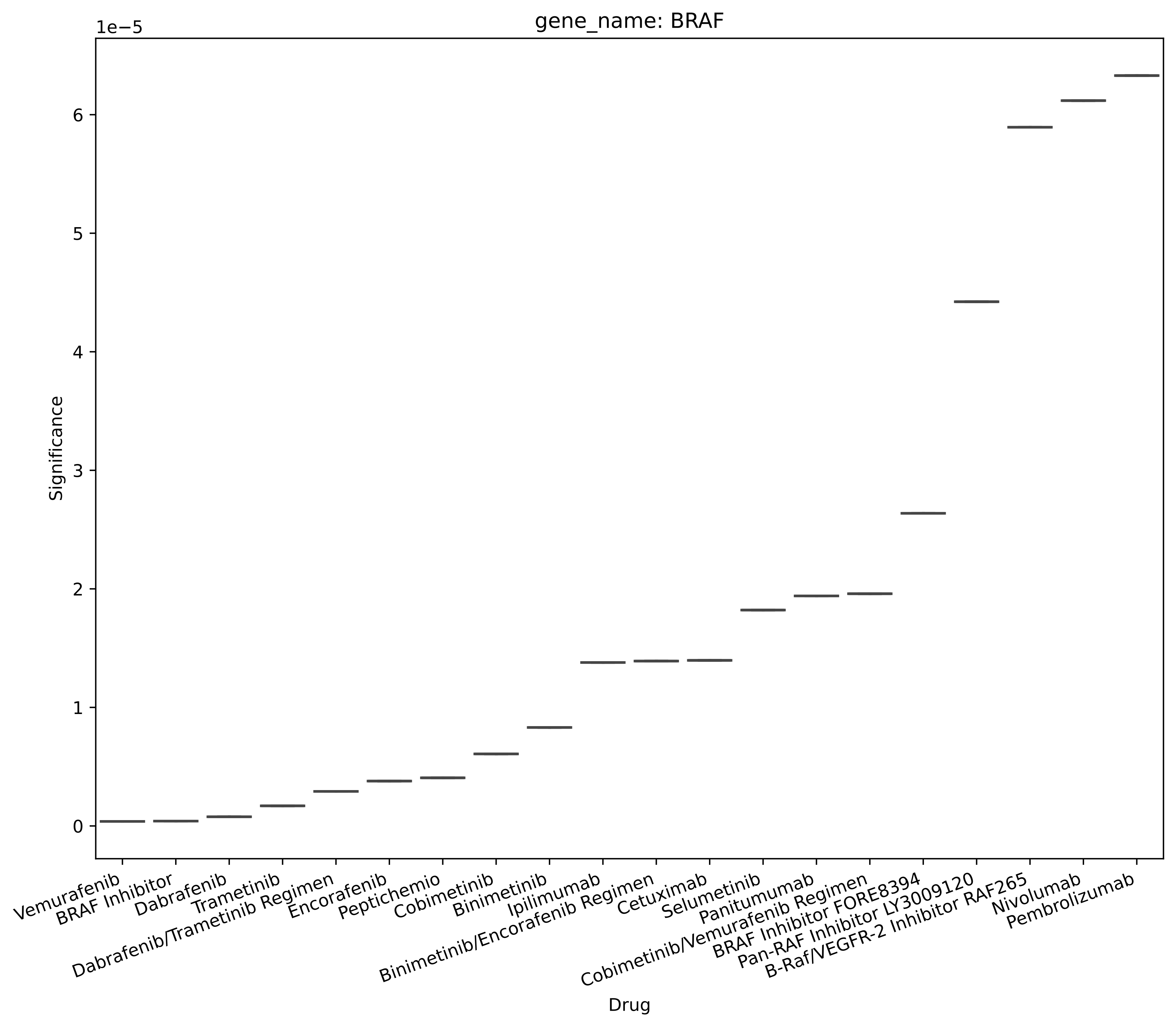

gene_name: BRAF
1e−5
6
5
4
3
2
1
0
Significance
Vemurafenib
BRAF Inhibitor
Dabrafenib
Trametinib
Dabrafenib/Trametinib Regimen
Encorafenib
Peptichemio
Cobimetinib
Binimetinib
Ipilimumab
Binimetinib/Encorafenib Regimen
Cetuximab
Selumetinib
Panitumumab
Cobimetinib/Vemurafenib Regimen
BRAF Inhibitor FORE8394
Pan-RAF Inhibitor LY3009120
B-Raf/VEGFR-2 Inhibitor RAF265
Nivolumab
Pembrolizumab
Drug

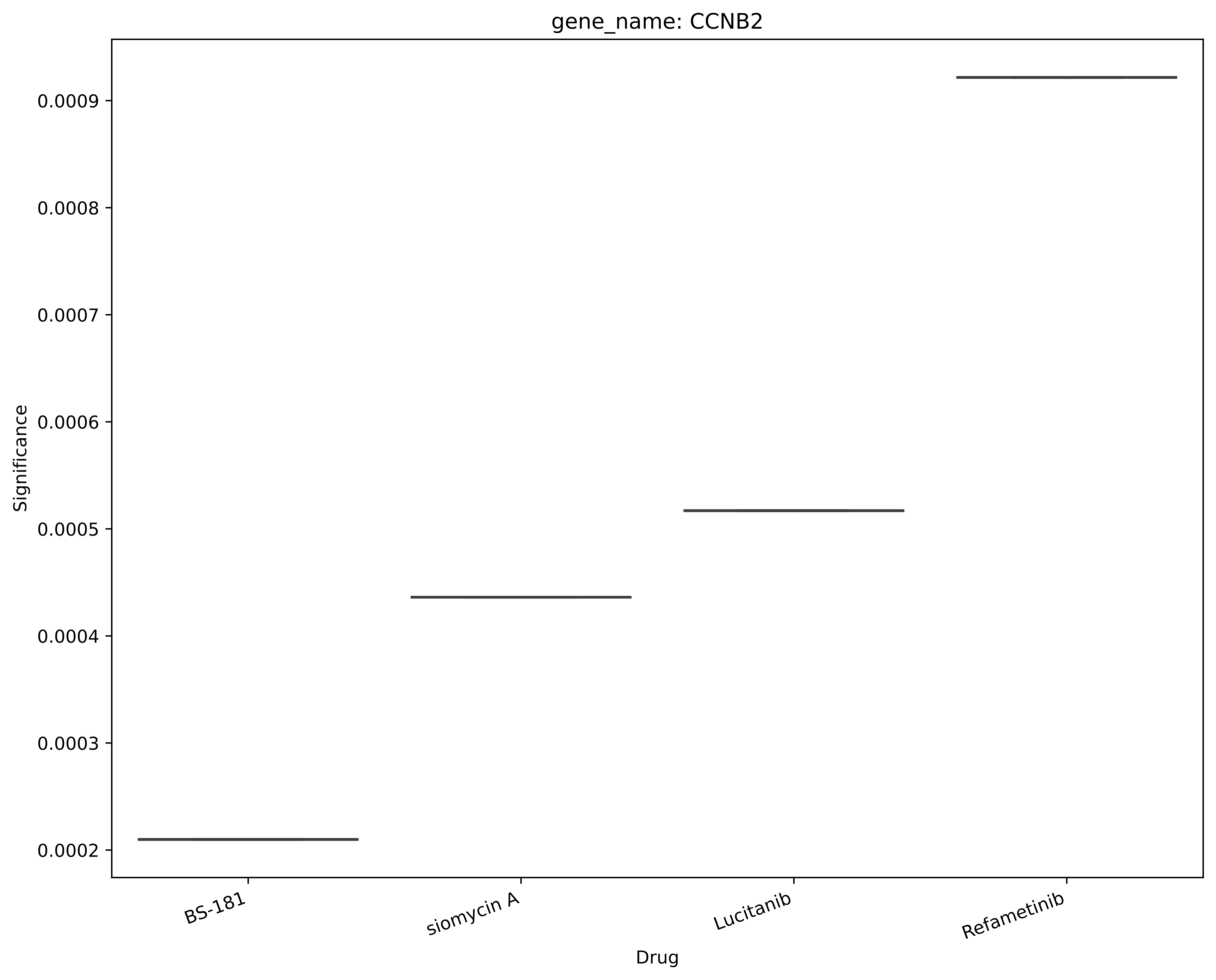
gene_name: CCNB2
Significance
0.0009
0.0008
0.0007
0.0006
0.0005
0.0004
0.0003
0.0002
BS-181
siomycin A
Lucitanib
Refametinib
Drug

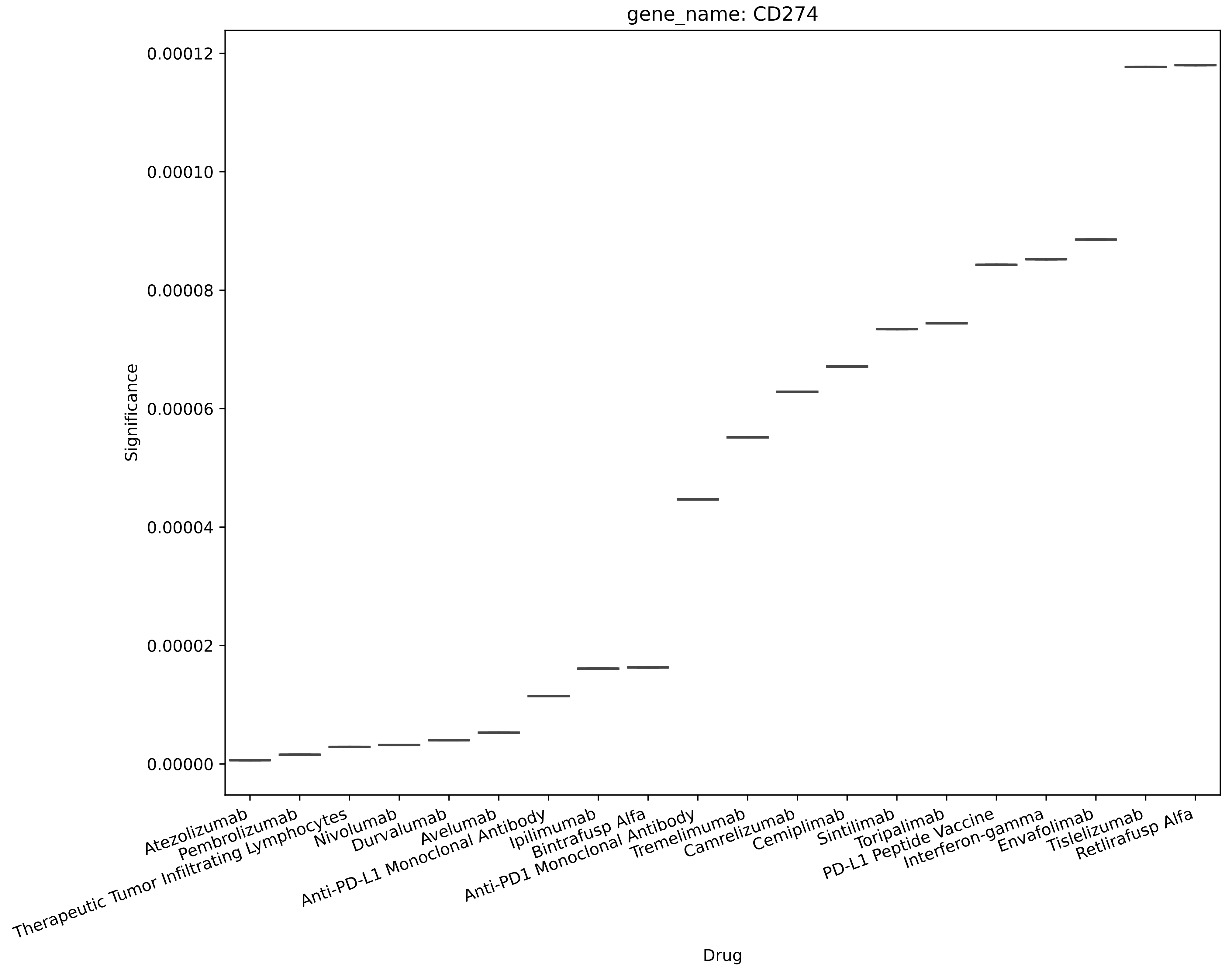
gene_name: CD274
Significance
0.00012
0.00010
0.00008
0.00006
0.00004
0.00002
0.00000
Atezolizumab
Pembrolizumab
Therapeutic Tumor Infiltrating Lymphocytes
Nivolumab
Durvalumab
Avelumab
Anti-PD-L1 Monoclonal Antibody
Ipilimumab
Bintrafusp Alfa
Anti-PD1 Monoclonal Antibody
Tremelimumab
Camrelizumab
Cemiplimab
Sintilimab
Toripalimab
PD-L1 Peptide Vaccine
Interferon-gamma
Envafolimab
Tislelizumab
Retlirafusp Alfa
Drug

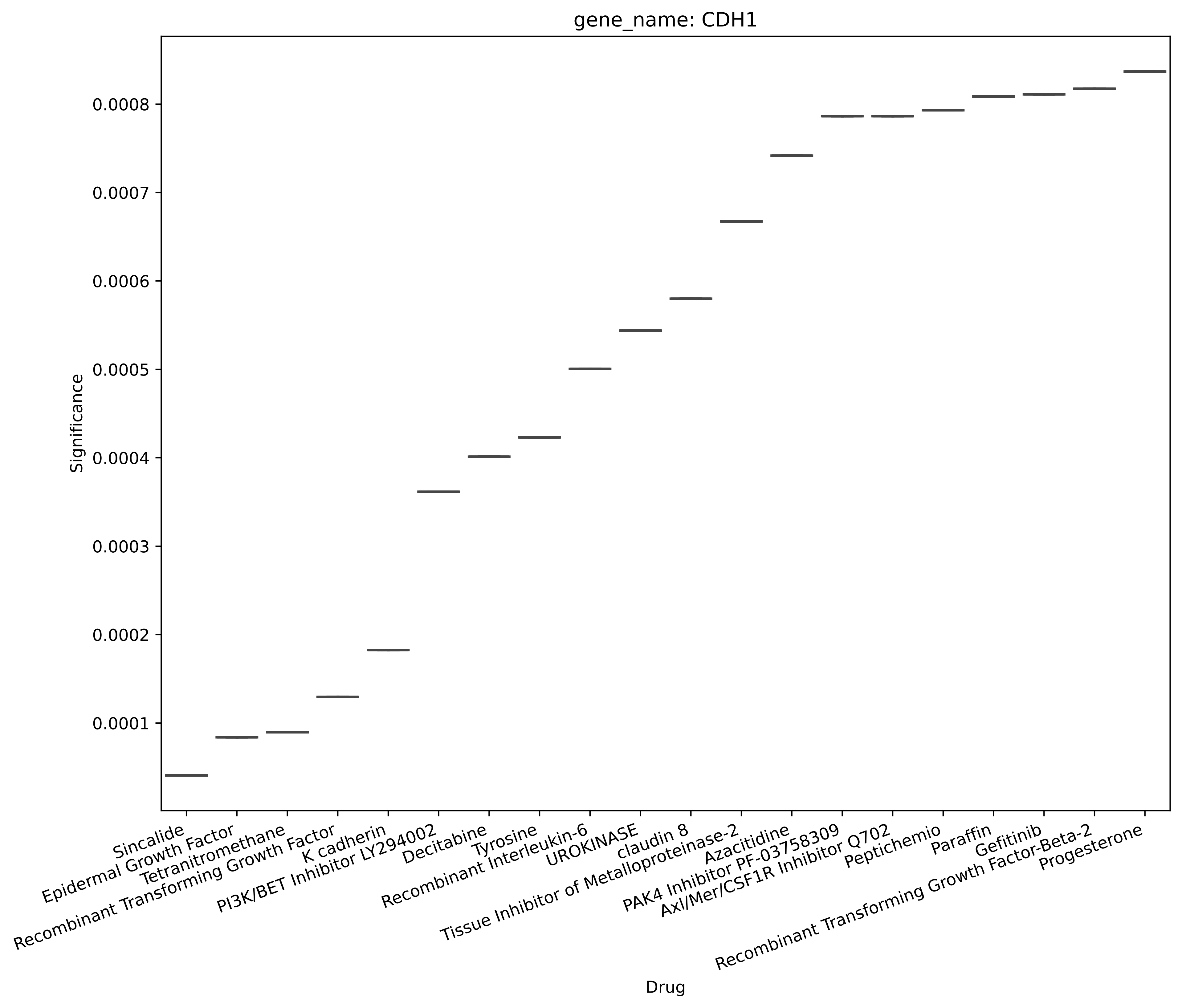

gene_name: CDH1
Significance
0.0008
0.0007
0.0006
0.0005
0.0004
0.0003
0.0002
0.0001
Sincalide
Epidermal Growth Factor
Tetranitromethane
Recombinant Transforming Growth Factor
K cadherin
PI3K/BET Inhibitor LY294002
Decitabine
Tyrosine
Recombinant Interleukin-6
UROKINASE
claudin 8
Tissue Inhibitor of Metalloproteinase-2
Azacitidine
PAK4 Inhibitor PF-03758309
Axl/Mer/CSF1R Inhibitor Q702
Peptichemio
Paraffin
Gefitinib
Recombinant Transforming Growth Factor-Beta-2
Progesterone
Drug

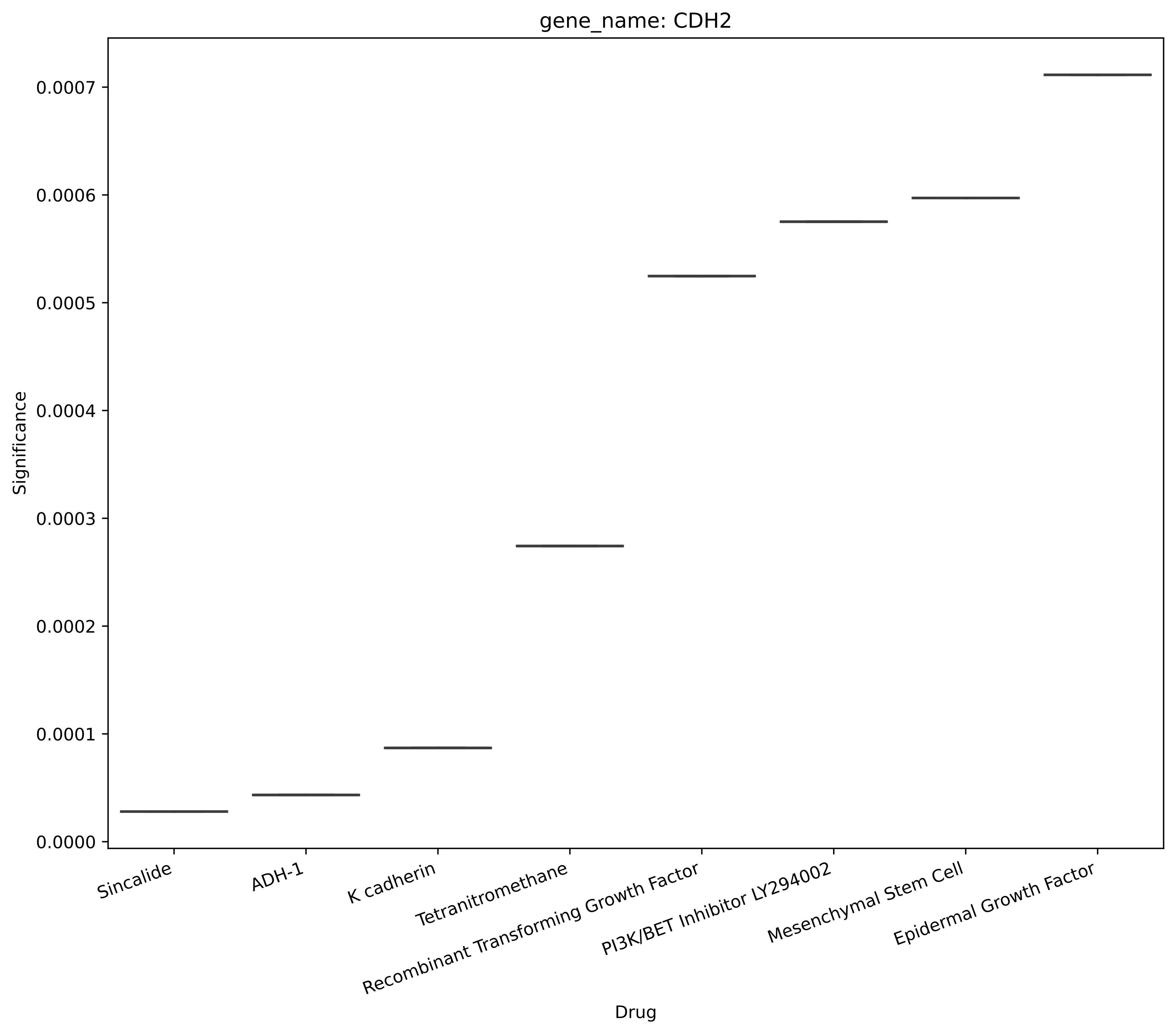
gene_name: CDH2
Significance
0.0000
0.0001
0.0002
0.0003
0.0004
0.0005
0.0006
0.0007
Sincalide
ADH-1
K cadherin
Tetranitromethane
Recombinant Transforming Growth Factor
PI3K/BET Inhibitor LY294002
Mesenchymal Stem Cell
Epidermal Growth Factor
Drug

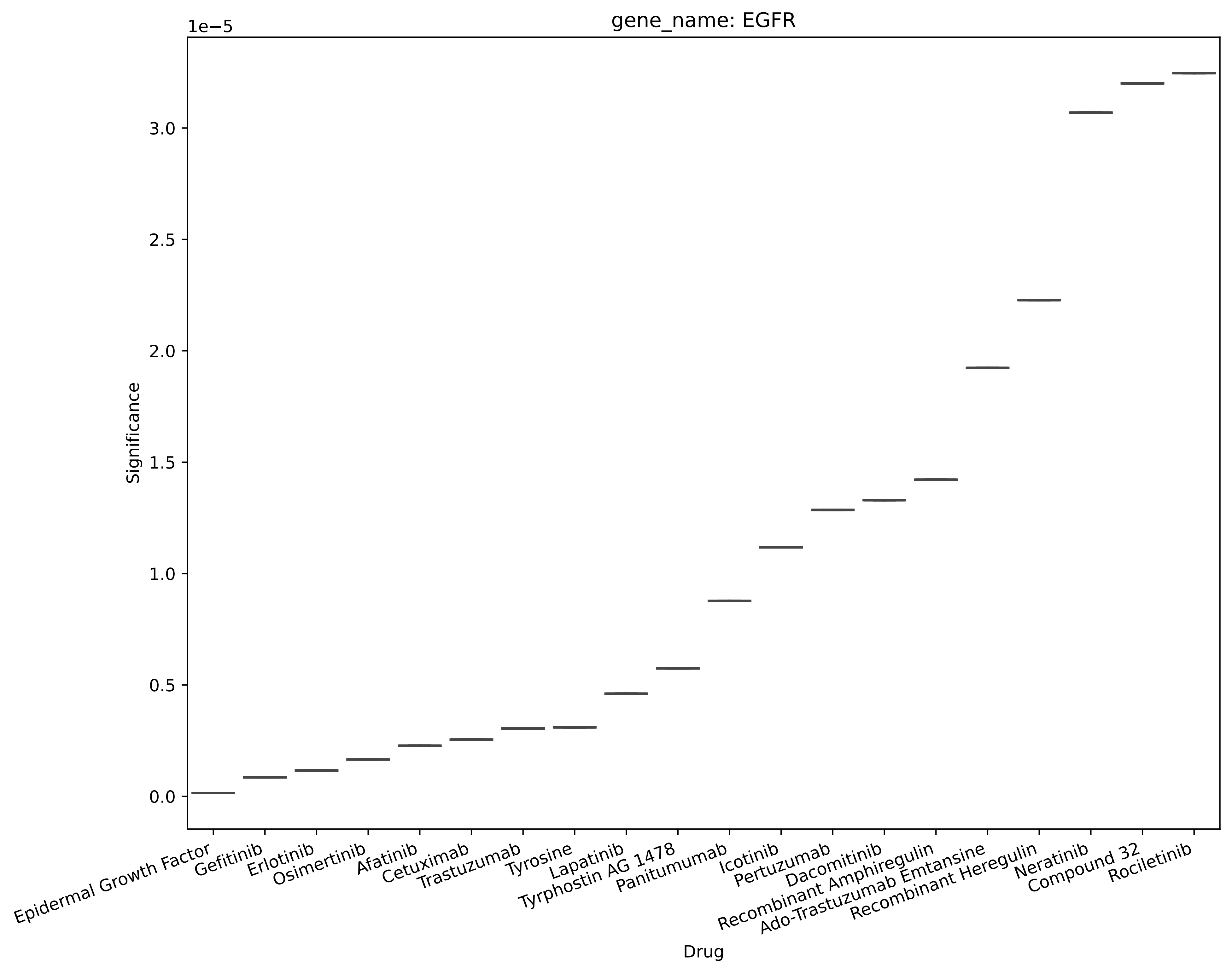
gene_name: EGFR
1e−5
3.0
2.5
2.0
1.5
1.0
0.5
0.0
Significance
Epidermal Growth Factor
Gefitinib
Erlotinib
Osimertinib
Afatinib
Cetuximab
Trastuzumab
Tyrosine
Lapatinib
Tyrphostin AG 1478
Panitumumab
Icotinib
Pertuzumab
Dacomitinib
Recombinant Amphiregulin
Ado-Trastuzumab Emtansine
Recombinant Heregulin
Neratinib
Compound 32
Rociletinib
Drug

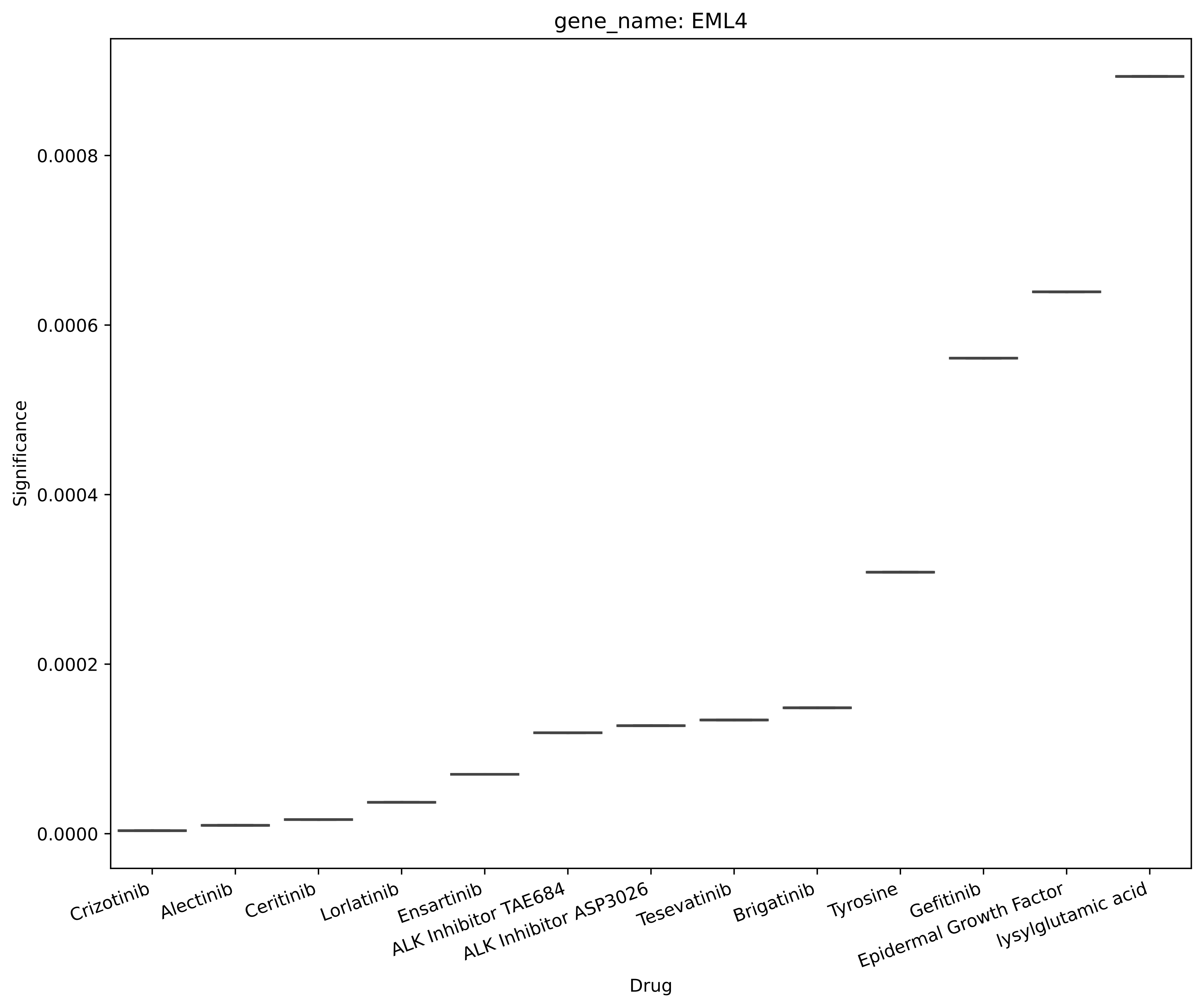
gene_name: EML4
Significance
0.0008
0.0006
0.0004
0.0002
0.0000
Crizotinib
Alectinib
Ceritinib
Lorlatinib
Ensartinib
ALK Inhibitor TAE684
ALK Inhibitor ASP3026
Tesevatinib
Brigatinib
Tyrosine
Gefitinib
Epidermal Growth Factor
lysylglutamic acid
Drug

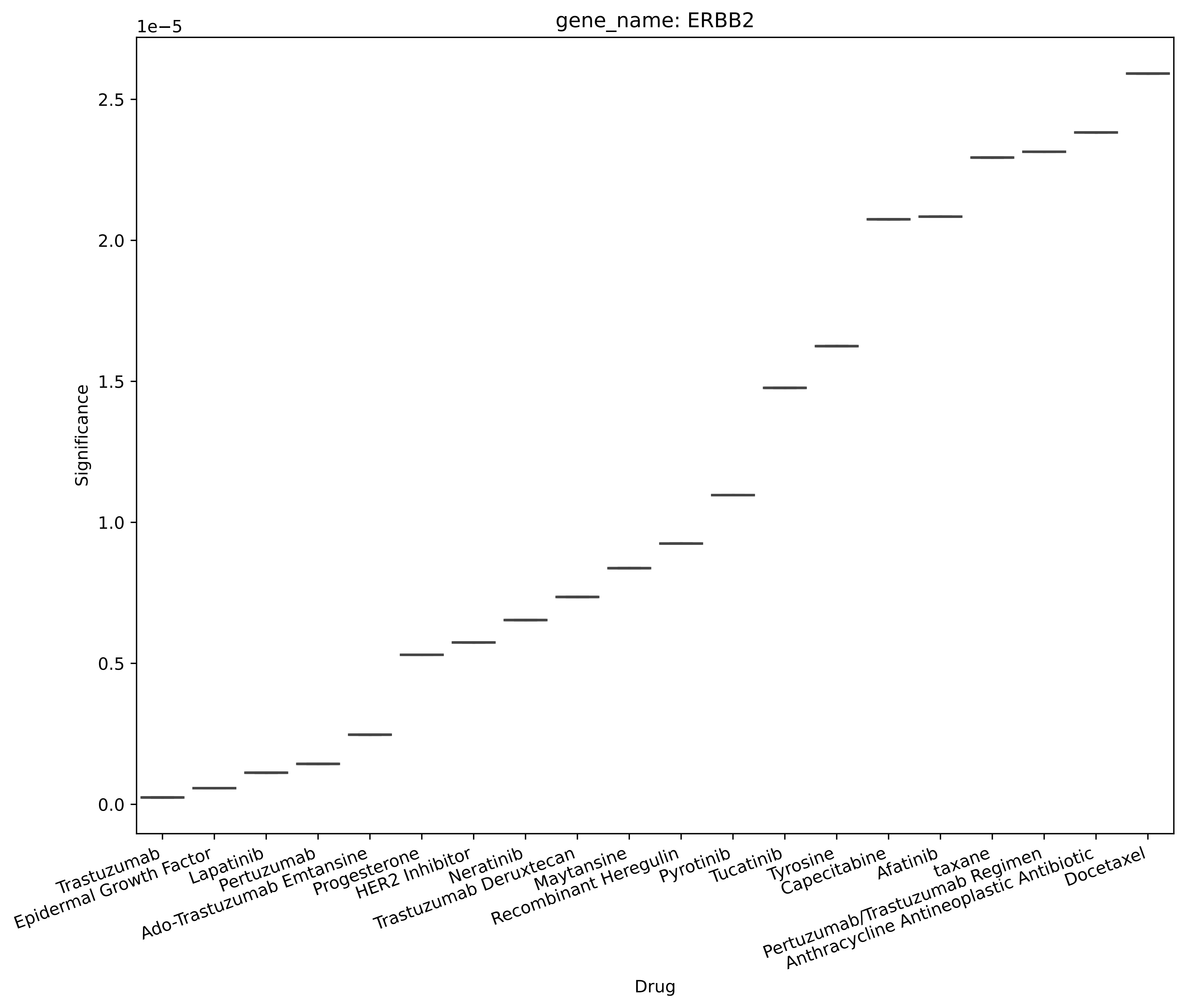

gene_name: ERBB2
1e−5
Significance
0.0
0.5
1.0
1.5
2.0
2.5
Trastuzumab
Epidermal Growth Factor
Lapatinib
Pertuzumab
Ado-Trastuzumab Emtansine
Progesterone
HER2 Inhibitor
Neratinib
Trastuzumab Deruxtecan
Maytansine
Recombinant Heregulin
Pyrotinib
Tucatinib
Tyrosine
Capecitabine
Afatinib
taxane
Pertuzumab/Trastuzumab Regimen
Anthracycline Antineoplastic Antibiotic
Docetaxel
Drug

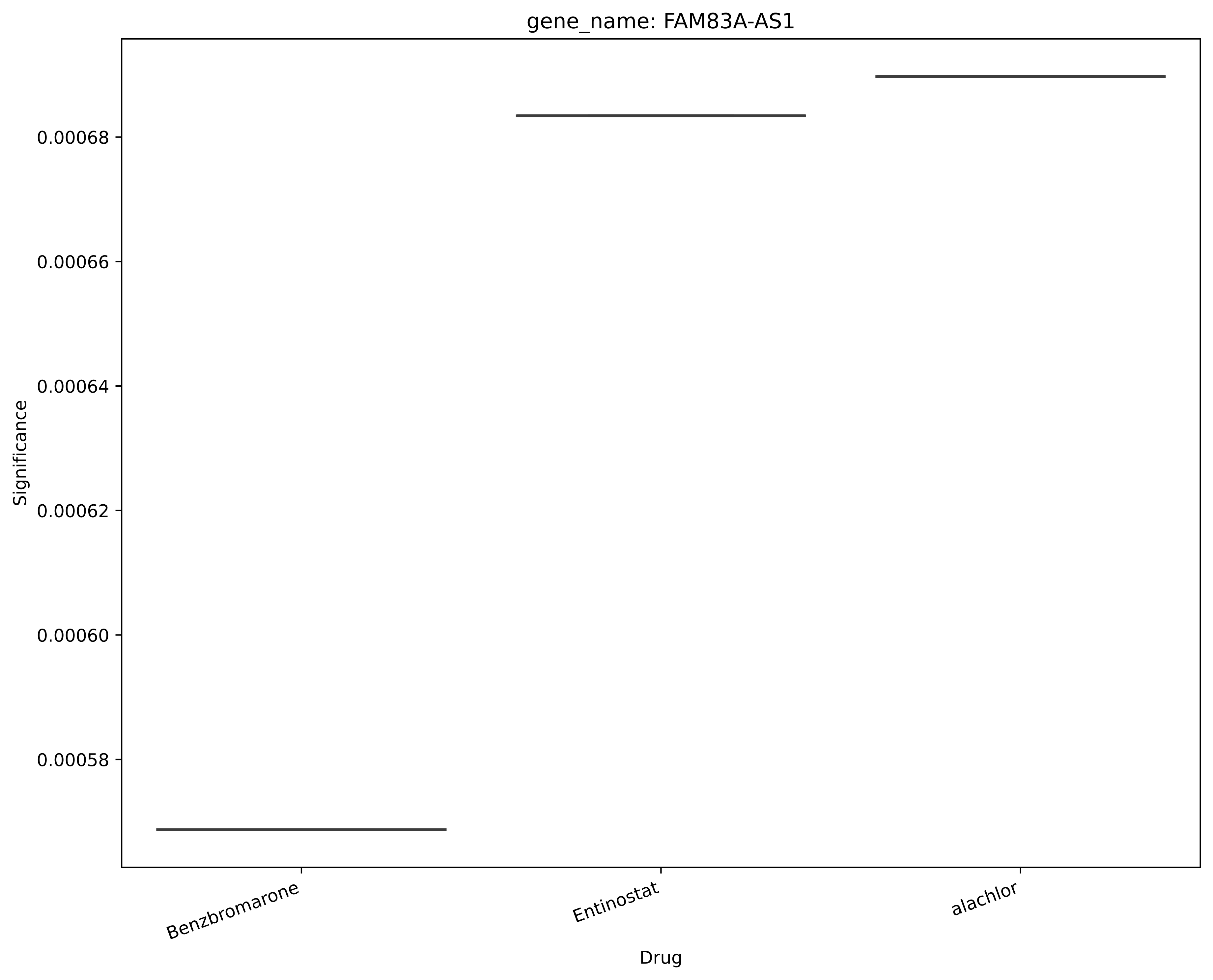
gene_name: FAM83A-AS1
0.00068
0.00066
0.00064
0.00062
0.00060
0.00058
Significance
Benzbromarone
Entinostat
alachlor
Drug

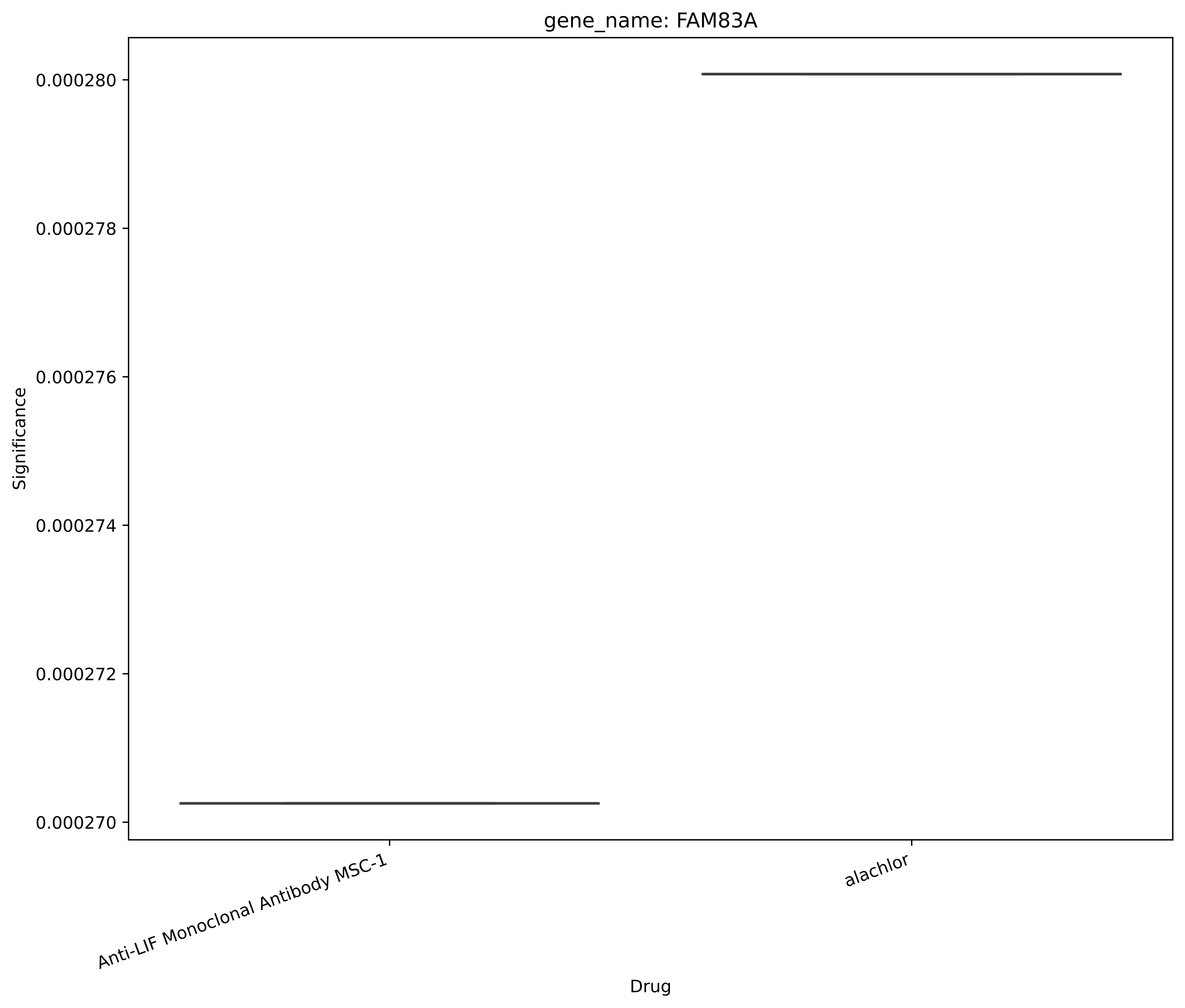

gene_name: FAM83A
Significance
0.000280
0.000278
0.000276
0.000274
0.000272
0.000270
Anti-LIF Monoclonal Antibody MSC-1
alachlor
Drug

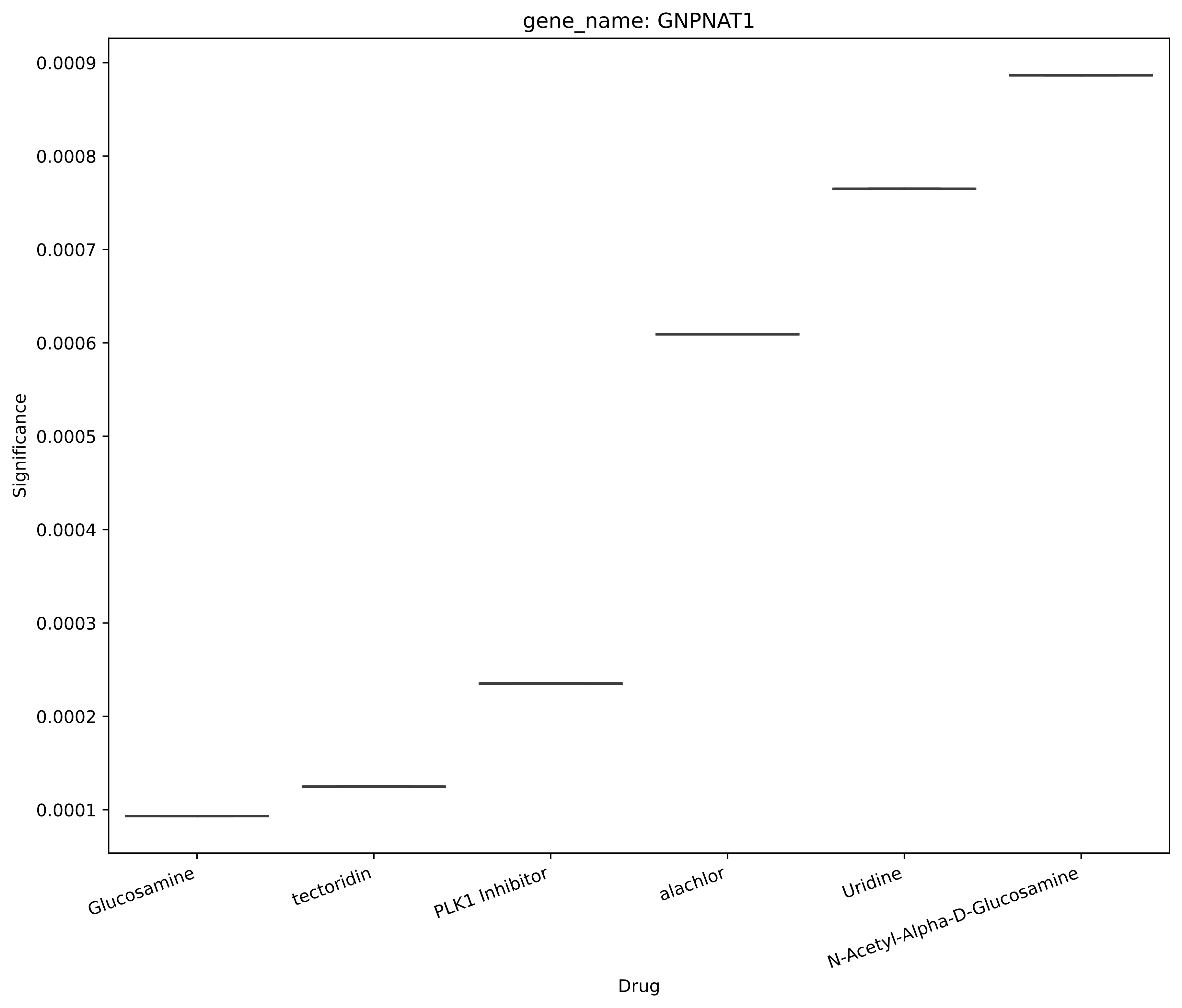
gene_name: GNPNAT1
Significance
0.0009
0.0008
0.0007
0.0006
0.0005
0.0004
0.0003
0.0002
0.0001
Glucosamine
tectoridin
PLK1 Inhibitor
alachlor
Uridine
N-Acetyl-Alpha-D-Glucosamine
Drug

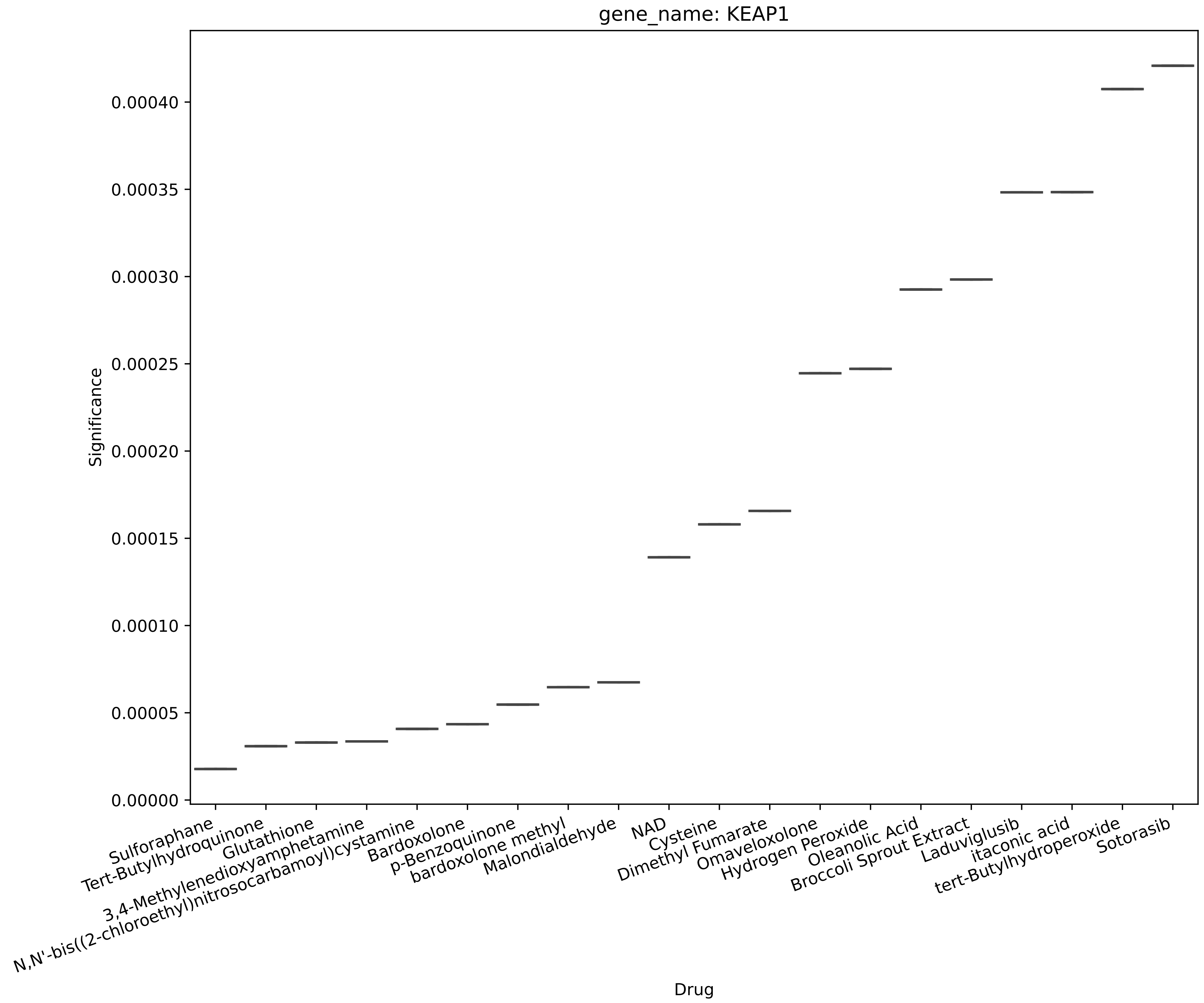

gene_name: KEAP1
Significance
0.00000
0.00005
0.00010
0.00015
0.00020
0.00025
0.00030
0.00035
0.00040
Sulforaphane
Tert-Butylhydroquinone
Glutathione
3,4-Methylenedioxyamphetamine
N,N'-bis((2-chloroethyl)nitrosocarbamoyl)cystamine
Bardoxolone
p-Benzoquinone
bardoxolone methyl
Malondialdehyde
NAD
Cysteine
Dimethyl Fumarate
Omaveloxolone
Hydrogen Peroxide
Oleanolic Acid
Broccoli Sprout Extract
Laduviglusib
itaconic acid
tert-Butylhydroperoxide
Sotorasib
Drug

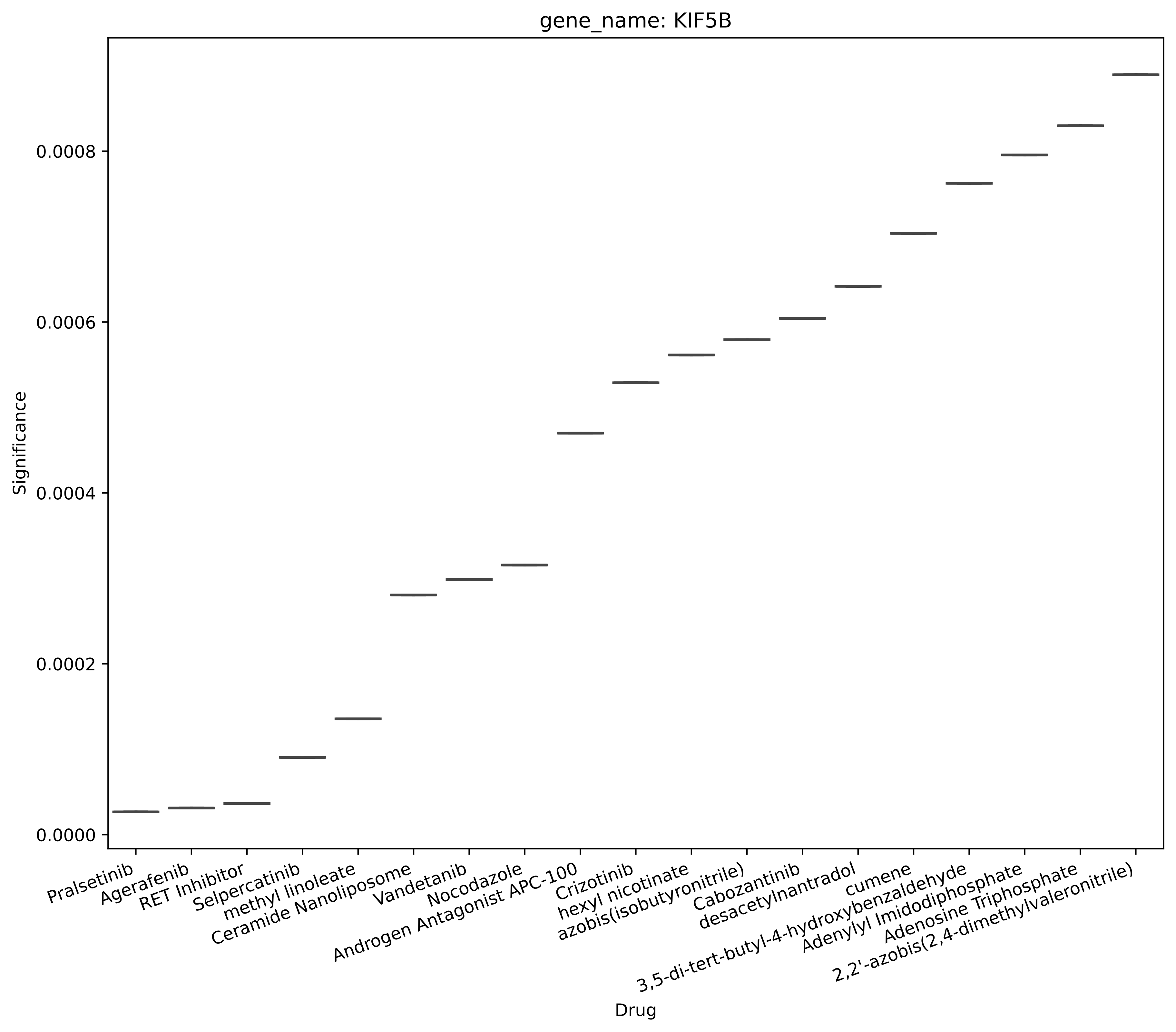

gene_name: KIF5B
Significance
0.0000
0.0002
0.0004
0.0006
0.0008
Drug
Pralsetinib
Agerafenib
RET Inhibitor
Selpercatinib
methyl linoleate
Ceramide Nanoliposome
Vandetanib
Nocodazole
Androgen Antagonist APC-100
Crizotinib
hexyl nicotinate
azobis(isobutyronitrile)
Cabozantinib
desacetylnantradol
3,5-di-tert-butyl-4-hydroxybenzaldehyde
cumene
Adenylyl Imidodiphosphate
Adenosine Triphosphate
2,2'-azobis(2,4-dimethylvaleronitrile)

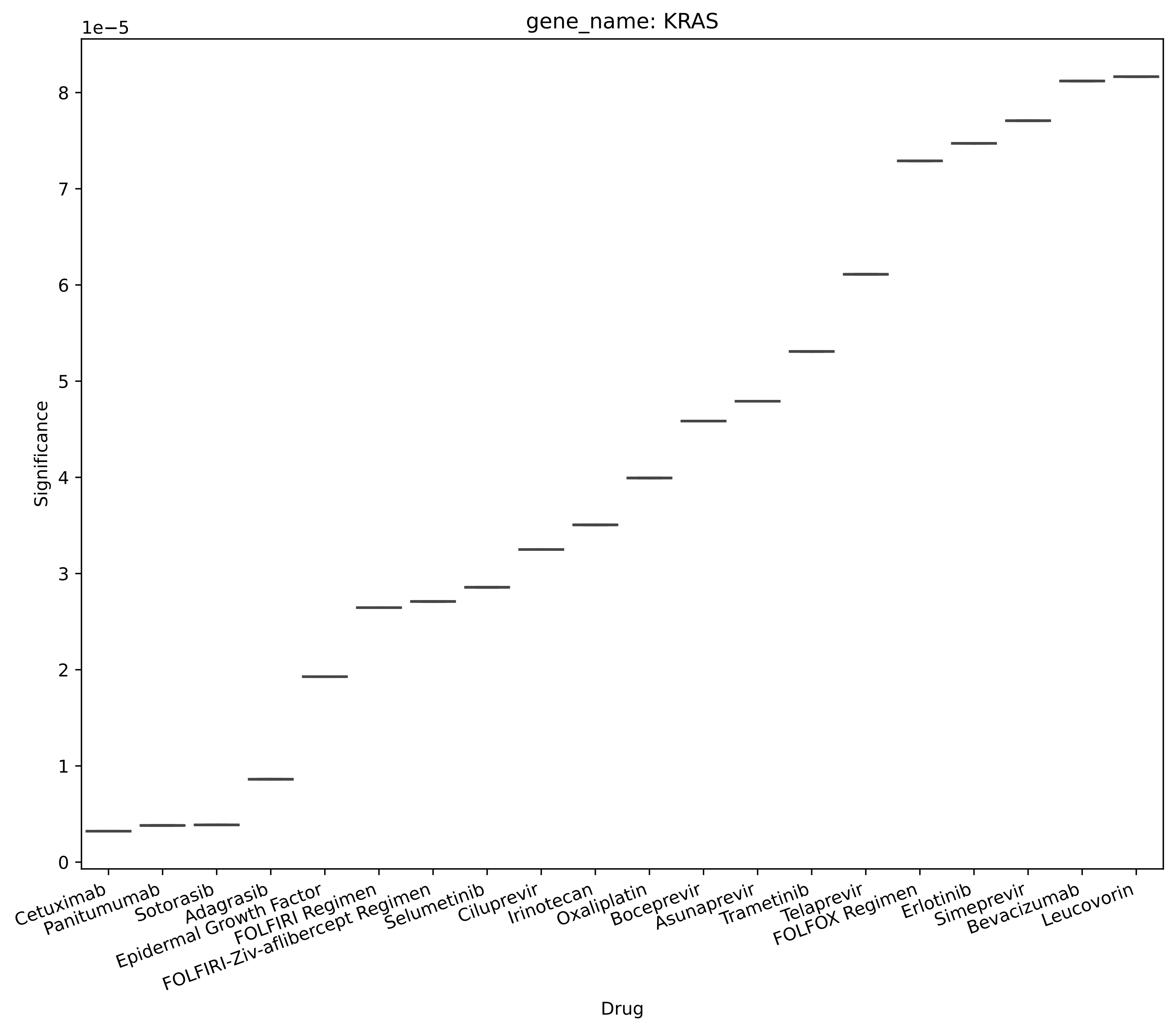

gene_name: KRAS
1e−5
Significance
0
1
2
3
4
5
6
7
8
Cetuximab
Panitumumab
Sotorasib
Adagrasib
Epidermal Growth Factor
FOLFIRI Regimen
FOLFIRI-Ziv-aflibercept Regimen
Selumetinib
Ciluprevir
Irinotecan
Oxaliplatin
Boceprevir
Asunaprevir
Trametinib
Telaprevir
FOLFOX Regimen
Erlotinib
Simeprevir
Bevacizumab
Leucovorin
Drug

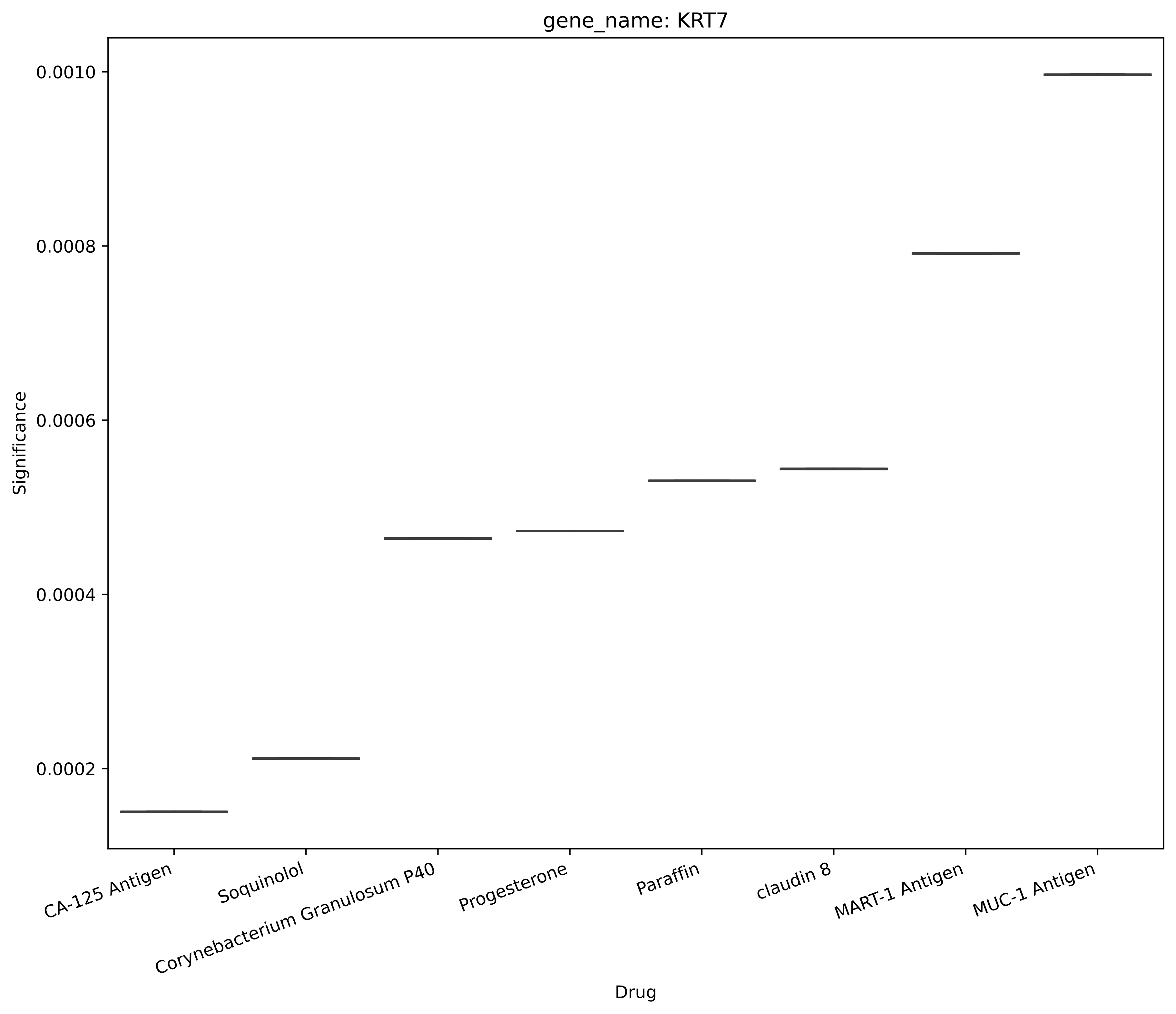
gene_name: KRT7
Significance
0.0010
0.0008
0.0006
0.0004
0.0002
CA-125 Antigen
Soquinolol
Corynebacterium Granulosum P40
Progesterone
Paraffin
claudin 8
MART-1 Antigen
MUC-1 Antigen
Drug

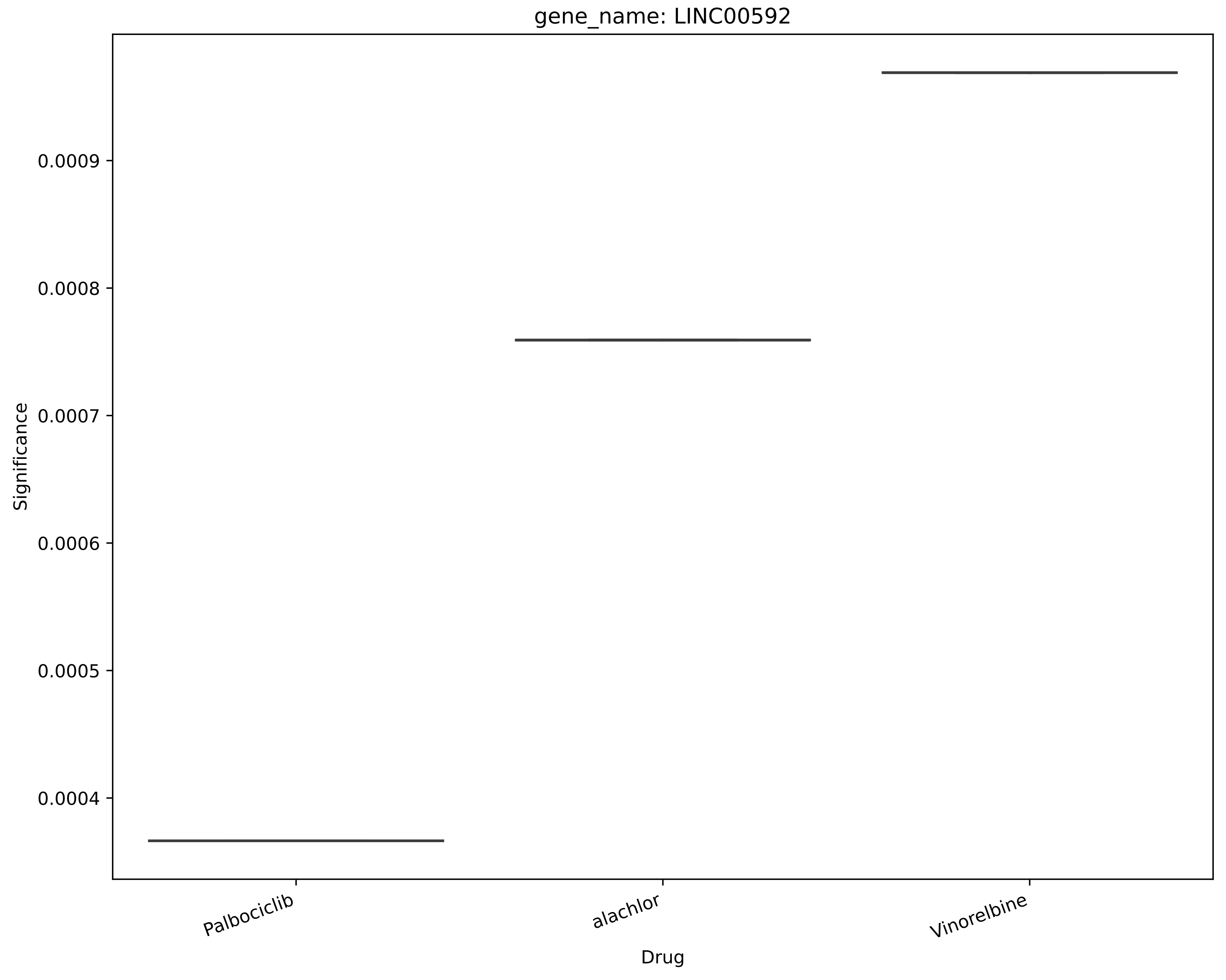
gene_name: LINC00592
Significance
0.0009
0.0008
0.0007
0.0006
0.0005
0.0004
Palbociclib
alachlor
Vinorelbine
Drug

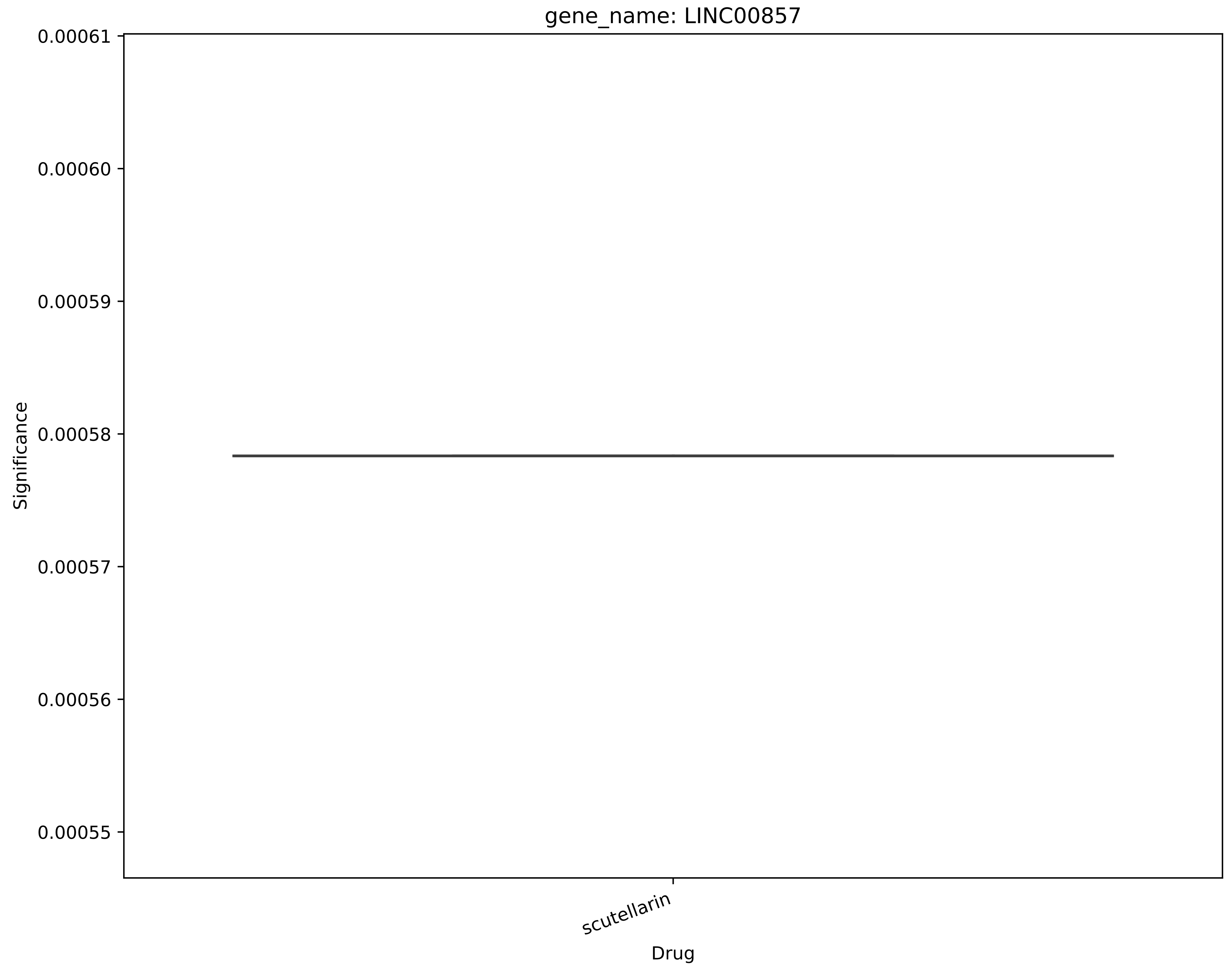
gene_name: LINC00857
0.00061
0.00060
0.00059
0.00058
0.00057
0.00056
0.00055
Significance
scutellarin
Drug

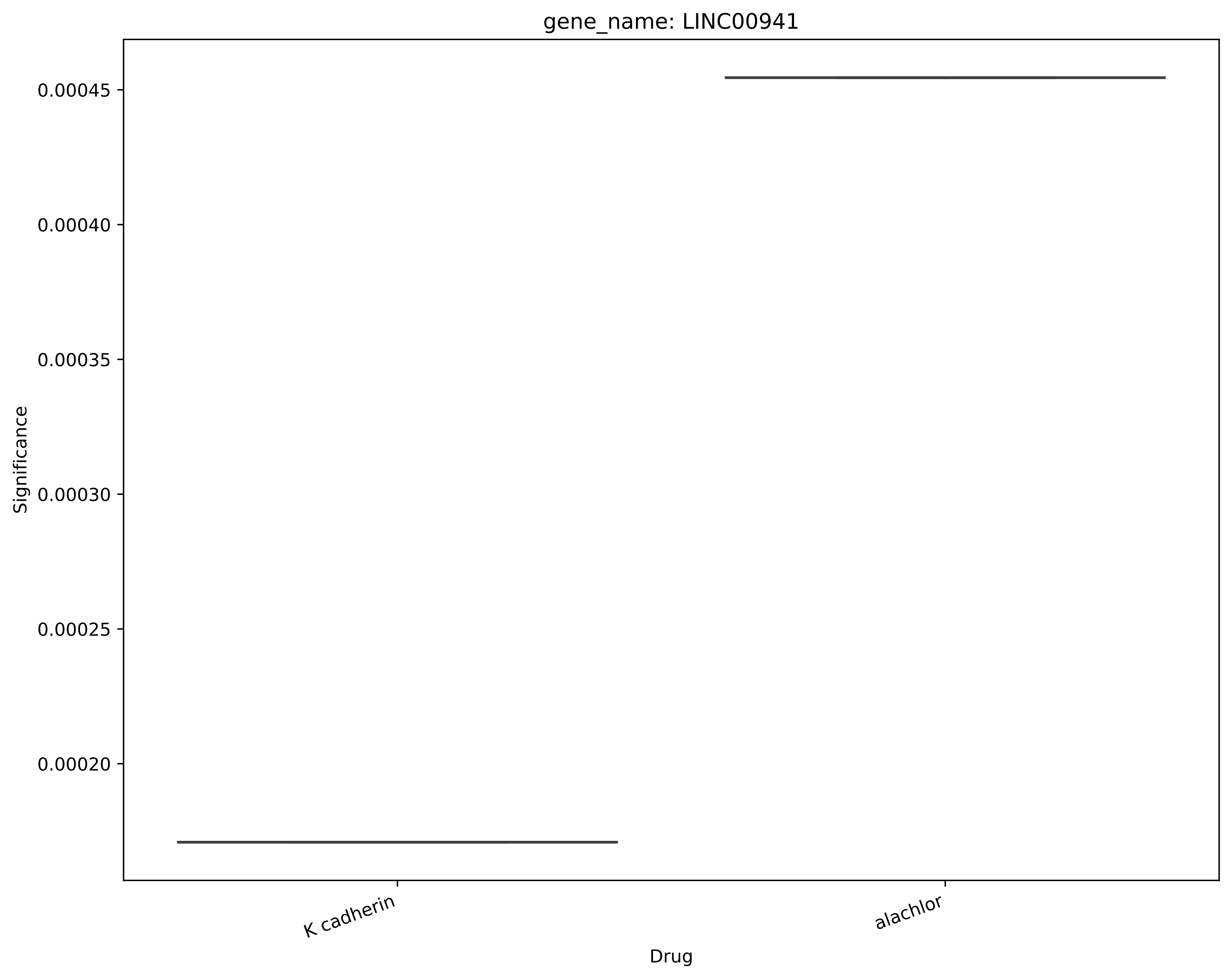
gene_name: LINC00941
0.00045
0.00040
0.00035
0.00030
0.00025
0.00020
Significance
K cadherin
alachlor
Drug

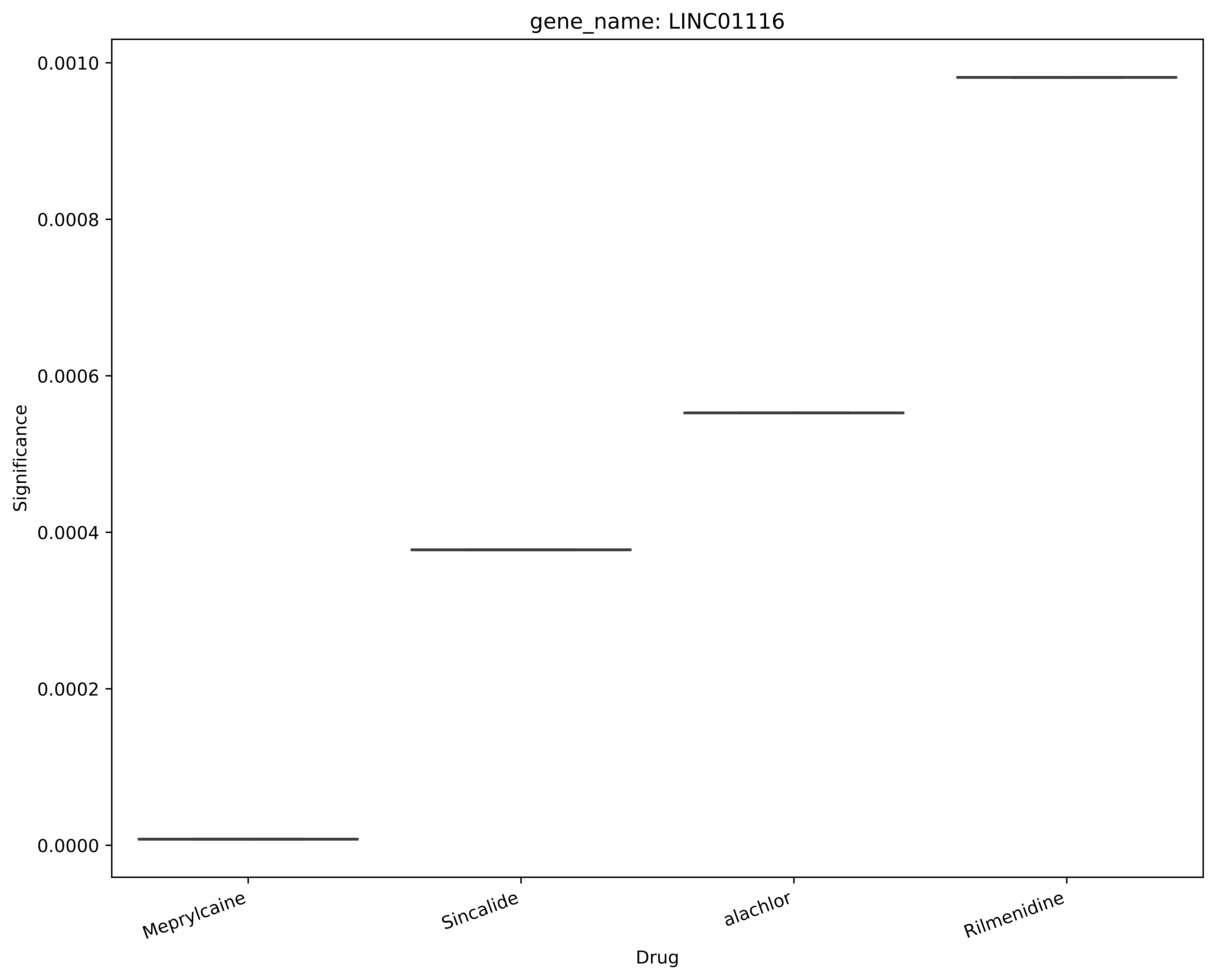

gene_name: LINC01116
Significance
0.0010
0.0008
0.0006
0.0004
0.0002
0.0000
Meprylcaine
Sincalide
alachlor
Rilmenidine
Drug

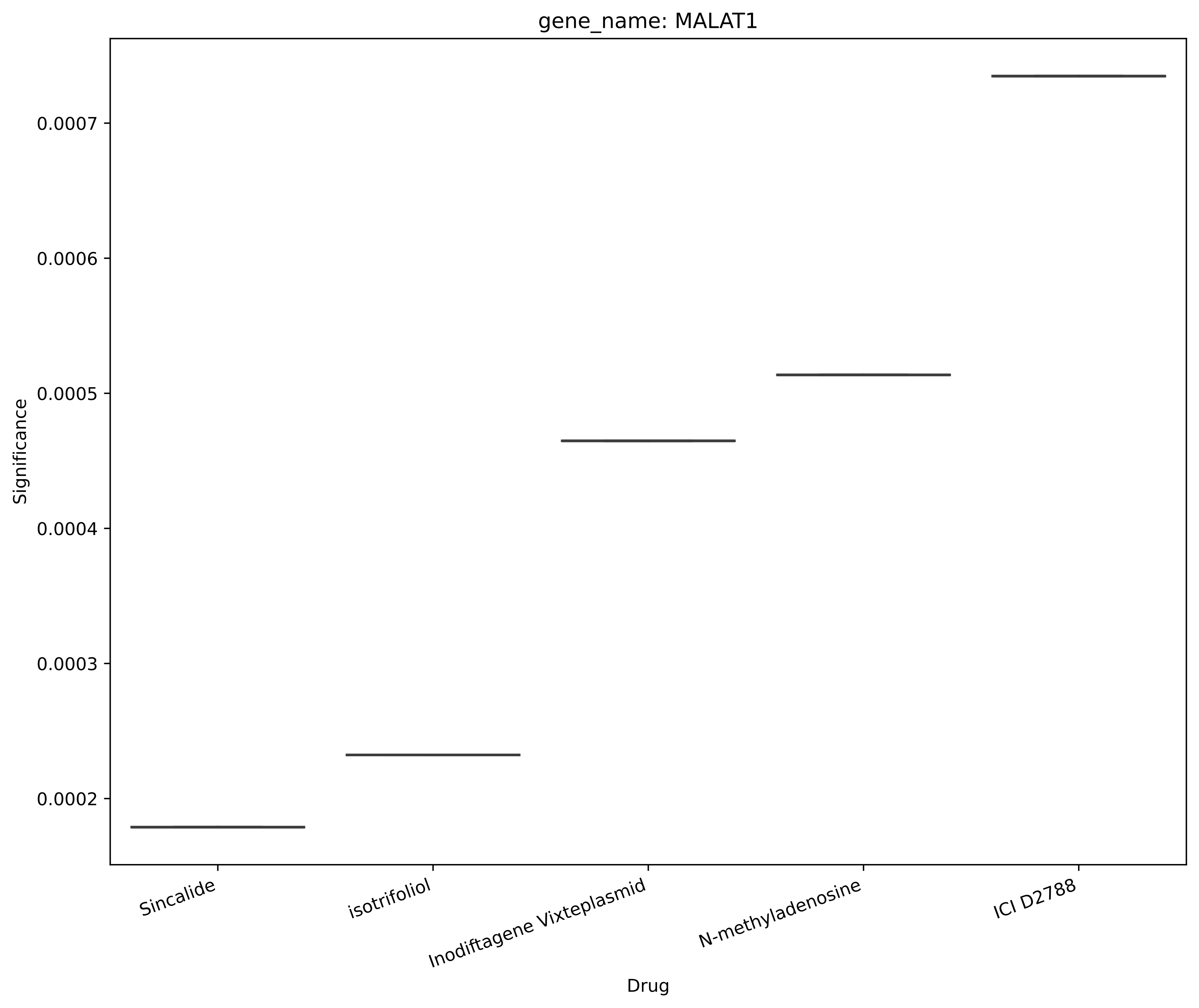
gene_name: MALAT1
Significance
0.0002
0.0003
0.0004
0.0005
0.0006
0.0007
Sincalide
isotrifoliol
Inodiftagene Vixteplasmid
N-methyladenosine
ICI D2788
Drug

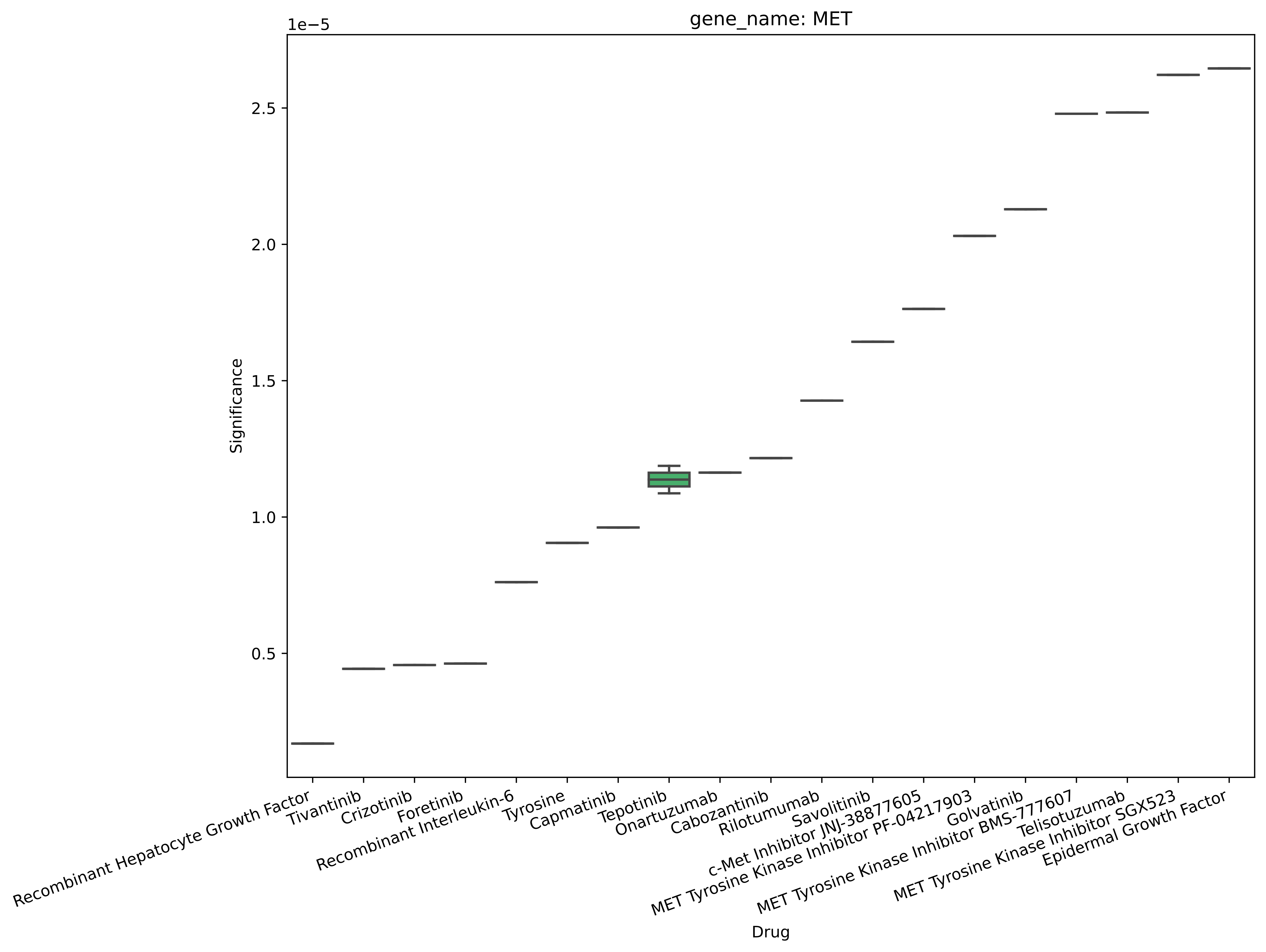

gene_name: MET
1e−5
Significance
2.5
2.0
1.5
1.0
0.5
Drug
Recombinant Hepatocyte Growth Factor
Tivantinib
Crizotinib
Foretinib
Recombinant Interleukin-6
Tyrosine
Capmatinib
Tepotinib
Onartuzumab
Cabozantinib
Rilotumumab
Savolitinib
c-Met Inhibitor JNJ-38877605
MET Tyrosine Kinase Inhibitor PF-04217903
Golvatinib
MET Tyrosine Kinase Inhibitor BMS-777607
Telisotuzumab
MET Tyrosine Kinase Inhibitor SGX523
Epidermal Growth Factor

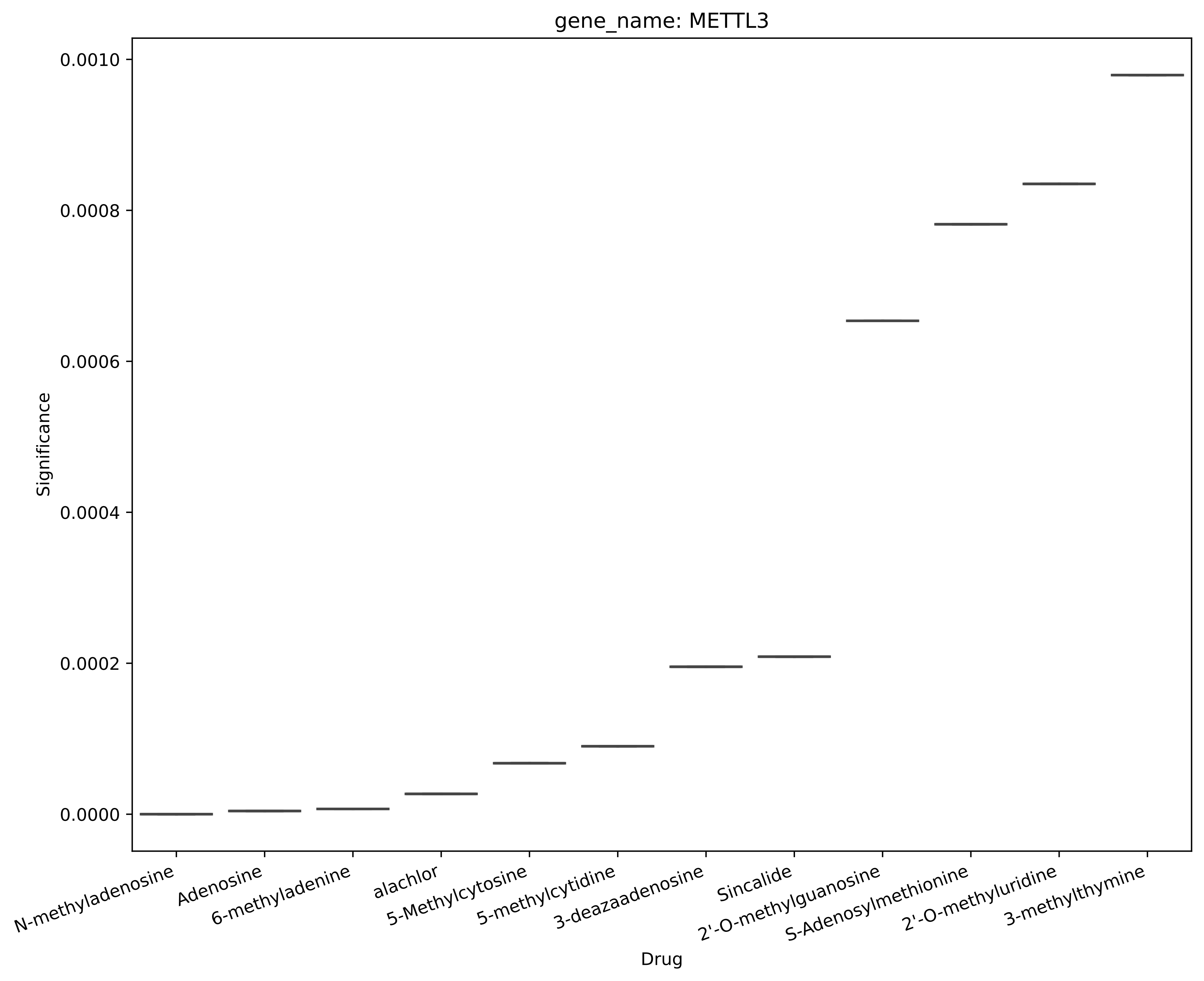

gene_name: METTL3
Significance
0.0010
0.0008
0.0006
0.0004
0.0002
0.0000
N-methyladenosine
Adenosine
6-methyladenine
alachlor
5-Methylcytosine
5-methylcytidine
3-deazaadenosine
Sincalide
2'-O-methylguanosine
S-Adenosylmethionine
2'-O-methyluridine
3-methylthymine
Drug

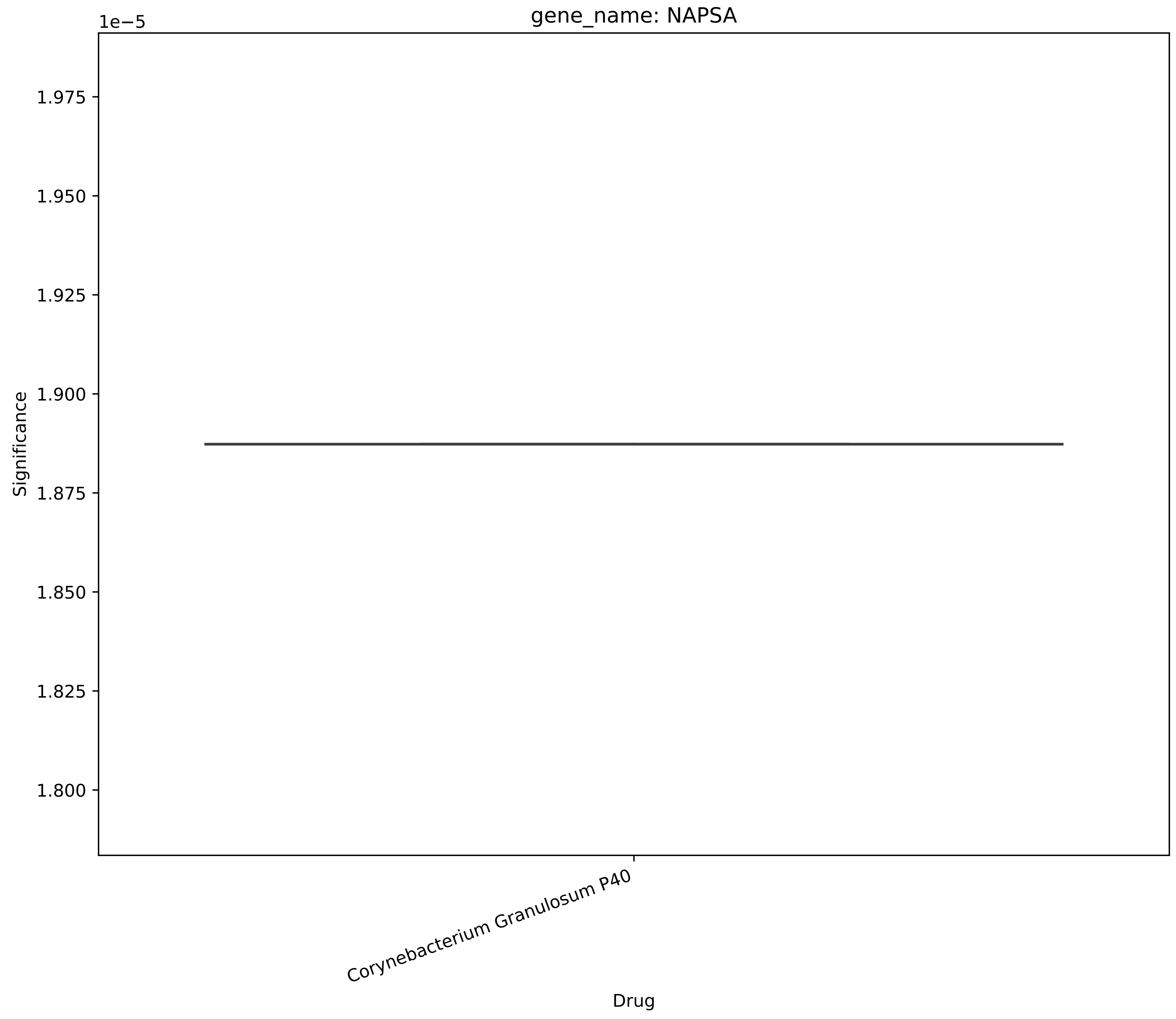
gene_name: NAPSA
1e−5
1.975
1.950
1.925
1.900
1.875
1.850
1.825
1.800
Significance
Corynebacterium Granulosum P40
Drug

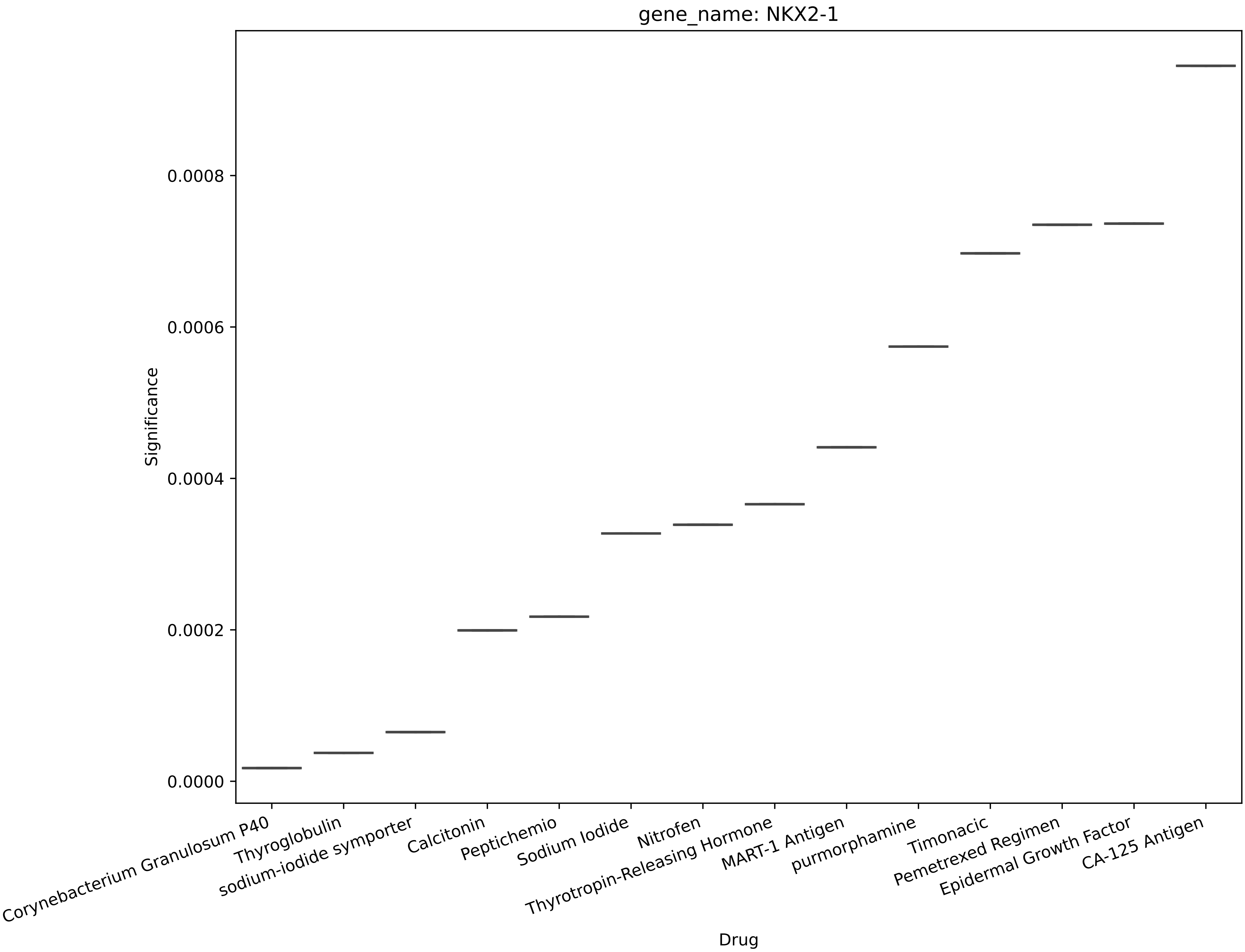
gene_name: NKX2-1
Significance
0.0008
0.0006
0.0004
0.0002
0.0000
Corynebacterium Granulosum P40
Thyroglobulin
sodium-iodide symporter
Calcitonin
Peptichemio
Sodium Iodide
Nitrofen
Thyrotropin-Releasing Hormone
MART-1 Antigen
purmorphamine
Timonacic
Pemetrexed Regimen
Epidermal Growth Factor
CA-125 Antigen
Drug

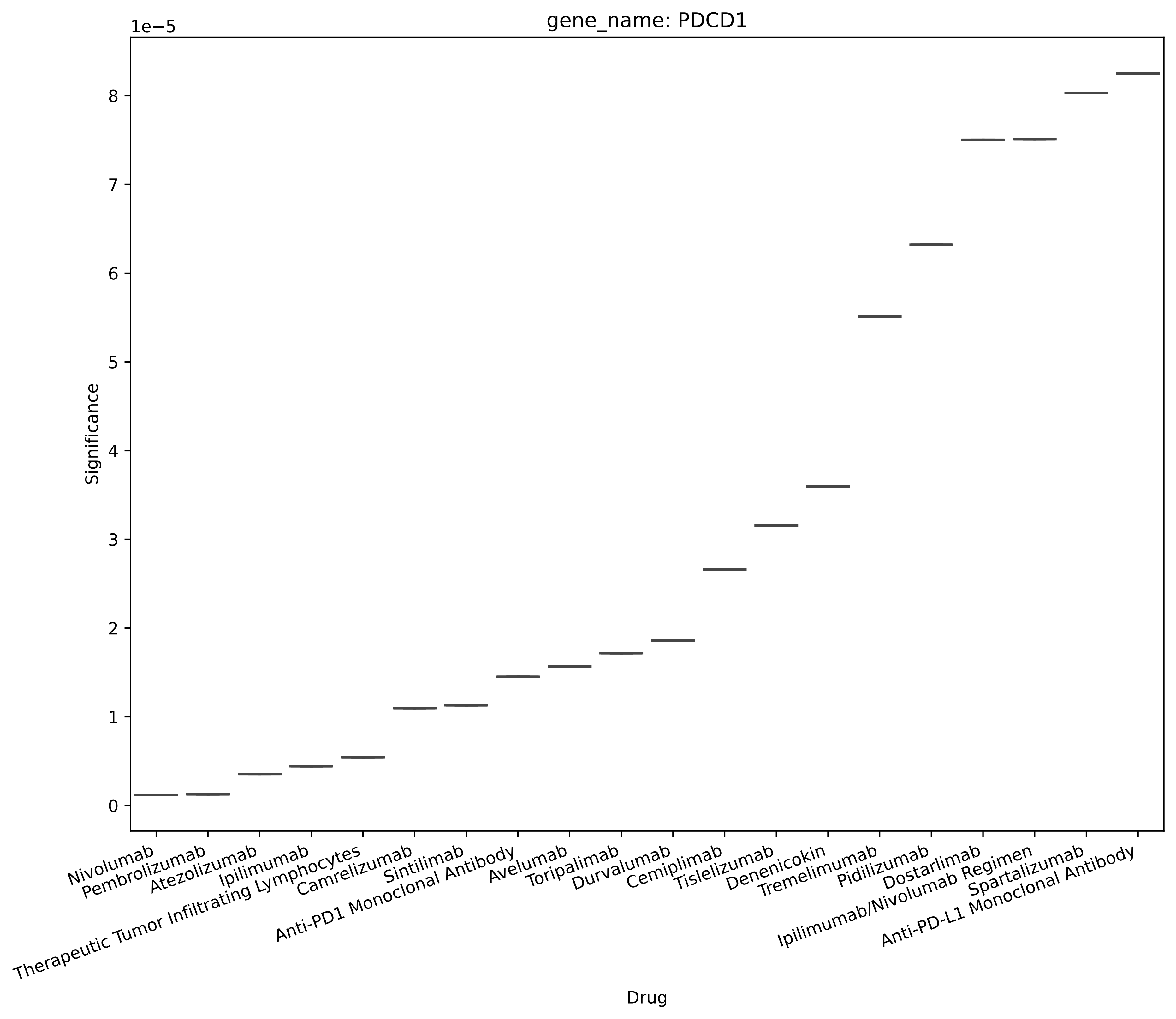
gene_name: PDCD1
1e−5
Significance
0
1
2
3
4
5
6
7
8
Drug
Nivolumab
Pembrolizumab
Atezolizumab
Ipilimumab
Therapeutic Tumor Infiltrating Lymphocytes
Camrelizumab
Sintilimab
Anti-PD1 Monoclonal Antibody
Avelumab
Toripalimab
Durvalumab
Cemiplimab
Tislelizumab
Denenicokin
Tremelimumab
Pidilizumab
Dostarlimab
Ipilimumab/Nivolumab Regimen
Spartalizumab
Anti-PD-L1 Monoclonal Antibody

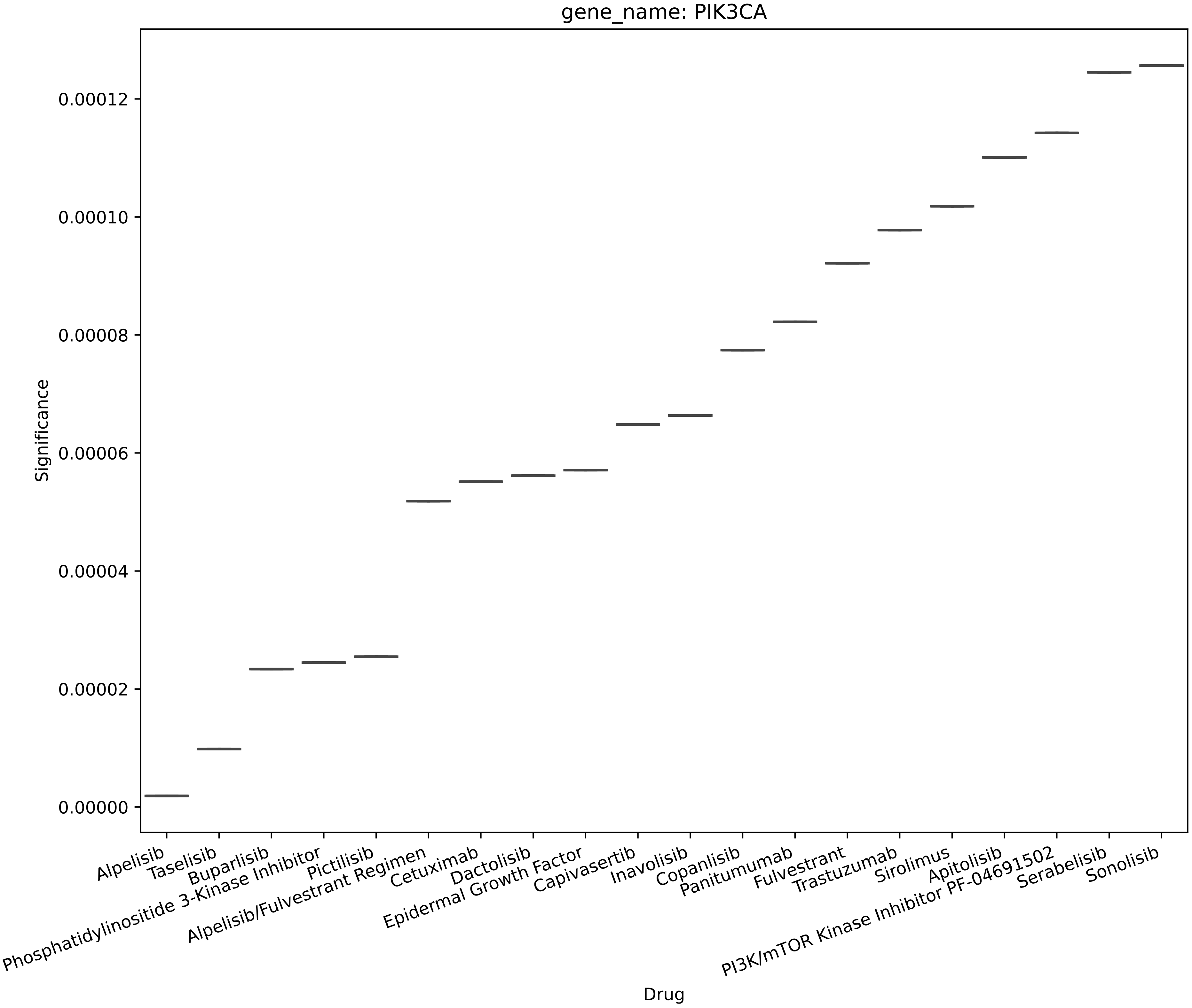
gene_name: PIK3CA
Significance
0.00000
0.00002
0.00004
0.00006
0.00008
0.00010
0.00012
Drug
Alpelisib
Taselisib
Buparlisib
Phosphatidylinositide 3-Kinase Inhibitor
Pictilisib
Alpelisib/Fulvestrant Regimen
Cetuximab
Dactolisib
Epidermal Growth Factor
Capivasertib
Inavolisib
Copanlisib
Panitumumab
Fulvestrant
Trastuzumab
Sirolimus
Apitolisib
PI3K/mTOR Kinase Inhibitor PF-04691502
Serabelisib
Sonolisib

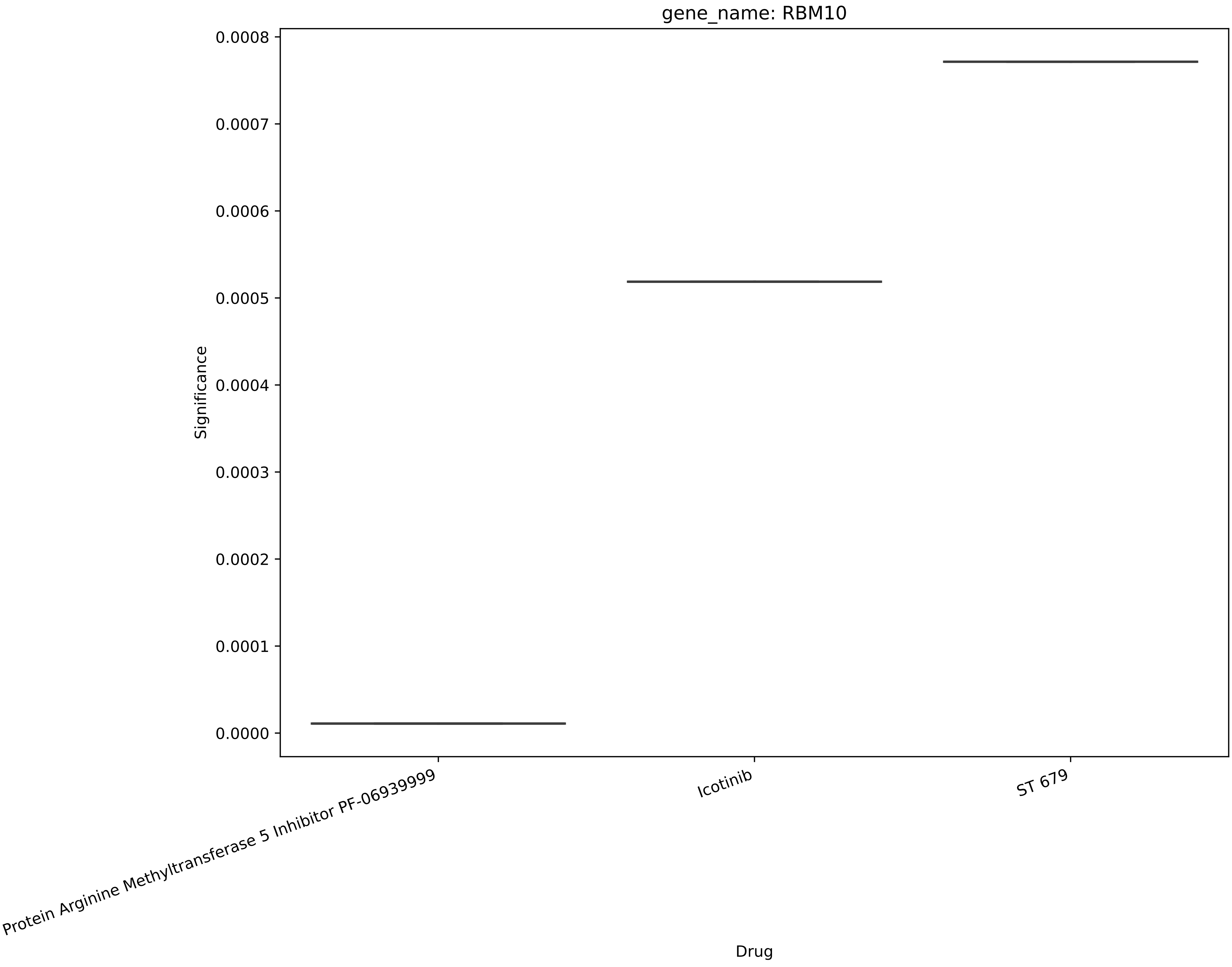
gene_name: RBM10
Significance
0.0008
0.0007
0.0006
0.0005
0.0004
0.0003
0.0002
0.0001
0.0000
Protein Arginine Methyltransferase 5 Inhibitor PF-06939999
Icotinib
ST 679
Drug

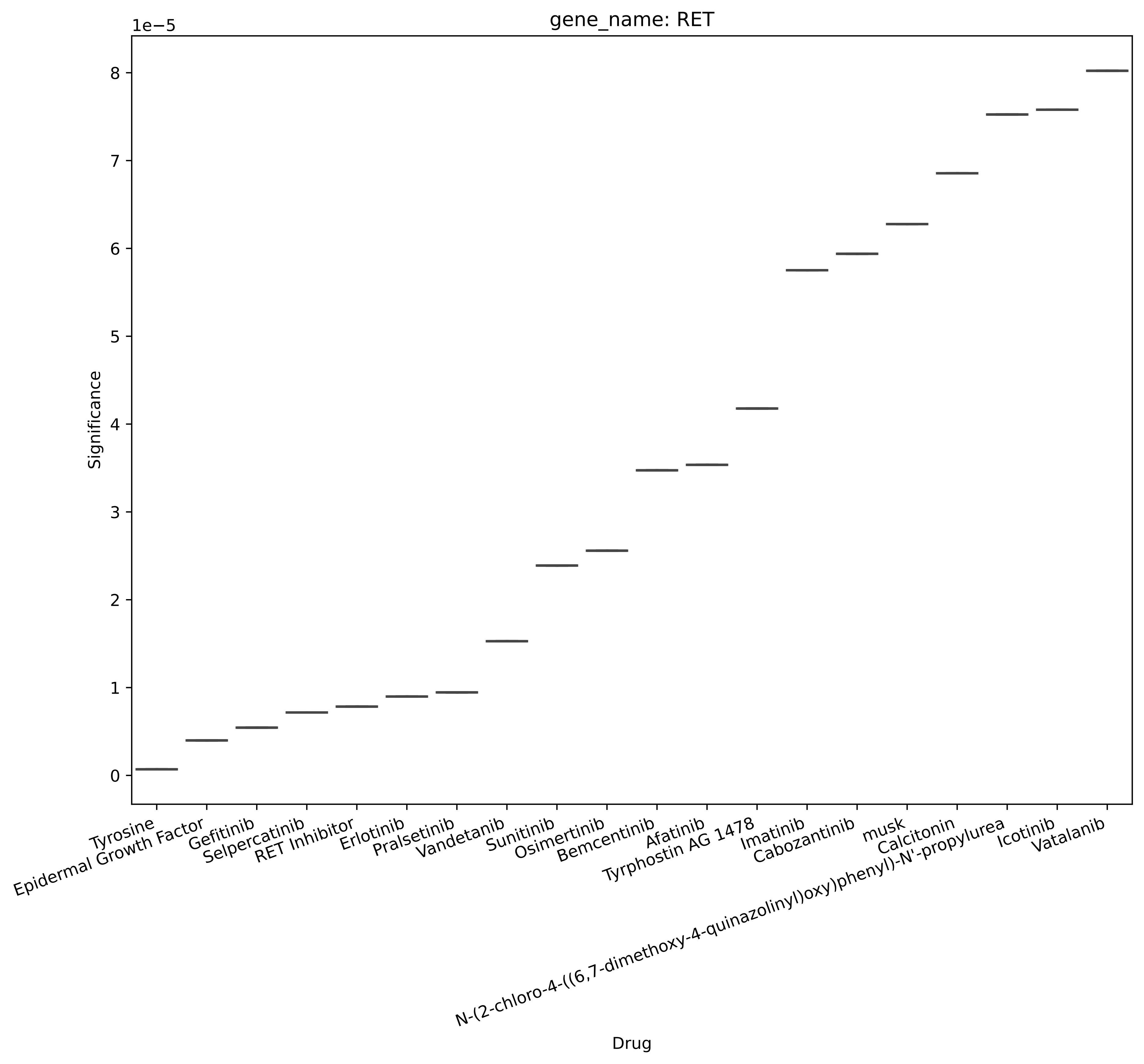

gene_name: RET
1e−5
Significance
0
1
2
3
4
5
6
7
8
Tyrosine
Epidermal Growth Factor
Gefitinib
Selpercatinib
RET Inhibitor
Erlotinib
Pralsetinib
Vandetanib
Sunitinib
Osimertinib
Bemcentinib
Afatinib
Tyrphostin AG 1478
Imatinib
Cabozantinib
musk
Calcitonin
N-(2-chloro-4-((6,7-dimethoxy-4-quinazolinyl)oxy)phenyl)-N'-propylurea
Icotinib
Vatalanib
Drug

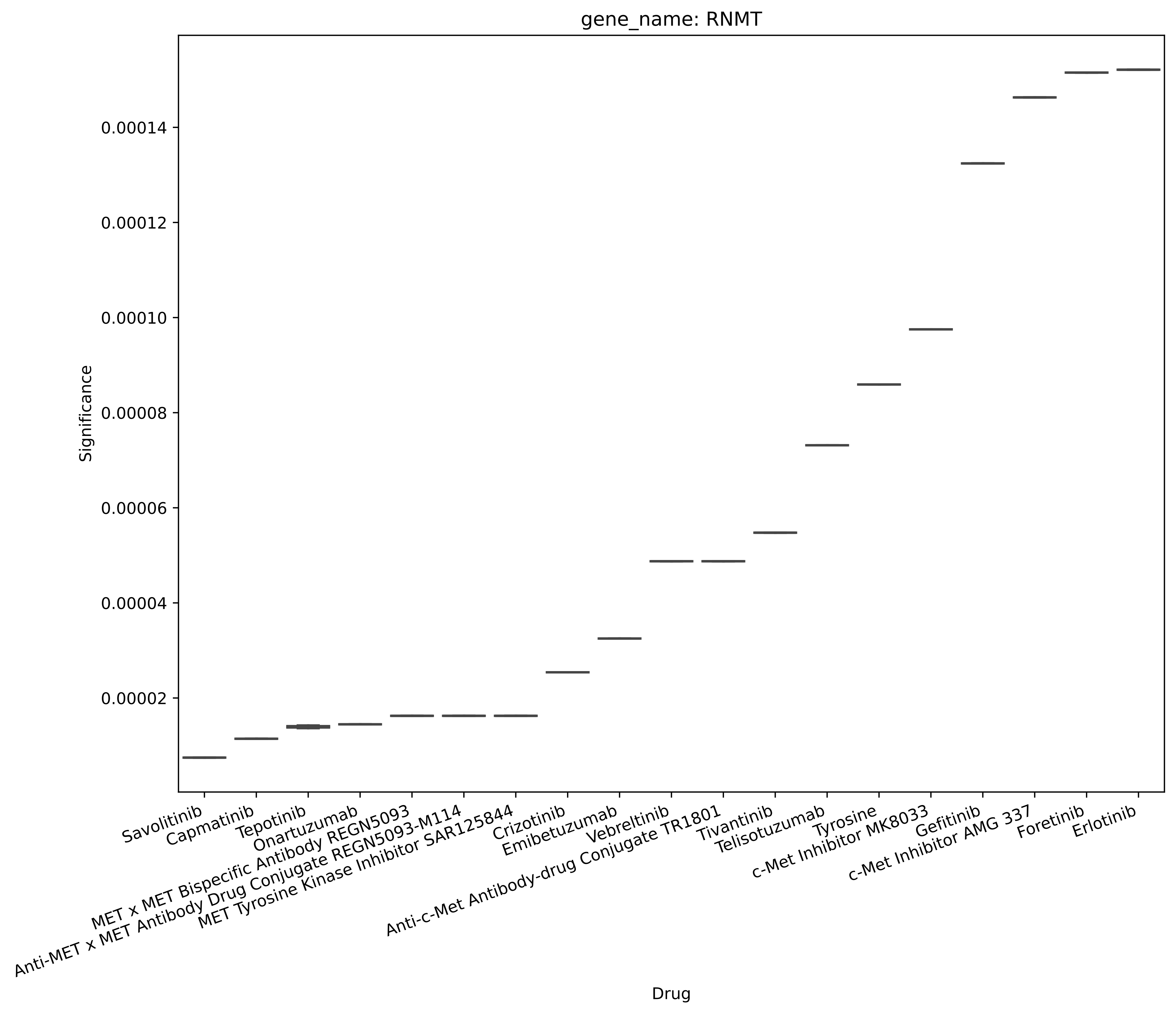

gene_name: RNMT
Significance
0.00014
0.00012
0.00010
0.00008
0.00006
0.00004
0.00002
Drug
Savolitinib
Capmatinib
Tepotinib
Onartuzumab
MET x MET Bispecific Antibody REGN5093
Anti-MET x MET Antibody Drug Conjugate REGN5093-M114
MET Tyrosine Kinase Inhibitor SAR125844
Crizotinib
Emibetuzumab
Vebreltinib
Anti-c-Met Antibody-drug Conjugate TR1801
Tivantinib
Telisotuzumab
Tyrosine
c-Met Inhibitor MK8033
Gefitinib
c-Met Inhibitor AMG 337
Foretinib
Erlotinib

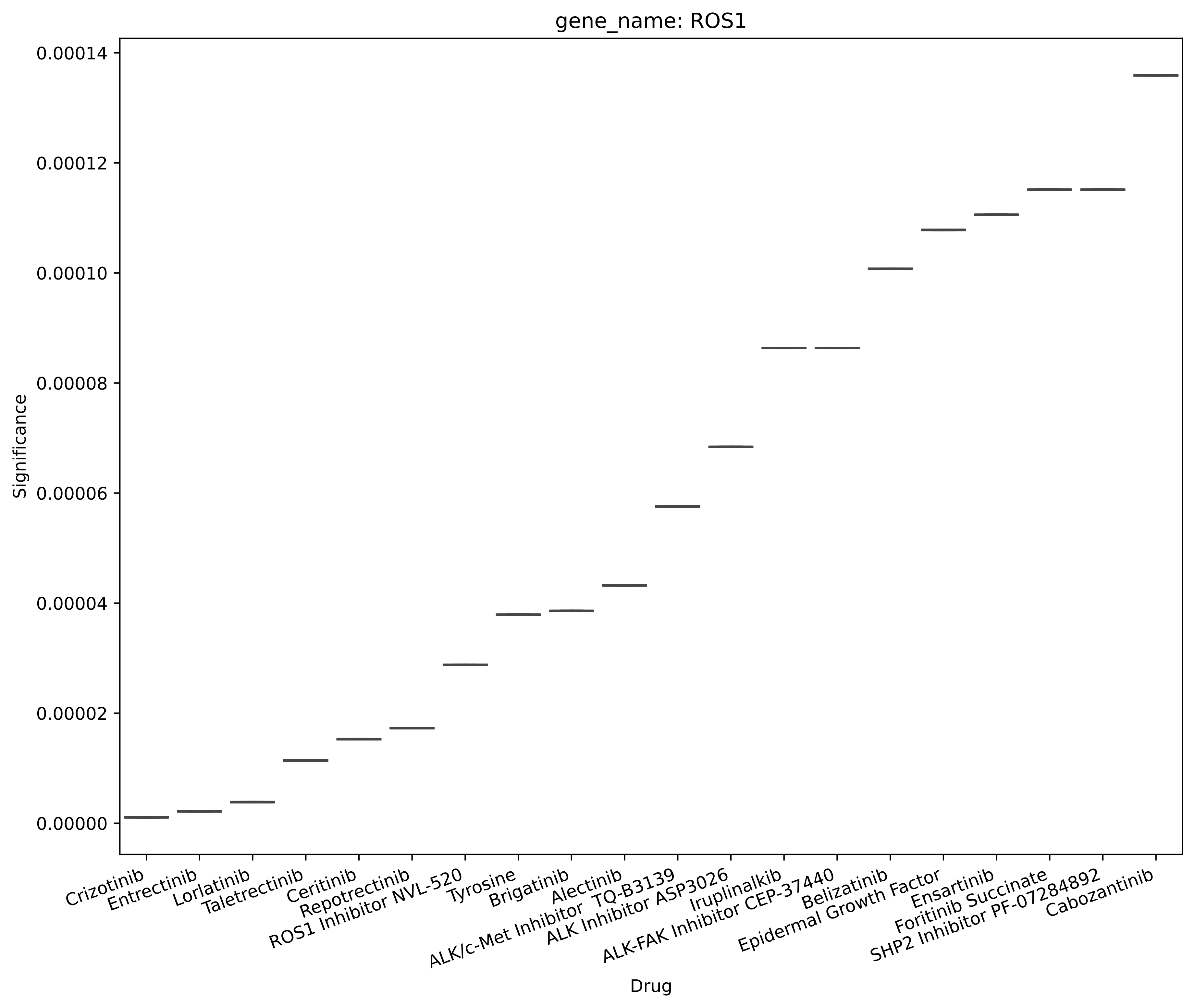

gene_name: ROS1
Significance
0.00014
0.00012
0.00010
0.00008
0.00006
0.00004
0.00002
0.00000
Crizotinib
Entrectinib
Lorlatinib
Taletrectinib
Ceritinib
Repotrectinib
ROS1 Inhibitor NVL-520
Tyrosine
Brigatinib
Alectinib
ALK/c-Met Inhibitor TQ-B3139
ALK Inhibitor ASP3026
Iruplinalkib
ALK-FAK Inhibitor CEP-37440
Belizatinib
Epidermal Growth Factor
Ensartinib
Foritinib Succinate
SHP2 Inhibitor PF-07284892
Cabozantinib
Drug

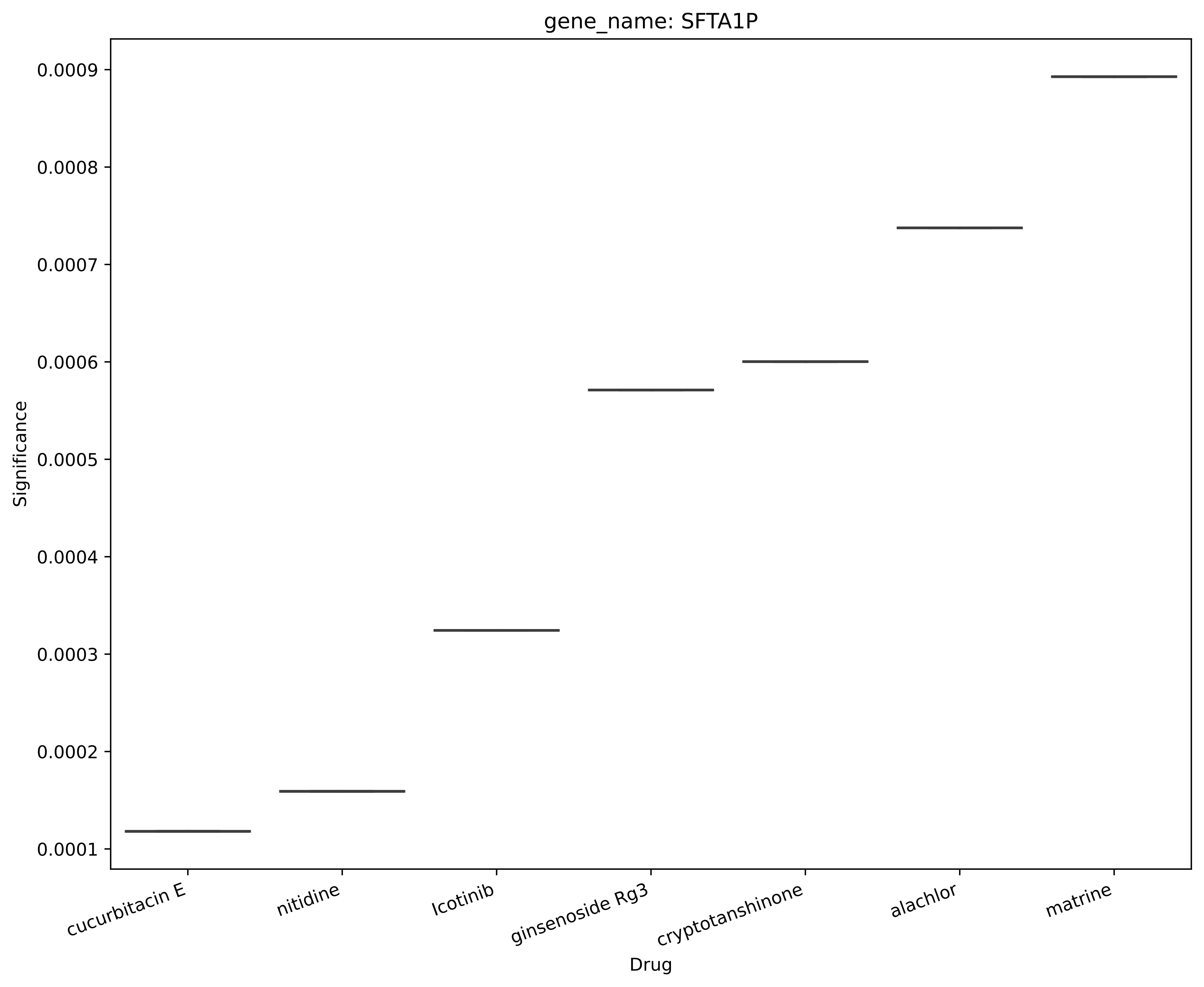
gene_name: SFTA1P
Significance
0.0009
0.0008
0.0007
0.0006
0.0005
0.0004
0.0003
0.0002
0.0001
cucurbitacin E
nitidine
Icotinib
ginsenoside Rg3
cryptotanshinone
alachlor
matrine
Drug

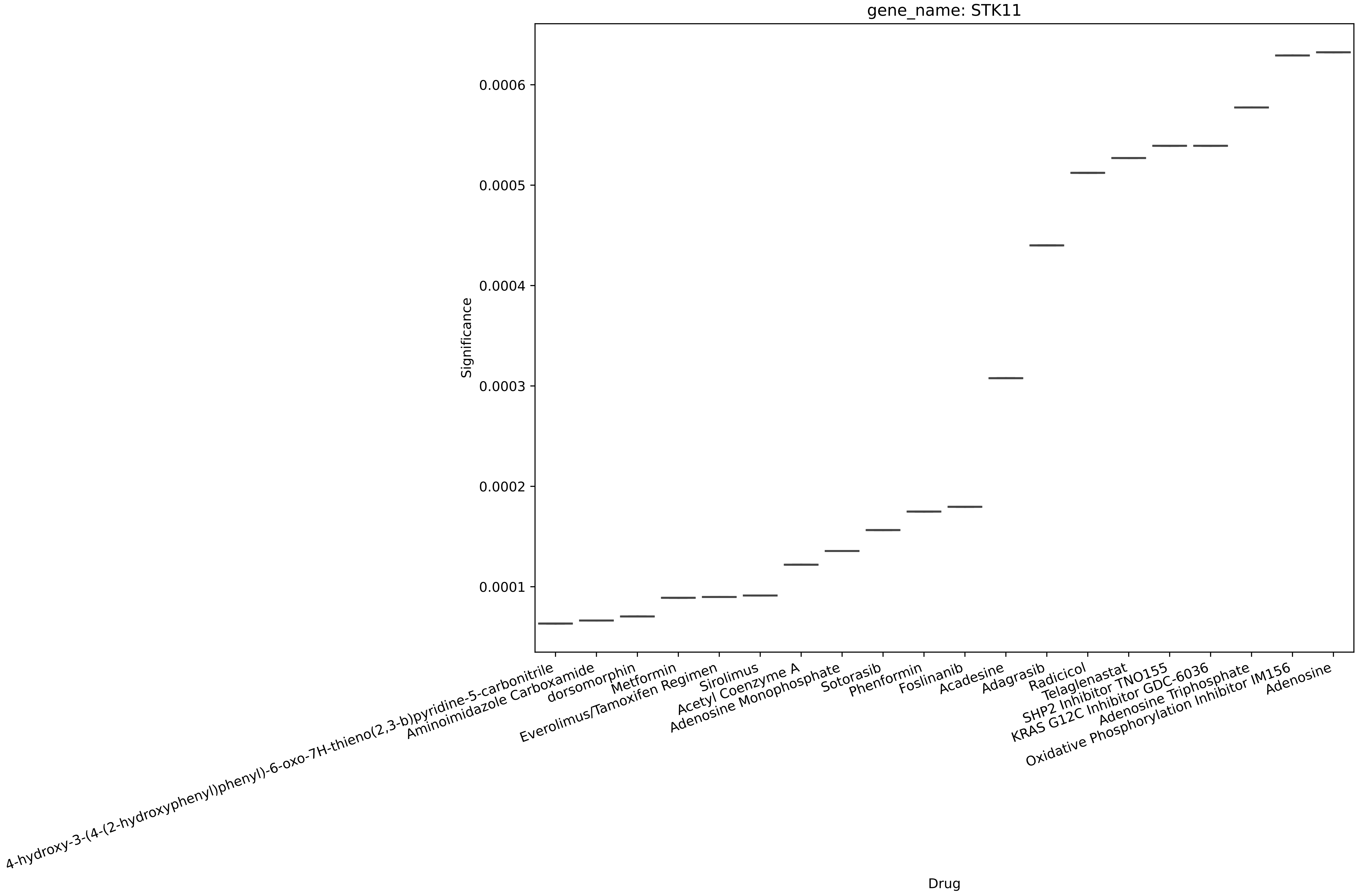
gene_name: STK11
Significance
0.0006
0.0005
0.0004
0.0003
0.0002
0.0001
Drug
4-hydroxy-3-(4-(2-hydroxyphenyl)phenyl)-6-oxo-7H-thieno(2,3-b)pyridine-5-carbonitrile
Aminoimidazole Carboxamide
dorsomorphin
Metformin
Everolimus/Tamoxifen Regimen
Sirolimus
Acetyl Coenzyme A
Adenosine Monophosphate
Sotorasib
Phenformin
Foslinanib
Acadesine
Adagrasib
Radicicol
Telaglenastat
SHP2 Inhibitor TNO155
KRAS G12C Inhibitor GDC-6036
Adenosine Triphosphate
Oxidative Phosphorylation Inhibitor IM156
Adenosine

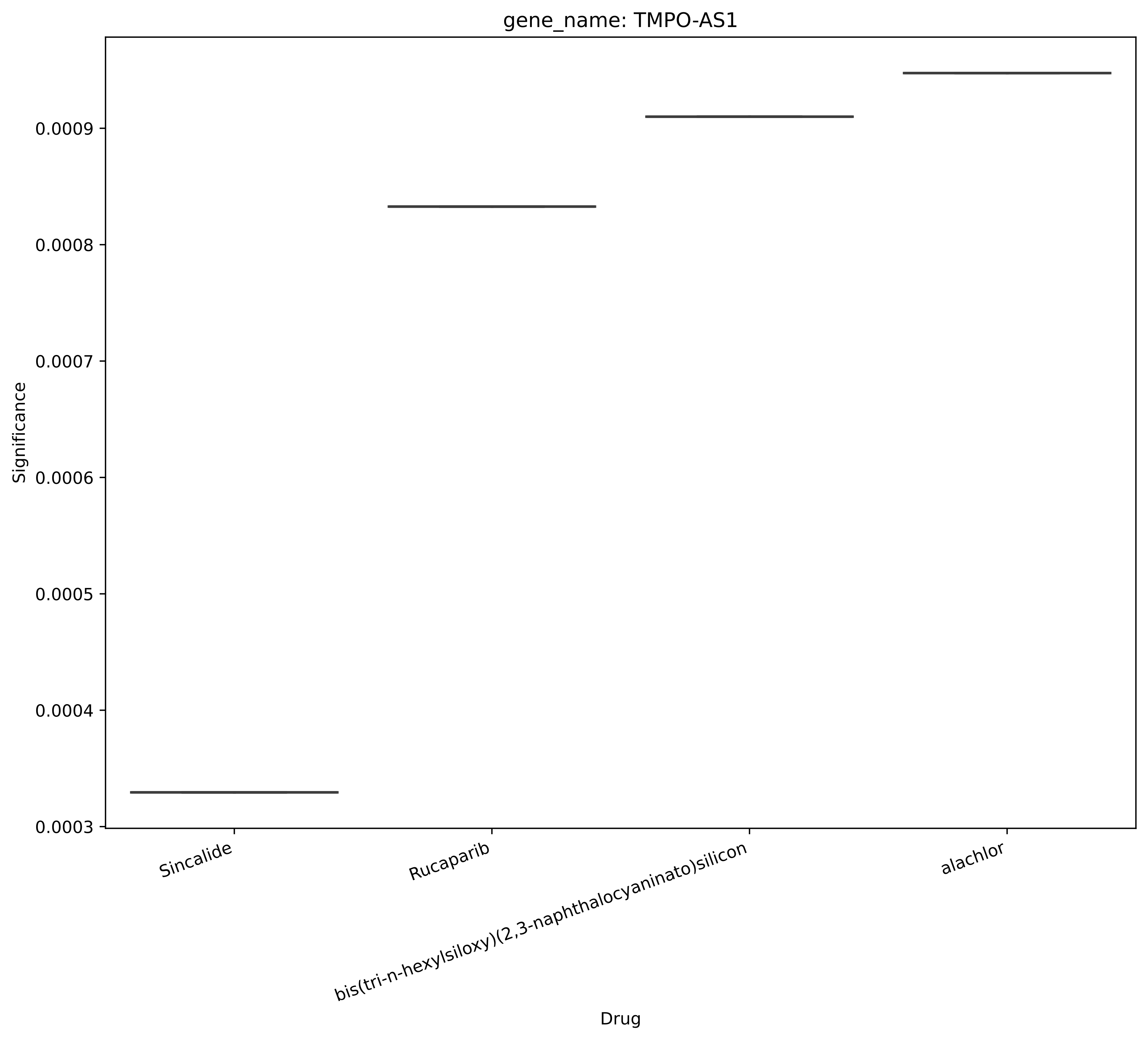
gene_name: TMPO-AS1
Significance
0.0009
0.0008
0.0007
0.0006
0.0005
0.0004
0.0003
Sincalide
Rucaparib
bis(tri-n-hexylsiloxy)(2,3-naphthalocyaninato)silicon
alachlor
Drug

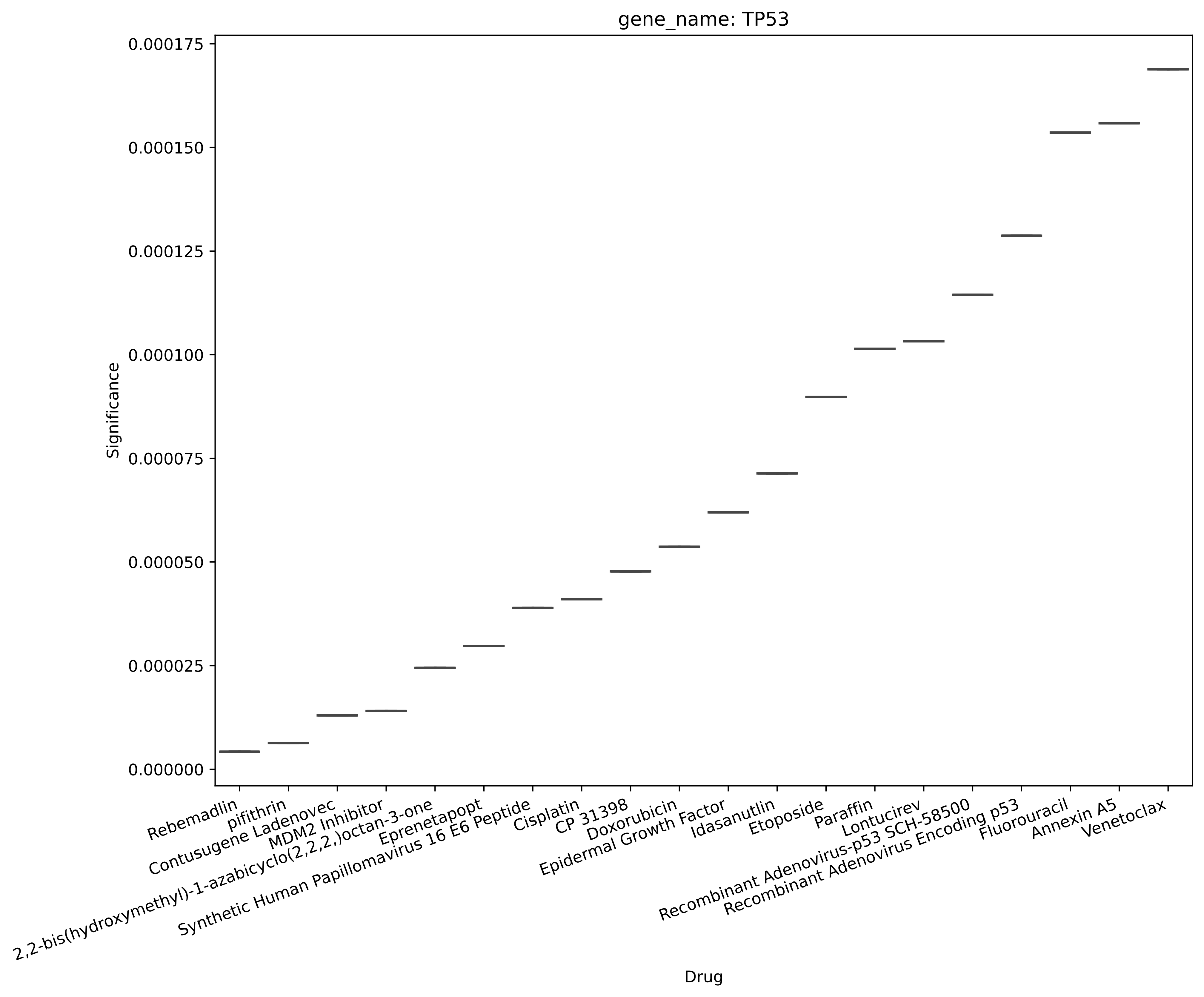

gene_name: TP53
Significance
0.000175
0.000150
0.000125
0.000100
0.000075
0.000050
0.000025
0.000000
Drug
Rebemadlin
pifithrin
Contusugene Ladenovec
MDM2 Inhibitor
2,2-bis(hydroxymethyl)-1-azabicyclo(2,2,2,)octan-3-one
Eprenetapopt
Synthetic Human Papillomavirus 16 E6 Peptide
Cisplatin
CP 31398
Doxorubicin
Epidermal Growth Factor
Idasanutlin
Etoposide
Paraffin
Lontucirev
Recombinant Adenovirus-p53 SCH-58500
Recombinant Adenovirus Encoding p53
Fluorouracil
Annexin A5
Venetoclax

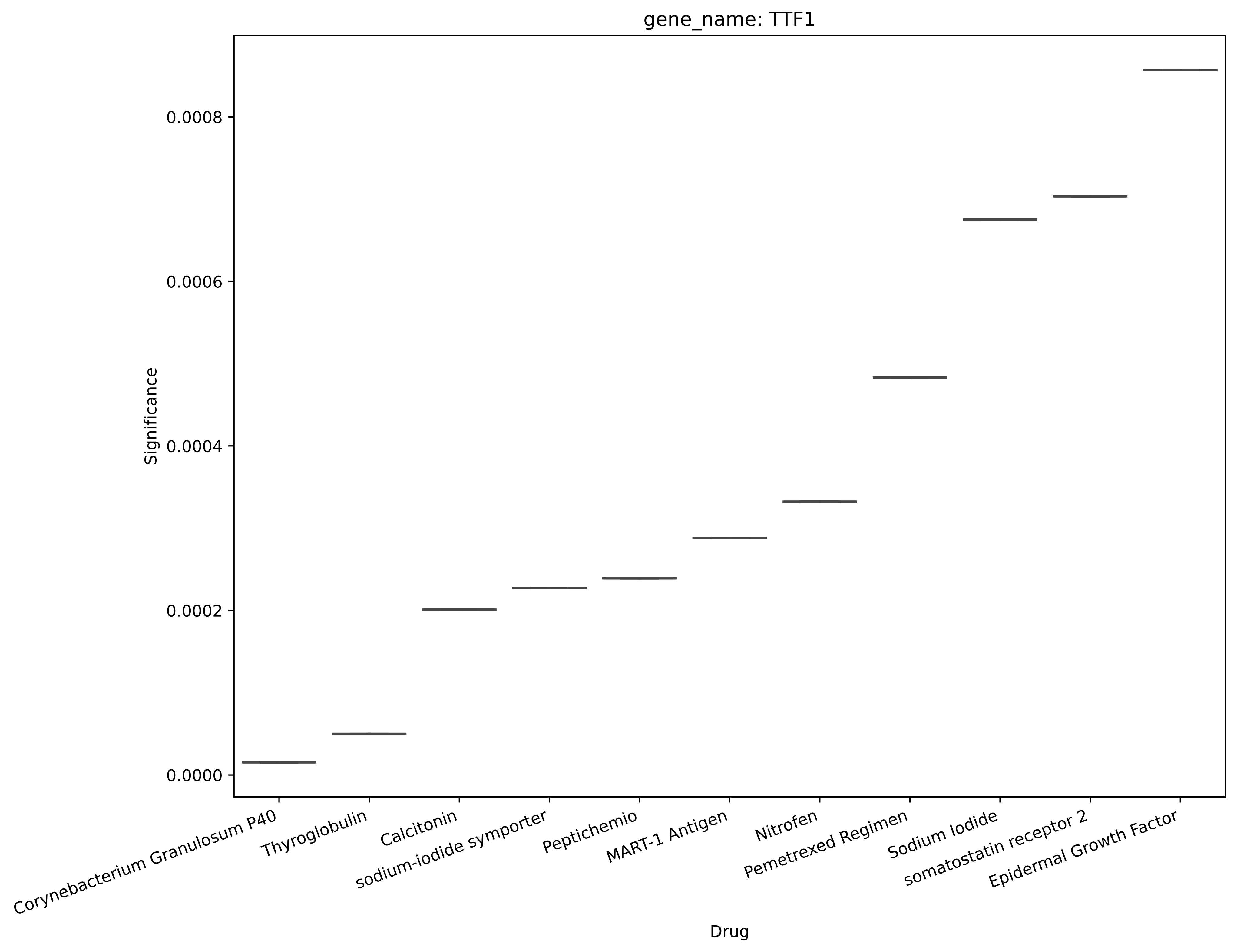

gene_name: TTF1
Significance
0.0000
0.0002
0.0004
0.0006
0.0008
Corynebacterium Granulosum P40
Thyroglobulin
Calcitonin
sodium-iodide symporter
Peptichemio
MART-1 Antigen
Nitrofen
Pemetrexed Regimen
Sodium Iodide
somatostatin receptor 2
Epidermal Growth Factor
Drug

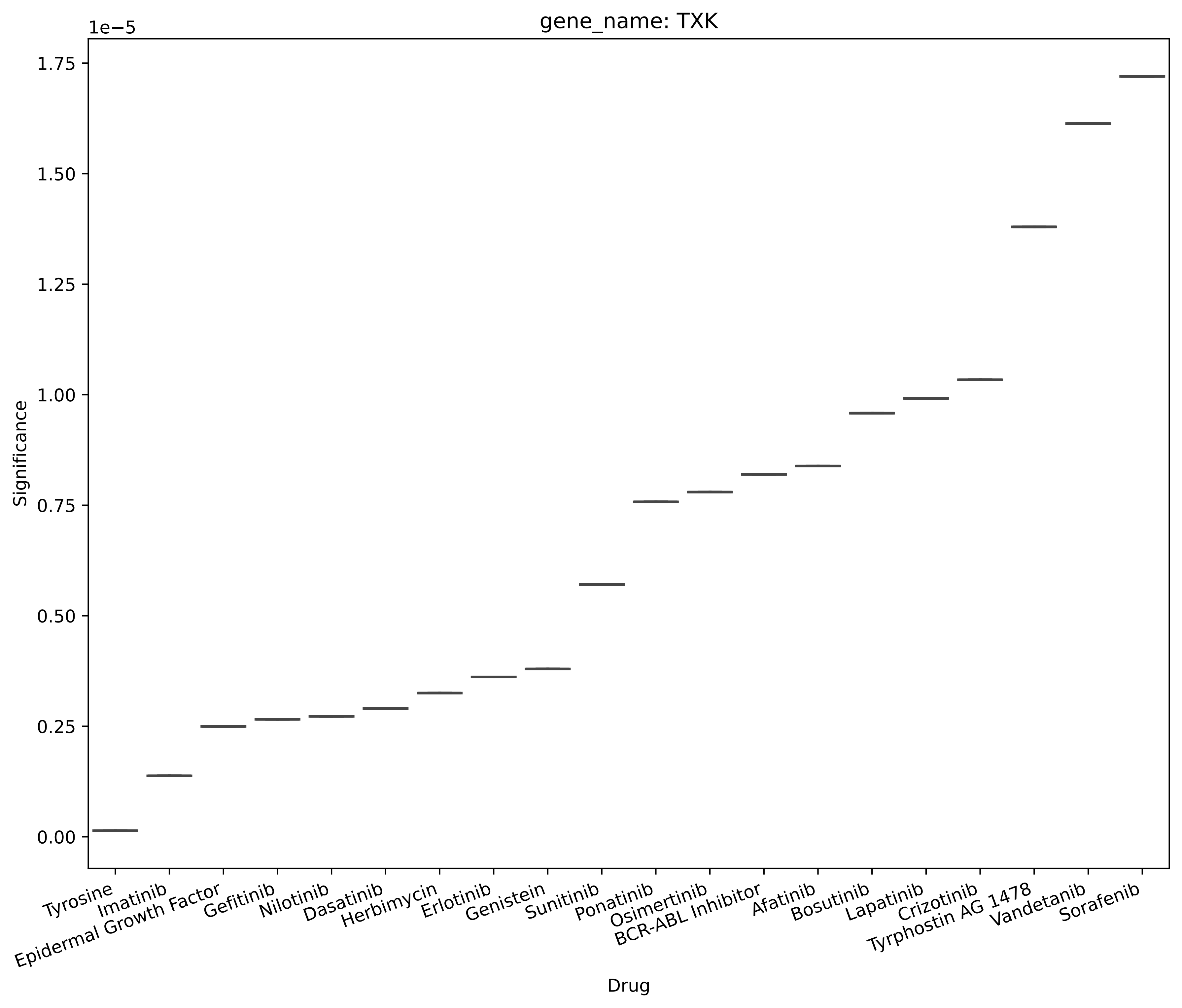
gene_name: TXK
1e−5
1.75
1.50
1.25
1.00
0.75
0.50
0.25
0.00
Significance
Tyrosine
Imatinib
Epidermal Growth Factor
Gefitinib
Nilotinib
Dasatinib
Herbimycin
Erlotinib
Genistein
Sunitinib
Ponatinib
Osimertinib
BCR-ABL Inhibitor
Afatinib
Bosutinib
Lapatinib
Crizotinib
Tyrphostin AG 1478
Vandetanib
Sorafenib
Drug

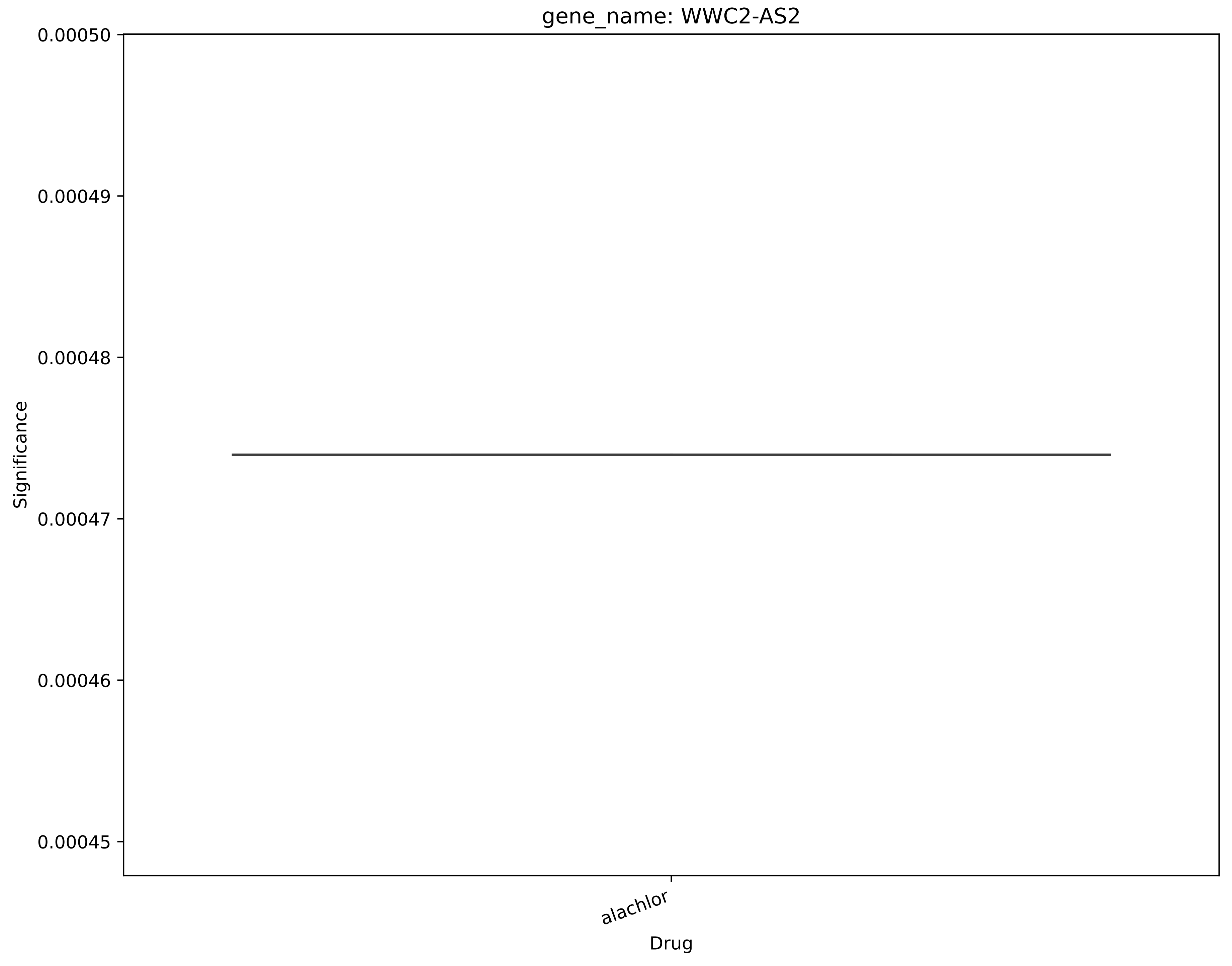
gene_name: WWC2-AS2
0.00050
0.00049
0.00048
0.00047
0.00046
0.00045
Significance
alachlor
Drug

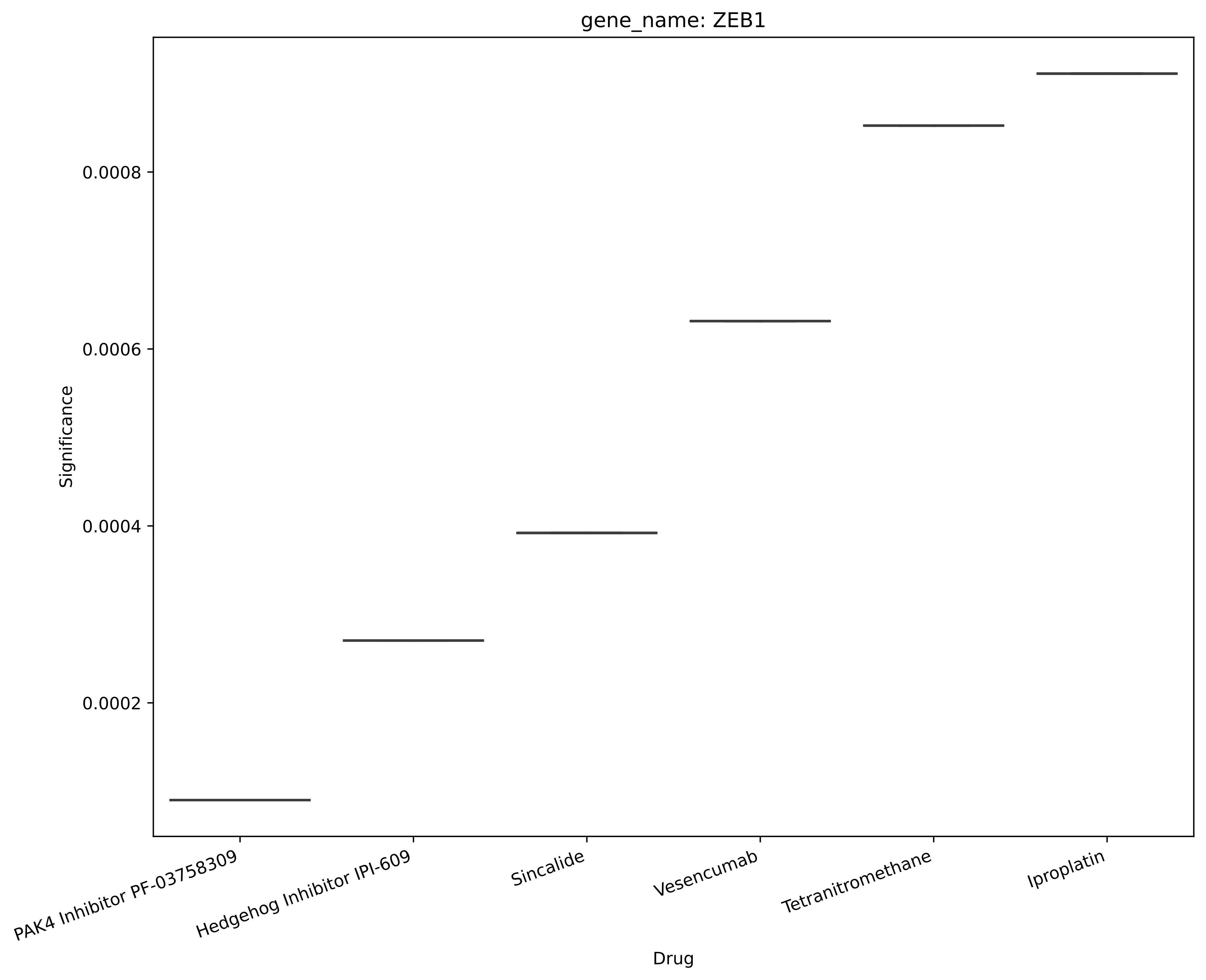

gene_name: ZEB1
Significance
0.0008
0.0006
0.0004
0.0002
PAK4 Inhibitor PF-03758309
Hedgehog Inhibitor IPI-609
Sincalide
Vesencumab
Tetranitromethane
Iproplatin
Drug
